# Plasticity of Human Microglia and Brain Perivascular Macrophages in Aging and Alzheimer’s Disease

**DOI:** 10.1101/2023.10.25.23297558

**Authors:** Donghoon Lee, James M. Vicari, Christian Porras, Collin Spencer, Milos Pjanic, Xinyi Wang, Seon Kinrot, Philipp Weiler, Roman Kosoy, Jaroslav Bendl, N M Prashant, Konstantina Psychogyiou, Periklis Malakates, Evelyn Hennigan, Jennifer Monteiro Fortes, Shiwei Zheng, Karen Therrien, Deepika Mathur, Steven P. Kleopoulos, Zhiping Shao, Stathis Argyriou, Marcela Alvia, Clara Casey, Aram Hong, Kristin G. Beaumont, Robert Sebra, Christopher P. Kellner, David A. Bennett, Guo-Cheng Yuan, George Voloudakis, Fabian J. Theis, Vahram Haroutunian, Gabriel E. Hoffman, John F. Fullard, Panos Roussos

## Abstract

The complex roles of myeloid cells, including microglia and perivascular macrophages, are central to the neurobiology of Alzheimer’s disease (AD), yet they remain incompletely understood. Here, we profiled 832,505 human myeloid cells from the prefrontal cortex of 1,607 unique donors covering the human lifespan and varying degrees of AD neuropathology. We delineated 13 transcriptionally distinct myeloid subtypes organized into 6 subclasses and identified AD-associated adaptive changes in myeloid cells over aging and disease progression. The GPNMB subtype, linked to phagocytosis, increased significantly with AD burden and correlated with polygenic AD risk scores. By organizing AD-risk genes into a regulatory hierarchy, we identified and validated *MITF* as an upstream transcriptional activator of *GPNMB*, critical for maintaining phagocytosis. Through cell-to-cell interaction networks, we prioritized *APOE-SORL1* and *APOE-TREM2* ligand-receptor pairs, associated with AD progression. In both human and mouse models, *TREM2* deficiency disrupted GPNMB expansion and reduced phagocytic function, suggesting that GPNMB’s role in neuroprotection was *TREM2*-dependent. Our findings clarify myeloid subtypes implicated in aging and AD, advancing the mechanistic understanding of their role in AD and aiding therapeutic discovery.

## Main

Despite the quantifiable neuropathology of β-amyloid plaques (Aβ) and neurofibrillary tangles (NFTs) (*1*), the exact neurobiological mechanisms underlying Alzheimer’s disease (AD) remain elusive. Brain myeloid-origin immune cells, including microglia and perivascular macrophages (PVMs), play crucial roles in the pathogenesis of AD (*2–9*), providing neuroprotective benefits by clearing lesions, but also exacerbating the disease through the induction of excessive neuroinflammation (*10*). While previous studies utilizing single-nucleus/- cell RNA sequencing (snRNA-seq/scRNA-seq) have made significant progress describing complex functional roles of murine and human microglia in AD (*5*, *11–14*), challenges with characterizing the wide spectrum of microglial heterogeneity and identifying more nuanced AD-associated subtypes still remain (*15*), largely due to limited sample sizes and differences in the single-cell technologies used. Among the issues that arise is the failure of nuclear fractions in snRNA-seq from frozen tissue to capture key genes related to microglial adaptation and response to pathogenic lesions (*16*). Moreover, microglia are highly reactive cells, and describing their adaptive nature using scRNA-seq in cells isolated from fresh tissue is challenging (*17*). To overcome those limitations, we present two independent human myeloid cohorts generated at single-cell resolution from the prefrontal cortex (PFC). In the first cohort, we isolated viable *ex-vivo* human myeloid cells from fresh postmortem PFC and deeply profiled both nuclear and cytoplasmic RNA. The second cohort focused on the breadth of the transcriptome, profiling human myeloid nuclei from a large number of demographically diverse frozen cortical tissues. By considering both the depth and the breadth of the human myeloid transcriptome, we establish a reproducible taxonomy and demonstrate the importance of microglia and PVM plasticity throughout the lifespan, across different stages of AD pathological and clinical severity, and genetic liability.

### Cellular taxonomy of human myeloid cells

In total, we profiled 832,505 human myeloid cells from the PFC of 1,607 unique donors. The first dataset, named FreshMG, includes samples from fresh autopsy tissue specimens of 137 unique postmortem donors recruited from two brain banks and contains individuals displaying varying degrees of AD neuropathology as well as controls (**Fig. 1A, Supplementary Fig. S1A**). FreshMG donors are aged between 26 and 107 years (average 80.7 years), comprising 76 females and 61 males. To enrich for myeloid cells, viable CD45+ cells were isolated via fluorescence-activated cell sorting (FACS). In addition, for a subset (n=3 donors, each with 8 technical replicates), we profiled surface-level protein markers using CITE-seq (*18*), using a panel of 154 unique antibodies, resulting in a total of 161 scRNA-seq libraries from fresh brain specimens. Following rigorous QC and initial clustering, we found a large, relatively homogeneous, cluster of myeloid cells along with small subsets of co-purified immune cells, such as monocytes, neutrophils, T, NK, and B cells. The myeloid cluster consisted of 543,012 microglia and PVMs robustly expressing 23,740 genes (**Supplementary Fig. S1E**).

**Figure 1.**
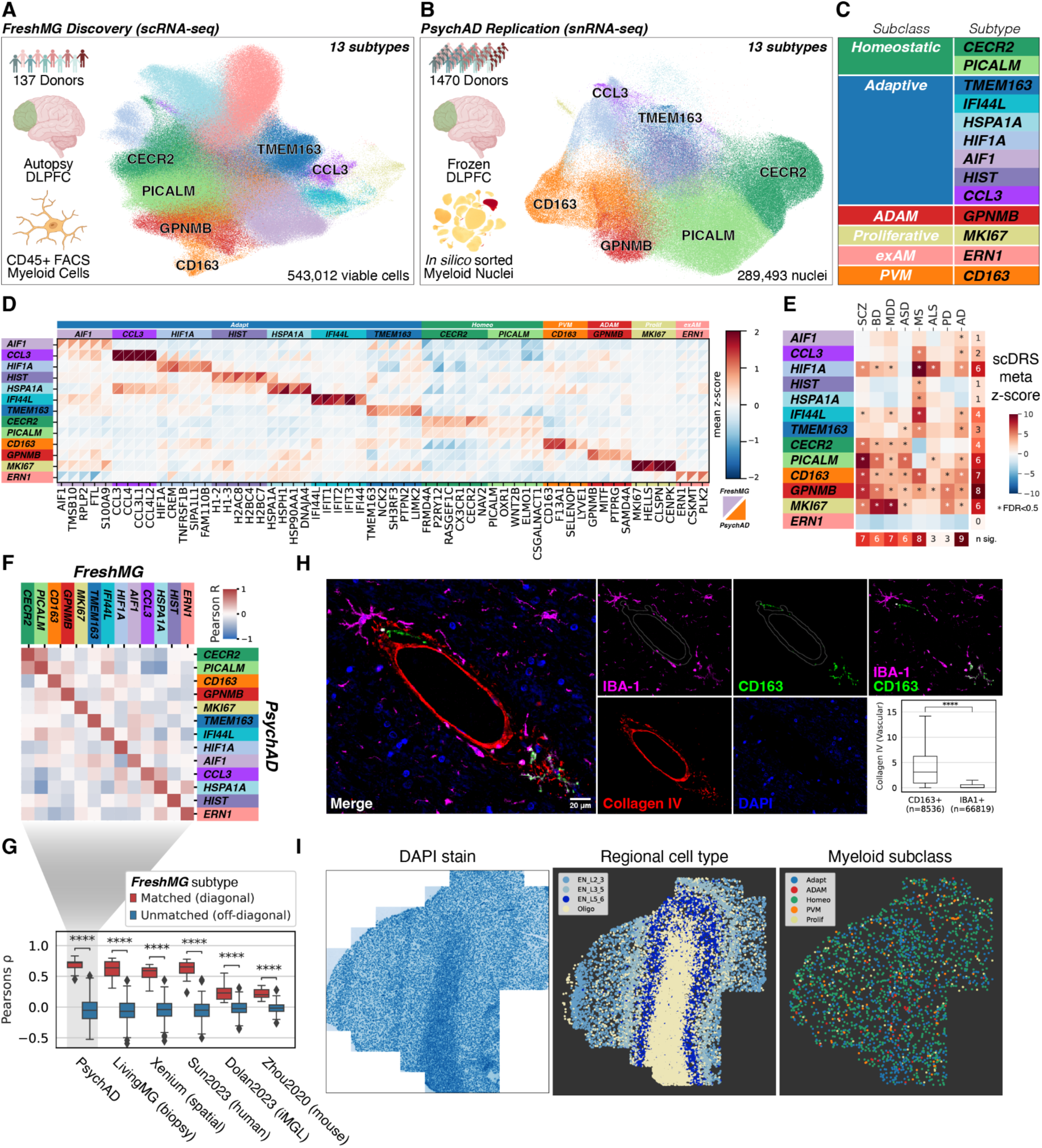
Overview of the human myeloid single-cell atlas. **(A)** the *FreshMG* discovery cohort (scRNA-seq) using live human myeloid cells from postmortem PFC and **(B)** the *PsychAD* replication cohort (snRNA-seq) using flash-frozen PFC tissues and *in-silico* sorted for microglia and PVMs. **(C)** Unified taxonomy of human myeloid subtypes. **(D)** Subtype-specific marker gene expression. Z-score normalized. Upper-triangle: *FreshMG*. Lower-triangle: *PsychAD*. **(E)** Enrichment of heritable disease risk (scDRS) by subtype using GWAS of 8 brain diseases. Meta-analysis between FreshMG and PsychAD. The asterisk denotes FDR < 0.05. SCZ: schizophrenia, BD: bipolar disorder, MDD: major depressive disorder, ASD: autism spectrum disorder, MS: multiple sclerosis, ALS: amyotrophic lateral sclerosis, and PD: Parkinson’s disease. **(F)** Pairwise Pearson correlation of the subtype-level taxonomy between *FreshMG* and *PsychAD* datasets using highly variable genes common in both datasets. **(G)** Validation of human myeloid taxonomy using independent, multi-modal, and published datasets. Human (*14*), iMGL: iPSC-derived microglia (*23*), and Mouse (*27*). Pairwise comparison of subtype-level taxonomy against the FreshMG annotation. Mann–Whitney U test between matched (diagonal) and unmatched (off-diagonal) subtypes. ****: p ≤ 1.0e-4. **(H)** Representative image of Akoya PhenoCycler multiplex immunofluorescence results showing CD163^+^/IBA-1^+^ cells are enriched near blood vessels (outlined by gray line), labeled by Collagen IV. Scale bar 20 µm. **(I)** Representative slide of Xenium *in situ* spatial transcriptomics data. Left: DAPI, Middle: laminar distribution of neuronal cell types, Right: distribution of myeloid cells annotated by subclasses.

The second dataset, named PsychAD, consists of frozen prefrontal cortex specimens and includes cases and controls from a cohort of 1,470 unique donors (**Fig. 1B**). PsychAD donors were aged between 0 and 108 years, (average 71.3 years), comprising 761 females and 709 males (**Supplementary Fig. S1B**). Frozen samples were subject to snRNA-seq profiling from which microglia and PVMs were sorted *in silico* after basic clustering. After rigorous QC, we identified 289,493 microglia and PVM nuclei robustly expressing 34,890 genes (**Supplementary Fig. S1F**). Next, we aligned and harmonized the scale of clinical variables to facilitate annotation of both datasets (**Methods**) and saw a strong positive correlation with measures of the severity of AD neuropathology, namely diagnostic certainty of AD, Consortium to Establish a Registry for Alzheimer’s Disease (CERAD) (*19*), and Braak stage (*20*) (**Supplementary Fig. S1C**). In contrast, the clinical measures of dementia severity were less well correlated with AD.

Our primary objective was to establish a comprehensive cellular taxonomy that is robust and reproducible; however, cross-validating these independent and large-scale single-cell datasets, each with a distinct transcriptomic origin (whole cell vs. nuclei), posed technical challenges. To overcome these, we devised an iterative cross-validation strategy, which involved establishing a reference state and validating it independently until both datasets were in agreement (**Methods**). Utilizing the FreshMG dataset, which provides comprehensive transcriptomic profiles from both nuclear and cytosolic fractions, we identified functionally distinct phenotypes of microglia and PVMs. Subsequently, we cross-validated the presence of these reference subtypes in the frozen specimen snRNA-seq PsychAD dataset. Our iterative process converged on 13 functionally distinct subtypes of human myeloid cells (**Fig. 1C, Supplementary Fig. S2A, Supplementary Tables S1-2**), and comparison between FreshMG and PsychAD revealed a high degree of consistency between the two cohorts, as evidenced by an average Pearson correlation of 0.77 across all identified subtypes (**Fig. 1F**). This rigorous methodology ensured the accuracy and reliability of our cellular taxonomy, laying a solid foundation for further analyses.

We grouped the cells using two levels of taxonomic hierarchy; the 13 distinct subtypes under six broad functional subclasses of human myeloid cells: Homeostatic (green), Adaptive (blue), Proliferative (yellow), AD-Associated or ADAM (red), *ex-vivo* Activated Microglia or exAM (pink), and PVM (orange) (**Fig. 1C**). Each subtype is associated with specific markers that not only aid in their identification but also hint at their functional significance (**Fig. 1D, Supplementary Fig. S2D**). Within the homeostatic microglia subclass, we highlight two subtypes, CECR2 and PICALM, both of which are associated with the regulation of GTPase activity. Homeostatic microglia make up the largest proportion of myeloid cells (**Supplementary Figs. S2B-C**) and express microglia-specific canonical markers such as *P2RY12* and *CX3CR1*. The CECR2 subtype uniquely expresses *CECR2* and *NAV2*, with other genes pointing towards cellular maintenance, phagocytosis, cell migration, and adhesion. The PICALM subtype shows elevated expression of *PICALM* and *ELMO1*, suggesting roles in the regulation of the immune response.

We identified 7 specialized microglial subtypes, each exhibiting unique adaptive responses to neuro-environmental cues. In general, the gene signatures across these adaptive microglia underscored an enhancement in antigen processing and presentation programs and the facilitation of MHC protein complex assembly. The CCL3 subtype is characterized by the upregulation of chemotactic genes, most notably the inflammatory cytokines *CCL3*, *CCL4*, and interleukin 1 beta (*IL1B*). In addition, the *IFI44L* subtype is enriched in interferon-inducible genes, like *IFIT1*, *IFIT2*, and *IFIT3*, suggesting a role in the antiviral innate immune response.

The AIF1, HIF1A, and HIST clusters share a common gene program related to immunoglobulin-mediated immune response, while the TMEM163 cluster focuses on antigen processing and presentation via MHC II. The final adaptive cluster, HSPA1A, is enriched for gene signatures responsible for adaptive response to unfolded protein, which is characterized by elevated activity of heat shock proteins and cellular stress response, with a potential role in AD neuropathology (*21*). In addition, we identified a subtype, the GPNMB, which is predominantly observed in individuals with AD (*22*, *23*). These AD-associated microglia (ADAM) feature elevated expression of glycoprotein non-metastatic melanoma protein B (*GPNMB*), microphthalmia-associated transcription factor (*MITF*), and protein tyrosine phosphatase receptor type G (*PTPRG*) genes, and functional enrichment analysis suggests increased phagocytic activity is a hallmark of these cells. Consistent with previous studies (*11*), we also identified a cluster of proliferative cells, MKI67, that is highly enriched in cell-cycle dependent genes (*STMN1*, *MKI67*, *TOP2A*). Lastly, we report a cluster, ERN1, showing specific expression of *ERN1* and *PLK2* genes that resemble activation patterns of exAM (*17*) (**Supplementary Information**). In addition to microglial subtypes, we identified a PVM cluster, named CD163, expressing a unique set of known PVM-specific markers, notably *CD163* and *F13A1*. The CD163 cluster displayed a significant enrichment of genes involved in endocytic processes, emphasizing its priming for receptor-mediated endocytosis and phagocytosis. While we observe a close similarity between ADAM and PVM clusters (**Supplementary Fig. S2A**), we found a clear separation between the two when we enriched for conserved murine disease-associated microglia (DAM) signatures as well as human DAM signature from iPSC-derived microglia (*5*, *23–25*) (**Supplementary Fig. S3A**).

We further annotated myeloid subtypes by estimating the enrichment with polygenic risk scores of heritable traits at single-cell resolution (`scDRS`; **Methods**; **Supplementary Fig. S3B, Supplementary Table S15**). We extended the analysis to a set of the brain related diseases beyond AD including schizophrenia (SCZ), bipolar disorder (BD), major depressive disorder (MDD), autism spectrum disorder (ASD), multiple sclerosis (MS), amyotrophic lateral sclerosis (ALS), and Parkinson’s disease (PD). The polygenic risk scores for each trait were highly reproducible between the FreshMG and PsychAD cohorts with AD and MS having the greatest correlation (**Supplementary Fig. S3C**). The meta-analysis of both FreshMG and PsychAD cohorts indicated that the 9 subtypes of myeloid cells were significantly associated with heritable AD risk, which was the largest of all brain diseases followed by MS, SCZ, and MDD (**Fig. 1E**). Notably, the GPNMB subtype had the widest coverage showing significant heritable risks for all 8 diseases.

### Multi-modal validation of human myeloid taxonomy

To show the utility of our annotation as the reference human myeloid taxonomy, we validated the reproducibility of 13 myeloid subtypes using several independent datasets. First, using a published human microglia dataset (*14*), we assessed the similarity of our taxonomy to existing microglia annotations. While we found 8 of their microglial states, including their brain-associated macrophage (BAM), resemble our subtypes (**Supplementary Fig. S3D**), the alignments were moderate for the remaining 5 states. After re-annotating their nuclei using our taxonomy as the reference (**Methods**), we confirmed the presence of all 13 subtypes (**Fig. 1G, Supplementary Fig. S3G**). We also discovered the subtype composition was comparable to our PsychAD snRNA-seq dataset (**Supplementary Figs. 2B-C**).

Since the taxonomy was established based on post-mortem tissues, we needed to ensure the taxonomy was not biased for post-mortem effects, and it can be reproduced using living brain tissues. Independent from the FreshMG and PsychAD cohorts, we generated an additional scRNA-seq dataset, called LivingMG, from brain biopsies, which were obtained from 25 unique human donors (26 libraries; 97,828 cells after QC) diagnosed with spontaneous intracerebral hemorrhage (ICH) (*26*). The brain tissue was collected during treatment and processed in an identical manner to the fresh autopsy material. It’s important to note that cortical biopsy samples were obtained from a site distal to the site of the hemorrhage and, in the absence of a secondary diagnosis, are considered neurotypical controls. We annotated myeloid cells using the taxonomy derived from the FreshMG dataset and confirmed the presence of all 13 subtypes in living cells. (**Fig. 1G**, **Supplementary Fig. S3E, S3J**).

Since the taxonomy was primarily derived from sc/snRNA-seq datasets, we utilized different technology and modalities to confirm the robustness of our myeloid taxonomy. To validate the spatial context, we conducted deep single-cell phenotyping and spatial analysis using multiplexed imaging assay (Akoya PhenoCycler) and demonstrated, for example, PVMs colocalize around blood vessels via staining for CD163 (**Fig. 1H**). Subsequently, we performed spatial transcriptomic characterization using the Xenium *in situ* technology on 11 tissue slides obtained from 8 individual donors (**Methods**). A custom panel of 366 genes, including both a pre-designed human brain panel and additional markers for myeloid subtypes, was used to further characterize the myeloid taxonomy (**Fig. 1I**). We showed the presence of 5 major subclasses excluding the exAM, which was not expected to be present in cryosectioned tissue (**Fig. 1G, Supplementary Figs. S3F, S4A-B**). While the resolution was limited in the Xenium data, we were able to stratify robust subtypes via stability analysis (**Methods**) and validated the presence of myeloid subtypes.

Lastly, we applied a multi-omic assay to further characterize the myeloid subtypes. We employed CITE-seq, jointly quantifying the transcriptome and 154 unique cell-surface proteins, to assess the preservation of the functional hierarchical structure at the protein-level. Using this approach, we confirmed the presence of distinct proteomic patterns for each myeloid subtype (**Supplementary Fig. S3K**). For example, within the homeostatic microglia subclass, the CECR2 subtype expressed *CD99* and *ITGB3*, while the PICALM subtype expressed *CLEC4C* and *TNFRSF13C* proteins as their markers. Likewise, the PVM cluster showed distinct surface markers, *CD163* and *CCR4*, while the ADAM cluster was specific for *CD9* and *CD44* proteins.

In summary, we used both external and independent datasets, as well as multi-omic modalities, to validate that the taxonomy is robust and consistent irrespective of the tissue source.

### Variation in human myeloid subtype composition on aging and AD

After determining 13 distinct subtypes of human brain myeloid cells, we examined the compositional variation of myeloid subtypes that are associated with aging in a subset of neurotypical donors who were free of dementia and diagnostic neuropathology from the FreshMG and PsychAD datasets. We normalized the subtype count ratio data using the centered log-ratio transformation and modeled using a linear mixed model, accounting for technical and demographic variables (**Methods**). Notably, the two homeostatic microglia subtypes displayed opposing trajectories with respect to aging (**Figs 2A left, D**). The CECR2 subtype showed progressive decline while the PICALM subtype showed a gradual increase with age. In addition, we saw an overall increase in the proportions of the ADAM and PVM subtypes with age. These findings were replicated using published human microglia snRNA-seq dataset (*14*) (**Supplementary Fig. S4C**). In contrast, we observed an age-related decline in the CCL3 subtype, indicating a possible reduction of chemotactic microglia in older brains. In parallel, we investigated sex-dependent variation in human myeloid subtypes, with or without taking age into consideration, but did not find any statistically significant compositional differences between males and females (**Figs. 2A middle, right**).

**Figure 2.**
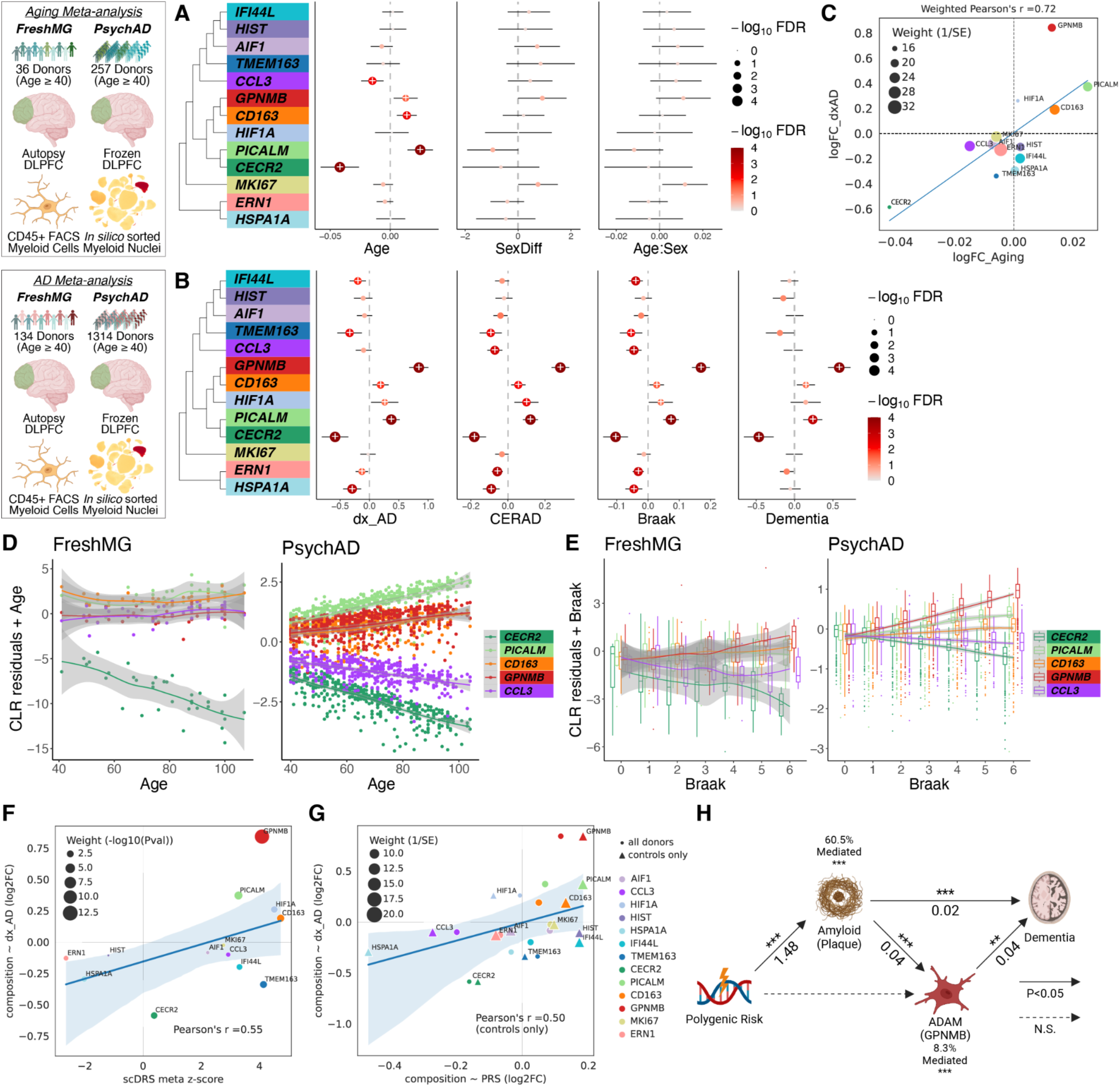
Variation in human myeloid subtype composition. **(A)** Compositional variation of myeloid subtypes by age, sex, and the interaction between age and sex using disease-free subset. CLR transformed composition data was modeled using a linear mixed model accounting for technical batch effects including tissue sources and sequencing pools and donor effects including age, sex, genetic ancestry, and PMI (see **Methods** on `crumblr`). Fixed effect meta-analysis using results from FreshMG and PsychAD cohorts. **(B)** Compositional variation of myeloid subtypes by four different neuropathological measures of the AD progression; diagnosis (dx_AD), CERAD, Braak staging, and dementia status, after accounting for technical and donor-level covariates. Fixed effect meta-analysis using both FreshMG and PsychAD cohorts. **(C)** Comparison of compositional variation between disease-free aging and AD. Subtypes were weighted by the inverse of standard error. **(D)** Covariate adjusted compositional variation with disease-free aging. CLR: centered-log-ratio. **(E)** Covariate adjusted compositional variation with Braak staging. **(F)** Correlation between `scDRS` meta z-scores and `crumblr` estimate of compositional variation by dx_AD as a coefficient. Weighted Pearson’s correlation using average -log10(P-value) as weights. **(G)** Correlation between `crumblr` estimate of compositional variation by PRS as a coefficient against `crumblr` estimate of compositional variation by dx_AD as a coefficient. Weighted Pearson’s correlation using inverse of average of standard error as weights. Circle denotes `crumblr` analysis using all donors while triangle denotes `crumblr` analysis using controls only. **(H)** Causal mediation analysis using PRS, Aβ plaque, composition of the GPNMB subtype, and clinical dementia status. ***: p ≤ 1.0e-3, **: p ≤ 1.0e-2, NS: p > 0.05.

Next, we examined the variation of subtype composition during onset and progression of AD. To minimize the effect of younger brains, we limited the analysis to donors 40 years and older, resulting in a dataset composed of 134 donors from the FreshMG and 1,314 donors from the PsychAD cohort. We first evaluated the involvement of myeloid subtypes using the centered log-ratio transformed count ratio data after accounting for technical and demographic variables (**Methods**). Overall, irrespective of different measures of AD phenotypes (dx_AD, CERAD, Braak, and Dementia), we observed robust changes in subtype proportions in both FreshMG and PsychAD cohorts (Fig. 2B). Similar to normal aging, two homeostatic subtypes showed opposing trends, where the CECR2 subtype showed a progressive decline with increasing AD burden while the PICALM subtype showed a gradual increase. While the trends were observed during the early stages of AD, a more substantial divergence occurred after Braak stage 3 (Fig. 2E). Likewise, we observed a consistent increase in the proportion of the PVM subtype. The most notable difference in the compositional variation of AD phenotypes compared to aging was the GPNMB subtype. The GPNMB subtype was an outlier and showed the largest effect size across all 4 AD phenotypes, suggesting that proliferation of the GPNMB subtype is a hallmark of AD (Fig. 2C). We further supported our findings by replicating the compositional variation analysis with previously published data (*14*). Consistent with our findings, we observed the GPNMB subtype was increasing in proportion while Homeo_CECR2 was decreasing with severe AD neuropathology (**Supplementary Figs. S4C-E**).

### Causal mediation analysis of polygenic AD risk scoring and myeloid subtypes

Having established that certain myeloid subtypes are enriched for AD genetic risk (Fig. 1E) and that their compositional landscape shifts in the presence of AD (Fig. 2B), we next sought to evaluate the association of per-cell polygenic AD risk scores with the compositional variation observed in AD (`scDRS`; **Methods**, **Supplementary Fig. S3B**). We observed a positive correlation (Pearson’s r = 0.55) indicating that the ratio of subtypes with higher polygenic AD risk scores increases in AD (Fig. 2F**; Supplementary Fig. S4F**). This also suggested heritable risks might play a role in driving the compositional changes of myeloid subtypes.

Next, we leveraged our population-scale cohort to calculate per-donor AD polygenic risk scores (PRS; **Methods**) and to assess how the interindividual variation in AD risk impact changes in myeloid subtype composition (**Supplementary Fig. S3B**). The proportion of the GPNMB subtype was significantly increased with AD PRS (**Supplementary Fig. S4G**). We observed a similar compositional variation between AD phenotype (dx_AD) and PRS (Fig. 2G **circle**), which was not driven by the AD status alone as the same compositional variation was observed using a disease-free subset (Fig. 2G **triangle**).

To further dissect the relationships between genetic risk for AD and the observed changes in GPNMB subtype composition, we conducted a series of causal mediation analyses using the PRS as an instrumental variable (**Methods**). By examining the indirect effects of AD PRS on the GPNMB subtype composition, we aimed to clarify whether the observed cellular changes were driven by genetic predisposition or were a downstream consequence of AD pathology (plaque). Our analysis revealed a significant indirect effect of AD PRS on the GPNMB subtype, mediated through accumulation of Aβ plaques (Average Causal Mediated Effect (ACME) = 0.0254, 95%CI = [0.0137, 0.04], pval<2e-16). This indirect effect accounted for 60.5% of the total effect (pval = 0.034). These findings suggest that the GPNMB subtype variation is more likely a consequence of AD pathology. Furthermore, we observed a significant mediation effect of the GPNMB subtype variation on severity of dementia (8.29% of the total effect mediated, pval = 0.00096), suggesting that modification of this subtype via therapeutics could be a feasible treatment strategy for AD.

### Variation in transcriptional regulation of human myeloid cells on aging and AD

We investigated the transcriptional regulation of human myeloid cells by examining the differential gene expression patterns associated with normal aging, and during the onset and progression of AD. In normal aging (**Supplementary Fig. S5A**), we discovered the increase in expression of the *MS4A6A* gene, a member of the MS4A family of cell membrane proteins, which are involved in the regulation of calcium signaling and have been implicated in neurodegenerative processes (*28*). The age-related gene expression changes for both homeostatic subtypes were enriched with actin filament-based process and actin cytoskeleton organization pathways, supporting their proposed roles in cell adhesion and migration (**Supplementary Fig. S5B**). The CD163 subtype was associated with the increase of cell adhesion processes as well as pathways related to cell proliferation. The gene signatures in the GPNMB subtype were enriched with immune response and activation. Overall, the increased involvement of PVM and ADAM subclasses indicated an upregulation of inflammatory responses in older individuals.

Next, we evaluated genes exhibiting differential expression patterns across four different measures of AD phenotypes (dx_AD, CERAD, Braak, and Dementia) (Fig. 3A). Our analysis led us to discover a set of AD-associated genes, including *PTPRG*, *DPYD*, and *IL15*, which displayed upregulation across all phenotypes capturing more severe AD stages. Pathway enrichment analysis revealed the PICALM and GPNMB subtypes share common pathways related to the regulation of cell adhesion (**Supplementary Fig. S5C**). In contrast, both the CECR2 and CD163 subtypes appear to be associated with negative regulation of cell projection organization.

**Figure 3.**
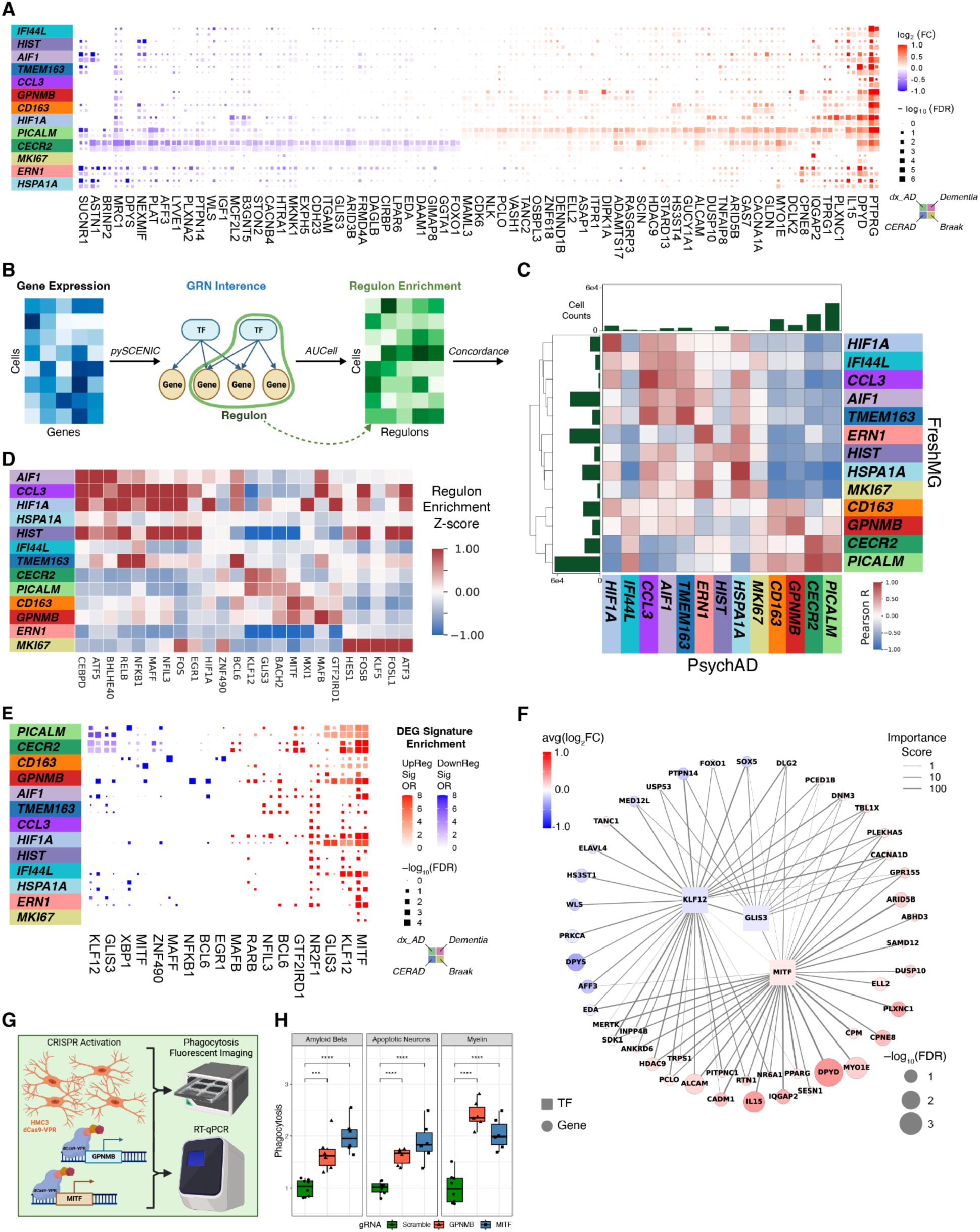
Transcriptional regulation of human myeloid cells. **(A)** Differentially expressed genes by four different measures of AD neuropathology adjusted for technical and donor-level covariates. Fixed effect meta-analysis using both FreshMG and PsychAD cohorts. **(B)** Schematic overview of GRN inference and TF-gene regulon enrichment for prioritization of upstream master regulators of AD. **(C)** Concordance of normalized regulon activity scores (AUCell) between FreshMG and PsychAD cohorts. Pairwise Pearson correlation. **(D)** Enrichment of regulon by subtypes. Meta-analysis of consensus regulon enrichment Z-score with Stouffer’s correction between FreshMG and PsychAD cohorts. Top 3 regulons per each subtype shown. **(E)** Enrichment of AD gene signatures by regulons. Fisher’s exact tests for enrichment of differentially expressed gene signatures in regulon target genes across myeloid subtypes. **(F)** TFs that modulate AD risk genes. Gene regulatory network visualization of *KLF12*, *MITF*, and *GLIS3* TFs and downstream target risk genes. Node colors represent gene expression changes from `dreamlet` analysis. Edge weights represent importance scores inferred from the SCENIC pipeline. **(G)** Schematic of phagocytosis assay. **(H)** Relative level of phagocytosis after CRISPR activation in HMC3 cell line.

Given the strong compositional shifts and gene signatures for AD phenotypes, we tested the presence of AD signatures in bulk microglia RNA-seq data (BulkMG; **Methods**). First, we created myeloid subtype signatures from both the FreshMG and PsychAD datasets by aggregating gene expression by subtype. We then compared the resulting subtype signatures to BulkMG gene expression data, stratified by AD case and control status. Interestingly, the Pearson correlation between subtypes and AD diagnosis clearly reflected the compositional shifts we observed across multiple AD phenotypes (**Supplementary Fig. S5D**). The CECR2, TMEM163, CCL3, and HSPA1A signatures closely correlated with the BulkMG from controls, while the PICALM, CD163, GPNMB, and HIF1A signatures closely matched those from AD cases. These results independently reproduce the observed changes in the myeloid transcriptome during the onset and progression of AD.

To model the dynamic changes that take place during the onset and progression of AD at a molecular level, we expanded our analysis from using discrete donor-level clinical variables to a continuous pseudotime measure by ordering cells along a disease trajectory. We estimated Braak-stage-informed ancestor-progenitor relations between observations through transport maps between neighboring disease stages using Moscot (*29*). We then quantified cell-cell transition probabilities, computed putative drivers, and constructed the disease-stage-informed pseudotime with CellRank 2 (*30*) (**Supplementary Fig. S5E**; **Methods; Supplementary Information**). As expected, we observed an increase in pseudotime with disease progression (**Supplementary Fig. S5F**). Stratified by subtypes, we observed that PICALM homeostatic microglia were assigned larger pseudotime values (late), compared to CECR2 homeostatic cells (early; p-value < 0.001, **Supplementary Fig. S5G**), indicating their association with disease progression and aligning with the compositional variation of AD phenotypes observed earlier. To identify potentially critical stages in disease progression, we compared changes in pseudotime across disease stages for each myeloid subtype (**Methods**). This analysis revealed that the change was most pronounced starting from Braak stage 3 (**Supplementary Fig. S5H**), which was also the critical time point the subtype composition diverged in AD.

### Upstream regulators of AD genes in human myeloid cells

After identifying potential AD risk genes, we analyzed myeloid gene regulatory networks (GRNs) to discover key upstream transcriptional regulators. Using SCENIC (*31*, *32*), we constructed GRNs based on expression data and known transcription factor (TF) binding motifs and defined units of regulatory hierarchy (regulons) (Fig. 3B**, Supplementary Table S12**). Subsequently, we assessed the enrichment of the regulon for each myeloid subtype independently (**Supplementary Table S13**, **Methods**), revealing high concordance between the FreshMG and PsychAD cohorts (Fig. 3C). We then derived combined regulon enrichment scores using meta-analysis (**Methods**) and observed strong regulon subtype-specificity (Fig. 3D). The CECR2 and PICALM homeostatic subtypes were defined by enrichment of *KLF12*, *GLIS3*, and *BACH2* regulons, while the PICALM, CD163, and GPNMB subtypes displayed exclusive enrichment of *MITF* regulon. To link inferred regulons to differentially expressed AD genes, we performed enrichment tests using 4 different types of AD risk signatures (**Methods**). Notably, the target genes of *MITF*, *KLF12*, and *GLIS3* TFs were significantly associated with AD risk profiles in the PICALM, CECR2, GPNMB, and HIF1A subtypes (Fig. 3E). *MITF* was preferentially enriched with upregulated AD signatures, whereas *KLF12* and *GLIS3* were more preferentially associated with downregulated AD signatures. Visualization of the joint *MITF-KLF12-GLIS3* regulon network with AD risk genes revealed coordinated modulation of both up and down-regulated candidate risk genes (Fig. 3F). These findings collectively suggest the coordinated activity of *MITF*, *KLF12*, and *GLIS3* in regulating AD risk gene expression in disease-associated microglia states. Functional enrichment analysis revealed that *MITF*, *KLF12*, and *GLIS3* target genes were involved in key biological processes for microglia function such as phagocytosis, cytokine production, and cellular response (**Supplementary Fig. S6A**). Our findings that *MITF* distinctly regulates phagocytic-related pathways are in line with previous findings from *in-vitro* models (*23*). In summary, by integrating differentially expressed genes in AD with GRNs, we nominate *MITF*, *KLF12*, and *GLIS3* as potential upstream master regulators of gene expression changes relevant to AD pathogenesis.

### Regulation of phagocytosis by MITF and GPNMB

We prioritized *MITF* as a potential upstream regulator of AD-associated gene expression critical for phagocytosis and the GPNMB subtype as the myeloid phenotype linked to AD. To better understand the mechanistic relationship between them, we devised a lentiviral CRISPR activation (CRISPRa) approach to activate genes in HMC3-VPR cell lines and measured the level of phagocytosis under different substrate conditions (Fig. 3G). We first discovered that the activation of *MITF* led to increased mRNA expression of *GPNMB* detected by qPCR but not the other way around (**Supplementary Fig. S6D**), indicating that *MITF* is the upstream regulator of *GPNMB* and validating our results using the GRN inference. Furthermore, we observed that the activation of either *GPNMB* or *MITF* led to increased phagocytosis regardless of substrate types (Fig. 3H). Activating *MITF* was more effective at increasing phagocytosis except under the myelin condition. When we added a drug (ML329) that inhibits the *MITF* pathway, the phagocytosis was significantly reduced in all substrate conditions (**Supplementary Fig. S6E**). Our results demonstrate the activation of phagocytosis requires a cascade of regulatory events that involves *MITF* and *GPNMB* in AD.

### Non-cell-autonomous mechanisms affecting AD-associated microglia

To gain mechanistic insights into how different human myeloid subtypes communicate with each other and mediate AD risk through non-cell-autonomous mechanisms, we investigated the change of cell-to-cell interactions (CCIs) at different stages of AD using the `LIANA` framework (*33*) (**Supplementary Fig. S7A, Supplementary Table S14**). This approach allows us to dissect how myeloid cell signaling influences neighboring cells, potentially driving disease progression and highlighting targets for therapeutic intervention. For each individual, we inferred the magnitude, specificity, and directionality of cell-to-cell communication using gene expression profiles and known ligand-receptor interactions. We observed strong concordance between the magnitude of CCI activities from the FreshMG and the PsychAD cohorts (Fig. 4A), primarily for the homeostatic, PVM, and ADAM subtypes, whereas rare subtypes like MKI67 and CCL3 were less reproducible. By evaluating the CCI magnitude scores as a function of all 4 AD phenotypes using a linear mixed model, we identified differential CCIs associated with AD (Fig. 4B**, Supplementary Fig. S7B**), which were highly concordant across all 4 AD phenotypes (**Supplementary Fig. S7C**). We identified a total of 1,015 CCIs at FDR of 5% that were upregulated or downregulated in AD. The *APOE-SORL1* and *APOE-TREM2* interaction scores were higher in AD and were prioritized as the top AD-relevant CCIs, while *MRC1-PTPRC* interactions were down-regulated in AD. To test for genetic association, we performed the gene-set enrichment analyses on CCI pairs with increased scores in AD using GWAS data (*34*) (Fig. 4C**, Methods**). We observed AD-associated receptors had a strong association with AD risk but not with ligands. Visualizing the CCIs as directional networks in the context of different myeloid subtypes placed the GPNMB subtype as the most affected hub for CCIs that were upregulated in AD (Fig. 4D). Notably, the GPNMB subtype served as the receiving node for the *APOE-TREM2* interaction. To better understand the downstream effect of genes participating in AD-associated CCIs, we performed pathway enrichment analysis, uncovering that GPNMB-related CCIs were enriched with lipid metabolism and regulation of proteolysis (Fig. 4E).

**Figure 4.**
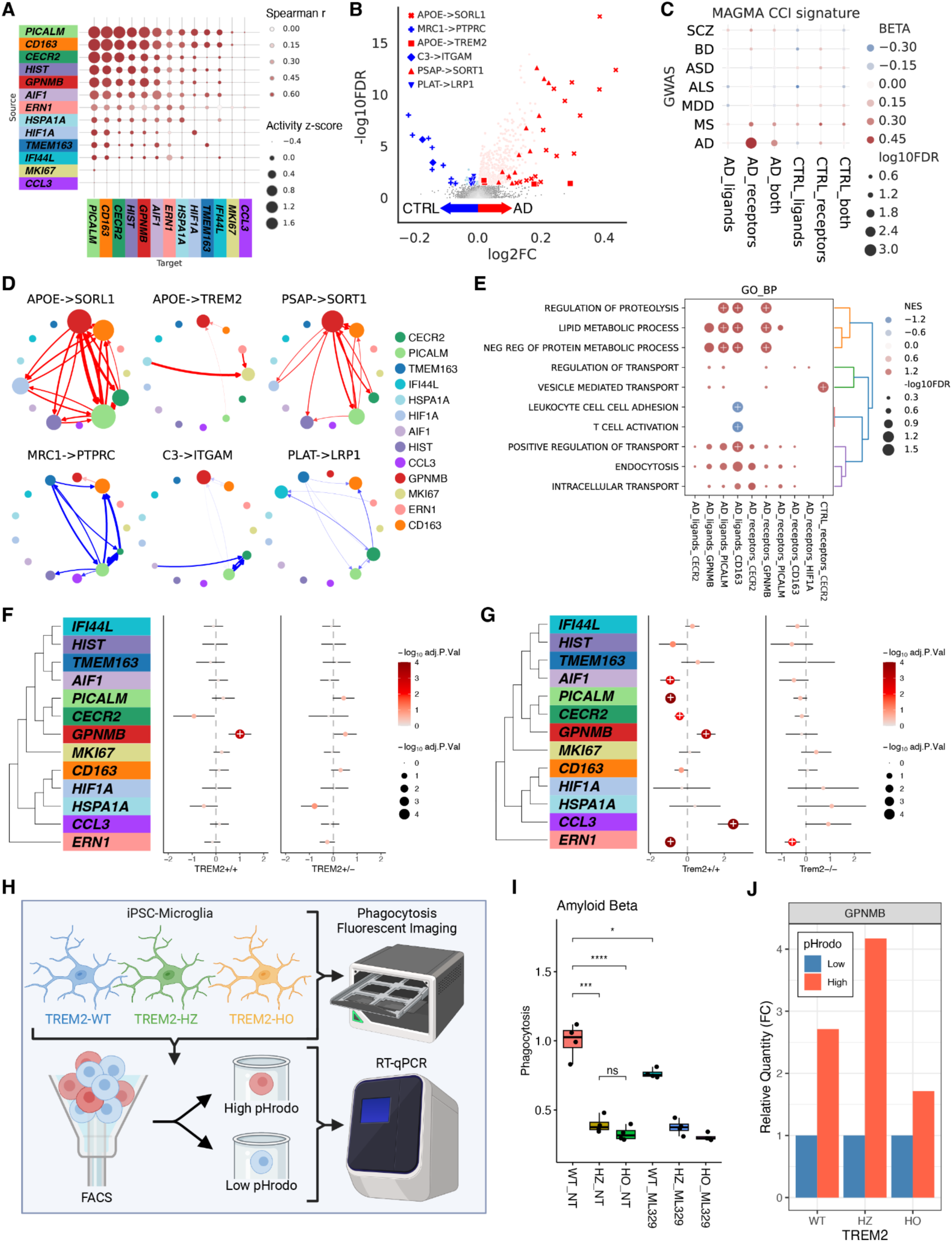
Non-cell-autonomous mechanisms. **(A)** Concordance of CCI scores among human myeloid cells between the FreshMG and the PsychAD cohorts. Pairwise Spearman correlation using aggregated CCI scores by subtype. Row labels correspond to the sender or ligand-producing cell. Column labels correspond to the receiver or receptor-producing cell. **(B)** Differential CCI analysis based on Braak stages. Meta-analysis of linear mixed model regression using both FreshMG and PsychAD cohorts. Estimated log fold change corresponds to increased representation in the high Braak stage (red) vs. the low Braak stage (blue). **(C)** `MAGMA` enrichment analysis on differential CCI, stratified by direction of regulation (AD vs CTRL) and role of interaction (ligands, receptors, or both). **(D)** Directed network visualization of the top CCI pairs. Top: AD-associated, Bottom: controls-associated CCIs. Nodes represent each subtype and directional edge weights represent the importance of interaction. The edge color represents the estimated log fold change from differential CCI analysis. **(E)** Gene set enrichment analysis of CCI pairs using Gene Ontology Biological Processes. CCIs aggregated by subtype, direction of regulation (AD vs CTRL), and role of interaction (ligands or receptors). The color scale represents the normalized enrichment score (NES). The dot size represents the FDR significance. + marks FDR < 0.05. **(F)** Compositional variation of myeloid subtypes by AD using *TREM2* missense mutation (R47H or R62H) carriers. Shared disease-free controls without TREM2 mutations were compared against AD cases with *TREM2* WT (+/+) and *TREM2* missense carriers (+/-). AD cases were sampled to match the size of *TREM2* mutation carriers. **(G)** Compositional variation of myeloid subtypes by AD using Trem2-deficient 5XFAD mice. Trem2+/+ 5XFAD and Trem2–/– 5XFAD mice were compared to disease-free control mice (Trem2+/+). **(H)** Schematic of isolating highly phagocytosing microglial cells using flow cytometry. **(I)** Relative level of phagocytosis among WT, *TREM2* heterozygous, and homozygous knockouts in iPSC-derived microglia using Aβ as substrates. **(J)** Relative mRNA expression of *GPNMB* measured by RT-qPCR for high and low phagocytosing microglia using Aβ as substrates.

### TREM2-dependent regulation of phagocytosis by AD-associated microglia

Previous studies have shown that *TREM2*, a myeloid cell receptor, plays a crucial role in the activation of disease-associated microglia, with variants increasing AD risk (*5*, *27*, *35–40*). Given the higher expression of *APOE-TREM2* CCI in the GPNMB subtype, we hypothesized that GPNMB expansion in AD is partially *TREM2*-dependent. To investigate this hypothesis, we first examined the impact of highly penetrant *TREM2* variants for AD (R47H; rs75932628; n = 21 and R62H; rs143332484; n = 26) (*39*) on changes of the microglia subtype composition. We found that carriers of these *TREM2* mutations did not exhibit an expansion of the GPNMB subtype during progression of AD (Fig. 4F), supporting a potentially protective role of this subtype in phagocytosis and the amelioration of AD pathology. To further explore this, we utilized the published snRNA-seq dataset on Trem2-deficient 5XFAD mice (*27*) (Fig.4G). Similar to the human data, in the 5XFAD mouse model, we show an increase in the proportion of the GPNMB subtype, which was absent in the Trem2-deficient 5XFAD mice.

Finally, we utilized isogenic induced pluripotent stem cell (iPSC)-derived microglia that were wild-type (WT), heterozygous (HZ), or homozygous (HO) for *TREM2*. *TREM2* knockout cells (HZ and HO) showed approximately 50% lower *GPNMB* and *MITF* mRNA expression compared to WT (**Supplementary Fig. S7D**). Phagocytosis assays using Aβ, myelin, and synaptic protein as substrates revealed significant reduction in phagocytic activity for both HZ and HO lines compared to WT (Fig. 4I**, Supplementary Fig. S7E**). Furthermore, inhibiting the *MITF* pathway (with ML329) leads to a significant reduction in Aβ phagocytosis in WT cells. We used FACS to separate microglia into high- and low-phagocytosing populations based on the fluorescence of pHrodo-labeled substrates (Fig. 4H**, Supplementary Fig. S7F**). *GPNMB* protein levels were higher in cells with high phagocytic activity (Fig. 4J). Similarly, RT-qPCR revealed that high-phagocytosing cells exhibited higher levels of *GPNMB* mRNA than low-phagocytosing cells across all substrate conditions.

## Discussion

The cell atlas presented here underscores the importance of the functional plasticity of human myeloid cells throughout life, reflecting their ability to dynamically adapt to their microenvironment. Our comprehensive analyses uncover striking similarities between normal aging and AD pathology. We speculate that the natural aging process is accelerated in AD, and follows a similar trend for all subtypes, with the exception of the AD-associated microglia, ADAM. ADAM is characterized by elevated expression of *GPNMB* transcripts and *CD44* protein. *GPNMB* is a multifaceted transmembrane protein involved in the regulation of inflammation and is implicated in several neurodegenerative diseases (*41–45*). When cleaved by proteases into its soluble form, *GPNMB* signals by binding to the *CD44* receptor to drive anti-inflammatory responses (*42*, *46*, *47*). Based on the following three main outcomes, our results collectively suggest that ADAM is involved in anti-inflammatory responses and confer neuroprotective benefits in AD.

First, ADAM shows a marked increase in prevalence with AD progression and correlates significantly with polygenic AD risk scores. It suggests that individuals with higher AD genetic predisposition may naturally exhibit increased activation of this subtype, positioning it as a potential biomarker for disease progression and reflecting an adaptive, though limited, neuroprotective response to neurodegenerative changes. This is consistent with small-scale studies supporting *GPNMB* as a cerebrospinal fluid biomarker for the early diagnosis and prognosis of AD (*48*, *49*). The significant increase in ADAM, driven by polygenic AD risk and mediated by Aβ accumulation, reveals the intricate interplay between genetic predisposition and cellular responses during AD progression. This association underscores the potential for targeted therapeutic strategies that modulate the ADAM subtype, potentially altering disease progression by mitigating the downstream effects of Aβ accumulation.

Second, we investigated cell-intrinsic factors that distinguish transcriptomic profiles between AD cases and controls. Through GRN analysis, we prioritized *MITF* as the master regulator of AD risk signatures, governing the expression of numerous AD-associated genes, including *APOE*, *DPYD*, *TREM2*, and *PTPRG* (*12*, *50*, *51*). The *MITF* network is notably enriched with markers of phagocytic activity and has been recognized as a crucial regulator of homeostatic microglial functions, particularly in promoting autophagic states and enabling microglia to migrate, detect, and clear Aβ/Tau proteinopathies (*22*, *23*, *52–54*). We confirmed that *MITF* is the upstream regulator of *GPNMB* and demonstrated that the activation of *GPNMB* is linked to increased phagocytosis. Prior work demonstrating the expression of *GPNMB* is dependent on phagocytosis of CNS-substrates (*23*), indicative of a positive feedback loop between *GPNMB* expression and phagocytosis.

Third, we examined non-cell-autonomous mechanisms that distinguish interactions and communication pathways influencing AD progression. The significant enrichment of AD genetic risk loci (*APP*, *TREM2*, *SORL1*, *SORT1*, *ABCA1*, *TSPAN14*) within the prioritized receptors of AD-associated CCIs suggests potential mechanisms behind their contribution to AD. We prioritize ADAM as a central hub in AD progression, participating in the highest number of AD-associated ligand-receptor interactions among microglia subtypes. Motivated by the AD-associated upregulation of *APOE-TREM2* ligand-receptor interactions in ADAM, we subsequently found that *TREM2* mutations diminish ADAM microglia, highlighting the dependency of this subtype on *TREM2* function. Corroborating previous observations (*23*, *35*), we demonstrate that phagocytosis is *TREM2*-dependent and regulated through *MITF*-mediated activation of *GPNMB*, reinforcing the importance of this pathway in maintaining microglial function and neuroprotection.

In conclusion, our study advances the field by providing a high-resolution view of human myeloid cell diversity and their adaptive roles in aging and AD. The identification of subtype-specific GRNs, including the *MITF-GPNMB* axis, that are *TREM2*-dependent, highlights promising therapeutic avenues for modulating microglial functions to potentially slow disease progression. Future studies should aim to validate these pathways in humanized models and explore pharmacological strategies that enhance neuroprotective myeloid subtypes, potentially altering the trajectory of AD and related diseases.

## Methods

### Sources and description of human biosamples

All brain specimens were obtained through informed consent via brain donation programs at the respective organizations. All procedures and research protocols were approved by the respective ethical committees of our collaborator’s institutions. The FreshMG samples (n = 137) were taken from 96 fresh postmortem autopsy samples obtained at the Mount Sinai/JJ Peters VA Medical Center NIH Brain and Tissue Repository (NBTR) in the Bronx, NY. An additional set of 41 fresh postmortem autopsy samples was obtained from participants in the Religious Orders Study or Rush Memory and Aging Project (ROSMAP) at Rush Alzheimer’s Disease Center (RADC) in Chicago, IL. Both studies were approved by an Institutional Review Board of Rush University Medical Center and all participants signed informed and repository consents and an Anatomic Gift Act (*55*). The PsychAD cohort comprises 1,470 donors from three brain banks, Mount Sinai NIH Brain Bank and Tissue Repository (MSSM; 1,023 samples), NIMH Human Brain Collection Core (HBCC; 295 samples), and ROSMAP (RUSH; 152 samples). Finally, LivingMG biopsies were collected from patients undergoing procedures for intracerebral hemorrhage evacuation (STUDY-18-01012A), as described previously (*9*).

### Collection and harmonization of clinical, pathological, and demographic metadata

Since the brain tissue specimens were collected from three different sites, the available clinical data varies as a function of source. As such, we used the following scheme to harmonize available clinical, pathological, and demographic metadata: the CERAD scoring scheme for neuritic plaque density (*19*) was harmonized for consistency across multiple brain banks, where the scores range from 1 to 4, with increasing CERAD number corresponding to an increase in AD burden; 1 = no neuritic plaque (normal brain), 2 = sparse (possible AD), 3 = moderate (probable AD), 4 = frequent (definite AD). Samples from ROSMAP used consensus summary diagnosis of no cognitive impairment (NCI), mild cognitive impairment (MCI), and dementia and its principal cause, Alzheimer’s dementia (*56–58*). MSSM/VA samples used clinical dementia rating (CDR), which was based on a scale of 0-5; 0 = no dementia, 0.5 = questionable dementia (very mild), 1 = mild dementia, 2 = moderate dementia, 3=severe dementia, 4 = profound dementia, 5 = terminal dementia. After consulting with clinicians, we created a harmonized ordinal variable where dementia is categorized into three levels of cognitive decline, independent of AD diagnosis; 0 = no cognitive impairment, 0.5 = MCI (mild cognitive impairment), and 1 = dementia. In addition to AD phenotype, we collected comprehensive demographic (age, sex, and genetic ancestry) and technical variables (brain bank, sequencing facility, sequence pooling information, postmortem interval (PMI; measured in minutes), APOE genotype) to describe each cohort (**Supplementary Fig. S1**, **Supplementary Table S3-4**).

### Clinical diagnosis of AD

For analysis comparing donors with AD cases and neurotypical controls, a binary clinical diagnosis variable for AD, **dx_AD**, was defined as follows. Individuals with CERAD 2, 3, or 4, Braak ≥ 3, and CDR ≥ 1 for MSSM/VA or Alzheimer’s dementia for ROSMAP were classified as AD cases. Controls were defined as individuals with CERAD 1 or 2 and Braak 0, 1, or 2.

### Measuring AD neuropathology

For analysis comparing donors with pathologic AD, the following variables were used to measure the severity of AD neuropathology. **CERAD score** (*19*). A quantitative measure of Aβ plaque density where 1 is normal, 2 is possible AD, 3 is probable AD, and 4 is definite AD (*56*). **Braak AD-staging score** measuring progression of neurofibrillary tangle neuropathology (Braak & Braak-score, or BBScore). A quantitative measure of the regional patterns of neurofibrillary tangle (NFT) density across the brain, where 0 is normal and asymptotic, 1-2 indicate initial stages where NFT begins to appear in the locus coeruleus and the transentorhinal region, 3-4 indicate progression to limbic regions, such as the hippocampus and amygdala, and 5-6 indicate NFT are widespread, affecting multiple cortical regions (*59*– *61*).

### Measuring cognitive impairment

For analysis comparing donors with AD-related dementia, the following variable was used to measure the severity of cognitive impairment. **Clinical assessment of dementia.** A harmonized variable of cognitive status based on CDR scale for MSSM/VA or NCI, MCI, Alzheimer’s dementia for ROSMAP. We used the three-level ordinal categories of clinical dementia to measure the severity of dementia, in which 0 indicates no dementia, 0.5 indicates minor cognitive impairment, and 1.0 indicates definite clinical dementia.

### Isolation and fluorescence-activated cell sorting (FACS) of microglia from fresh brain specimens (FreshMG and LivingMG)

Fresh brain tissue specimens were placed in tissue storage solution (Miltenyi Biotech, #130-100-008) and stored at 4 °C for ≤ 48hrs before processing using the Adult Brain Dissociation Kit (Miltenyi Biotech, #130-107-677), according to the manufacturer’s instructions. RNase inhibitors (Takara Bio, #2313B) were used throughout cell preparation. Following demyelination (Miltenyi Myelination removal beads - Miltenyi Biotech, #130-096-433) cells were incubated in antibody (CD45: BD Pharmingen, Clone HI30, #555483 and CD11b: BD Pharmingen, Clone ICRF44, #560914) at 1:500 for 1 hour in the dark at 4 °C with end-over-end rotation. Prior to fluorescence-activated cell sorting (FACS), DAPI (Thermoscientific, #62248) was added to facilitate the selection of viable cells. Viable (DAPI negative) CD45/CD11b positive cells were isolated by FACS using a FACSAria flow cytometer (BD Biosciences). Following FACS, cellular concentration and viability were confirmed using a Countess automated cell counter (Life technologies).

### Isolation and fluorescence-activated nuclear sorting (FANS) of nuclei from frozen brain specimens (PsychAD), with hashing

All buffers were supplemented with RNAse inhibitors (Takara, #2313B). 25 mg of frozen postmortem human brain tissue was homogenized in cold lysis buffer (0.32 M Sucrose, 5 mM CaCl2, 3 mM Magnesium acetate, 0.1 mM, EDTA, 10 mM Tris-HCl, pH8, 1 mM DTT, 0.1% Triton X-100) and filtered through a 40 µm cell strainer. The flow-through was underlaid with sucrose solution (1.8 M Sucrose, 3 mM Magnesium acetate, 1 mM DTT, 10 mM Tris-HCl, pH8) and centrifuged at 107,000 g for 1 hour at 4 °C. Pellets were resuspended in PBS supplemented with 0.5% bovine serum albumin (BSA). 6 samples were processed in parallel. Up to 2 M nuclei from each sample were pelleted at 500 g for 5 minutes at 4 °C. Nuclei were re-suspended in 100 µl staining buffer (2% BSA, 0.02% Tween-20 in PBS) and incubated with 1 µg of a unique TotalSeq-A nuclear hashing antibody (Biolegend) for 30 min at 4 °C. Prior to FANS, volumes were brought up to 250 µl with PBS and 7aad (Invitrogen, #00-6993-50) added according to the manufacturer’s instructions. 7aad positive nuclei were sorted into tubes pre-coated with 5% BSA using a FACSAria flow cytometer (BD Biosciences).

### scRNA-seq and CITE-seq library preparation (FreshMG and LivingMG)

Following FACS, 10,000 cells were processed using 10x Genomics single cell 3’ capture reagents (10x Genomics, #1000268), according to the manufacturer’s instructions. In parallel, CITE-seq was performed on a subset of samples (n = 3 donors, n = 8 replicates per donor) using the TotalSeq™-A Human Universal Cocktail (BioLegend, #399907) with 154 unique cell surface antigens, including principal lineage antigens, and includes 9 isotype control antibodies to survey surface antigens. CITE-seq was performed according to the manufacturer’s instructions. For the CITE-seq experiment, a total of 80,000 cells were loaded on 10x Genomics B chips (10,000 of each uniquely barcoded sample aliquot per B chip lane), with a total targeted recovery of around 40,000 cells.

### snRNA-seq and hashing library preparation (PsychAD)

Following FANS, nuclei were subjected to 2 washes in 200 µl staining buffer, after which they were re-suspended in 15 µl PBS and quantified (Countess II, Life Technologies). Concentrations were normalized and equal amounts of differentially hash-tagged nuclei were pooled. A total of 60,000 (10,000 each) pooled nuclei were processed using 10x Genomics single cell 3’ v3.1 reagents (10x Genomics, #1000268). Each pool was run across x2 10x Genomics lanes to create a technical replicate. At the cDNA amplification step (step 2.2) during library preparation, 1 µl 2 µm HTO cDNA PCR “additive” primer v3.1 was added (*62*). After cDNA amplification, supernatant from 0.6x SPRI selection was retained for HTO library generation. cDNA library was prepared according to the 10x Genomics protocol. HTO libraries were prepared as previously described(*62*). cDNA and HTO libraries were sequenced at NYGC using the Novaseq platform (Illumina).

### Processing of scRNA-seq data (FreshMG and LivingMG)

We developed a tracking platform to record all technical covariates (such as 10x Genomics kit lotnumber, dates of different preparations, viable cell counts, etc.) and quality metrics derived from data preprocessing. **Alignment**. Paired-end scRNA-seq reads were aligned to the hg38 reference genome and the count matrix was generated using 10x cellranger count (v7.0.0). Subsequently, we used the CellBender (*63*) to carefully separate out true cells from empty droplets with ambient RNA from raw unfiltered cellranger output. **QC**. We performed the downstream analysis by aggregating gene-count matrices of multiple samples. A battery of QC tests was performed to filter low-quality libraries and non-viable cells within each library using Pegasus (v1.7.0)(*64*). Viable cells were retained based on UMI (1,000 ≤ n_UMI ≤ 40,000), gene counts (500 ≤ n_genes ≤ 8,000), and percentage of mitochondrial reads (percent_mito ≤ 20).

We also checked for possible contamination from ambient RNA, a fraction of reads mapped to non-mRNA like rRNA, sRNA, pseudogenes, and known confounding features such as lncRNA MALAT1. Further filtering was carried out by removing doublets using the Scrublet method (*65*). After filtering, the retained count matrix was normalized and log-transformed. Batch correction. We assessed the correlation between all pairs of technical and biological variables using Canonical Correlation Analysis and used the Harmony method (*66*) to regress out unwanted confounding variables such as the source of brain tissue. Clustering. From the kNN graph calculated from the PCA, we clustered cells in the same cell state using Leiden (*67*) clustering. We use UMAP (*68*) for the visualization of resulting clusters. Cells identified as T cells, NK cells, monocytes, neutrophils, oligodendrocytes, and astrocytes were removed, and those identified as microglia and PVMs were carried forward for subsequent taxonomic analysis. **Annotation of LivingMG**. After subsetting the data for microglia and PVMs, we used myeloid taxonomy from the FreshMG dataset as reference to annotate the LivingMG dataset. We used the same set of highly variable genes from the FreshMG dataset and employed scANVI (*69*) to transfer both subclass and subtype level annotations (**Supplementary Fig. S3J**).

### Processing of snRNA-seq data (PsychAD)

**Alignment**. Samples were multiplexed by combining 6 donors in each nuclei pool using hashing, and each biosample was processed in duplicate to produce technical replicates. Paired-end snRNA-seq libraries were aligned to the hg38 reference genome using STAR solo (*70*, *71*) and multiplexed pools were demultiplexed using genotype matching via vireoSNP (*72*). After per-library count matrices were generated, the downstream processing was performed using pegasus v1.7.0 (*64*) and scanpy v1.9.1 (*73*). **QC**. We applied rigorous three-step QC to remove ambient RNA and retain nuclei for subsequent downstream analysis. First, the QC is applied at the individual nucleus level. A battery of QC tests was performed to filter low-quality nuclei within each library. Poor-quality nuclei were detected by thresholding based on UMI (1,179 ≤ n_UMI ≤ 200,000; determined based on median absolute deviation of n_UMI distribution), gene counts (986 ≤ n_genes ≤ 15,000; determined based on median absolute deviation of n_genes distribution), and percentage of mitochondrial reads (percent_mito ≤ 1).

We also checked for possible contamination from ambient RNA, the fraction of reads mapped to non-mRNA like rRNA, sRNA, and pseudogenes, as well as known confounding features, such as the lncRNA MALAT1. Second, the QC was applied at the feature level. We removed features that were not robustly expressed in at least 0.05% of nuclei. Lastly, the QC was applied at the donor level. We removed donors with very low nuclei counts, which can introduce more noise to the downstream analysis. We also removed donors with low genotype concordances. Further filtering was carried out by removing doublets using the Scrublet method (*65*). **Batch correction**. We assessed the correlation between all pairs of technical variables using Canonical Correlation Analysis and used the Harmony method (*66*) to regress out unwanted variables such as the effect of brain tissue sources. **Clustering**. Highly variable features were selected from mean and variance trends, and we used the k-nearest-neighbor (kNN) graph calculated on the basis of harmony-corrected PCA embedding space to cluster nuclei in the same cell type using Leiden (*67*) clustering algorithms. We used UMAP (*68*) for the visualization of the resulting clusters. **Isolation of myeloid cells**. Identified cell-type clusters were annotated based on manual curation of known gene marker signatures obtained from Human Cell Atlas and human DLPFC study (*74*). Classes of immune cells, including Microglia and PVM, were isolated and subjected to myeloid subtype annotation and downstream analysis.

### Processing of bulk RNA-seq data (BulkMG)

RNA was extracted from aliquots of up to 100,000 FACS-sorted CD45+ microglia using the Arcturus PicoPure RNA isolation kit (Applied Biosystems). RNA-sequencing libraries were generated using the SMARTer Stranded Total RNA-Seq Kit v2 (Takara Bio USA, #634411). Libraries were quantified by Qubit HS DNA kit (Life Technologies, #Q32851) and by quantitative PCR (KAPA Biosystems, #KK4873) before sequencing on the Hi-Seq2500 (Illumina) platform obtaining 2×100 paired-end reads.

Count matrices were generated using Kallisto pseudo-mapping (*75*) using the standard Genecode v38 reference (starting with 235,227 transcripts for 60,535 unique genes). For gene-level analyses, 21,856 features were retained for downstream analyses after filtering for features with CPM > 1 in at least 15% of samples. Correct identity of the samples was confirmed by concordance between the genetic variants obtained from RNA-seq with those obtained from ATAC-seq, or directly available genotypes, as available.

### Spatial validation using Akoya PhenoCycler

FFPE sections from both AD and control cases were used for the Akoya PhenoCycler experiment. The experiments were performed according to the manufacturer’s protocol, with the Neuroinflammation Module, Neuroscience Core Panel and Immune Module provided by Akoya. Briefly, samples were deparaffinized and hydrated. For antigen retrieval, samples were boiled in Tris-EDTA pH 9 for 20 minutes in a programmable pressure cooker. Samples were stained in Antibody Cocktail Solution containing antibodies (**Supplementary Table S5**) and PhenoCycler Blocking Buffer. Following staining, samples were washed, fixed, and loaded on the PhenoCycler, with data generated using the automatic workflow. Akoya PhenoCycler results were saved as .qpproj files. and protein expression quantified using QuPath (*76*). After the sections were annotated, cells were segmented with the QuPath extension StarDist fluorescent cell detection script, with dsb2018_paper.pb as a training model. Protein expression was quantified using raw channel intensity with spatial boundaries of cells inferred by export measurement and export detection commands using QuPath.

### Spatial transcriptomic characterization using Xenium *in situ*

**Custom panel design**. Xenium Human Brain Gene Expression Panel (10x Genomics, #1000599) and a custom panel of 100 genes (**Supplementary Table 16**) were selected for the Xenium experiment. The 100-gene custom panel consisted mainly of subclass markers selected based on specificity and gene expression level. The custom gene list was sent to 10X genomics and the probe design was performed using their in-house pipeline. **Tissue preparation**. Fresh frozen tissue specimens of DLPFC were dissected into small blocks on ice. Tissue blocks were snap frozen by submerging in an isopentane (Sigma-Aldrich, #320404-1L) bath chilled with dry ice and stored at −80 °C. Before cryosectioning, tissue blocks were allowed to equilibrate to the cryostat (Microm, #HM505) chamber temperature, and were mounted with OCT (Tissue-Tek® O.C.T. Compound, Sakura Finetek USA, #4583). After trimming, good quality 10 µm sections were flattened on the cryostat stage and placed on pre-equilibrated Xenium slides (Xenium Slides & Sample Prep Reagents, 10x Genomics, #1000460). 2-3 sections were placed on each slide. Sections were further adhered to by placing a finger on the backside of the slide for a few seconds and were then refrozen in the cryostat chamber. Slides were sealed in 50 ml tubes and stored at −80°C until Xenium sample preparation. **Sample preparation**. Xenium sample preparation was performed according to the manufacturer’s protocol; “*Xenium In Situ for Fresh Frozen Tissues – Fixation & Permeabilization, CG000581, Rev C*” and *“Xenium In Situ Gene Expression - Probe Hybridization, Ligation & Amplification, User Guide, CG000582, Rev C’’*. Briefly, fresh frozen sections mounted on Xenium slides from the previous step were removed from −80 °C storage on dry ice prior to incubation at 37 °C for 1 min. Samples were then fixed in 4% paraformaldehyde (Formaldehyde 16% in aqueous solution, VWR, #100503-917) in PBS for 30 min. After rinsing in PBS, the samples were permeabilized in 1% SDS (sodium dodecyl sulfate solution) for 2 min and then rinsed in PBS before being immersed in the pre-chilled 70% methanol and incubated for 60 min on ice. After rinsing the samples in PBS, the Xenium Cassettes were assembled on the slides. Samples were incubated with a probe hybridization mix containing both the Xenium Human Brain Gene Expression Panel (10x Genomics, #1000599) and the 100 custom gene panel at 50 °C overnight to allow the probes to hybridize to targeted mRNAs. After probe hybridization, samples were rinsed with PBST, and incubated with Xenium Post Hybridization Wash Buffer at 37 °C for 30 min. Samples were then rinsed with PBST and a ligation mix was added. Ligation was performed at 37 °C for 2 hrs to circularize the hybridized probes. After rinsing the samples with PBST, Amplification Master Mix was added to enzymatically amplify the circularized probes at 30 °C for 2 hrs. After washing with TE buffer, auto-fluorescence was quenched according to the manufacturer’s protocol and nuclei stained with DAPI prior to Xenium *in situ* analysis. Nuclear segmentation. The prepared samples were loaded into the Xenium analyzer and run according to manufacturer’s instructions “*Xenium Analyzer User Guide CG000584 Rev B*”. After the Xenium analyzer was initiated, the correct gene panel was chosen, and decoding consumables (Xenium Decoding Consumables, 10x Genomics, #1000487) and reagents (Xenium Decoding Reagents, 10x Genomics, #1000461) were loaded. The bottom of the slides was carefully cleaned with ethanol prior to loading. Once the samples were loaded and the run was initiated, the instrument scanned the whole sample area of the slides using the DAPI channel, and regions of interest were selected to maximize the capture area. Results were generated by the instrument using default settings. By default, the Xenium analyzer uses 15 µm nuclei expansion distance for segmentation of cells. To test the idea of nuclei only segmentation, we resegment the results with 0 µm nuclei expansion, by using the Xenium ranger and the following scripts:

`xeniumranger resegment --id=demo --xenium-bundle=/path/to/xenium/files--expansion-distance=0 --resegment-nuclei=True`

### Identification of myeloid cells

After generating the cell-by-gene count matrices based on nuclear segmentation, nuclei were filtered by the number of detected transcripts (n_counts ≥ 30). The count matrices from all samples were merged, log-normalized, and subjected to PCA, kNN graph calculation, and Leiden clustering. To assign major cell type labels to each cell, we combined this unsupervised clustering approach with supervised label transfer with scANVI (*69*). In short, nuclei from the RADC dataset (a subset of the full PsychAD study) with known labels for eight major CNS cell types and 27 subclass labels were used as a reference to assign labels to all cells in the unfiltered Xenium dataset. Then, we assigned labels to each Leiden cluster according to the following criteria - any cluster containing >90% of cells with a single label was assigned that label; all other clusters were removed from further analysis. In addition, cells within retained clusters were removed if their individual scANVI label did not match the label assigned to their cluster. To retain a pure microglia and PVM nuclei population, we first filtered the Xenium data for the Immune class. This population was further filtered based on the label transfer of PsychAD subclasses, to retain only nuclei with the “Microglia” and “PVM” subclass labels. This filtered data (∼24,000 nuclei) was then re-processed and normalized up to PCA computation, followed by integration with batch correction using harmony (*66*), with the batch label set as the ID of the Xenium slide (each slide contained 2-3 tissue samples), and corrected for variation in the number of detected transcripts per nucleus. The top 30 harmony-corrected PCs were then used for neighbor graph calculation, UMAP visualization, and Leiden clustering. **Taxonomy of myeloid cells**. To identify subtypes of myeloid cells in the Xenium data, we relied on the scANVI label transfer method. We used the PsychAD cohort as a reference since we expected it to be more similar to our Xenium data than the FreshMG cohort (since the PsychAD cohort was also frozen *in situ*, and contains only nuclear transcripts). Label transfer was performed for subclass (subtype) annotation as described below. However, due to the high degree of transcriptional similarity between microglia sub-populations, as well as the relative sparsity of measured informative genes, we adopted a more stringent approach and assigned microglia subclass (subtype) labels based on the stability of obtained predictions. We ran scANVI for subclass (subtype) label transfer 23 (21) independent times, differing only by a randomly generated initial condition. Then, we defined stably-predicted nuclei as those that had 17 (16) predictions in internal agreement, and assigned “unstable” labels to nuclei whose combined predictions did not fulfill this condition. Subclass predictions were subsequently refined to obtain stable subtype prediction by subsetting the dataset to Homeo (or Adapt) stably predicted nuclei and running 10 independent subtype-level predictions per subset, with predictions being accepted as stable with 8 or more consistent “votes”. **Consensus subclass and subtype labels from stability analysis.** To further strengthen the reliability of our predictions, we attempted to combine our microglia subclass and subtype prediction into a single consensus. Of note, consensus labels could be either at the subclass or subtype level, depending on the set of predicted labels obtained for a given nucleus. Our consensus voting process began by assigning pseudo-subclasses to nuclei from the subtype predictions, by asking whether at least 16 (out of 21) subtype predictions agree for a given nucleus at the subclass level (even if the subtype prediction was “unstable”). Next, cells were assigned a subclass label if the 2 subclass predictions did not contradict each other (i.e., they either agreed, or one method had a stable prediction and the other was unstable). Cells with stable subclass predictions by this methodology were then further assigned a subtype if their subtype predictions were similarly stable and consistent. Based on these criteria, ∼76% of microglia were stably assigned a subclass. Subtype annotation, however, proved more challenging, and many cells were not stably assigned to a single subtype in this analysis. **Label transfer from snRNA-seq data using scANVI.** Throughout this work, we utilize scANVI to perform reference-based label transfer as a way to assist in identifying pre-defined populations on cells. In each such instance, we performed the following steps. First, snRNA-seq gene expression data was subset to the genes also measured in our Xenium data. Next, we used the scvi-tools package (*77*, *78*) to train machine learning models for dimensionality reduction based on the reference dataset and its assigned labels (e.g., class and subclass labels from the PsychAD cohort).

Unless stated otherwise, model training was run with the following parameters. scVI models were run with 5 layers and 20 latent variables (30 for microglia sub-populations) and were trained for 50 (75) epochs; scANVI models were trained for 50 epochs with a minimal sample of 100 cells per cluster per epoch; transfer models were trained for 100 epochs. Following training, the model was applied to query Xenium data to assign labels. To assess the performance of each transfer model, we performed a “self label transfer”, predicting labels in the reference data (using the subset gene pool) and evaluated the rate of correct prediction and biases in label misassignment for each predicted category. Where appropriate, we also computed the Pearson correlation coefficients for gene expression between the reference and query data, limited to the shared subset of genes across predicted labels.

### Preparation of phagocytosis substrates

#### Myelin

Human myelin was isolated from human brain tissue using a modified protocol (*23*, *79*). Dissecting media [RPMI (Sigma, #R8758), 10% FBS (Avantor, #97068-091), 0.4 mg/mL collagenase (Sigma, #10269638001), 2 mg/mL DNAse I (Sigma, #10104159001)] was preheated in a 12-well tissue culture plate (Corning, #3513) in a 37 °C incubator. Using a scalpel, 75-125 mg of human brain white matter was minced and transferred to the preheated dissecting media and incubated at 37 °C for 30 minutes, pipetting the solution after 15 minutes. After 30 minutes, brain homogenate was transferred to a 2 mL Dounce homogenizer (Kimble, #885302) on ice, homogenized, sieved using a 40 μm cell strainer (Greiner, #542040) and transferred to a 15 mL conical tube. The homogenate was centrifuged at 400 x g for 10 minutes at 4 °C and the resulting pellet resuspended in 1.5 mL Ca²⁺/Mg²⁺-free DPBS (Gibco, #14200075). This was combined with 500 μL of fresh isotonic percoll solution [1 part Ca²⁺/Mg²⁺-free DPBS, 9 parts Percoll (Cytiva, #17-0891-02)] and mixed by pipetting. 2 mL of DPBS was gently layered on top of the Percoll-homogenate solution creating two separate layers. The solution was then centrifuged at 3000 x g for 10 minutes at 4 °C resulting in a disc of myelin between the lower and upper layers of Percoll and DPBS. The myelin was transferred to a new 1.5 mL tube and centrifuged at max speed for 10 minutes at 4 °C. The myelin pellet was washed twice with DPBS and protein concentration measured using the Pierce Rapid Gold BCA Protein Assay Kit (Pierce, #A53225). **Synaptic protein**. Human synaptic protein was isolated from fresh human brain tissue using the Syn-PER Synaptic Protein Extraction Reagent protocol (ThermoFisher, #87793). Human brain tissue was homogenized in Syn-PER Reagent and synaptic protein isolated by centrifugation. **Amyloid beta**. Amyloid-β (Aβ) was aggregated using the Beta Amyloid (1–42) Aggregation Kit (rPeptide, #A-1170-025) according to the manufacturer’s protocol. In brief, lyophilized Aβ was reconstituted and incubated at 37 °C overnight to allow for protein aggregation. After 24 hours, aggregates were collected by centrifugation and the pellet was rinsed once in 1 mL Ca²⁺/Mg²⁺-free DPBS (Gibco, #14200075) before being resuspend in 200 μL of filtered 0.1 M sodium bicarbonate (Sigma, #S5761) for pHrodo labeling. **pHrodo Labeling.** Isolated myelin, synaptic protein, and aggregated amyloid-beta was labeled using 1 μL (10.2 mM) pHrodo Red SE (Fisher, #P36600) per 1 mg of protein and incubated for 1 hour at room temperature protected from light. Labeled protein was then washed three times with DPBS before resuspending in PBS to a 100x stock concentration (1.25 mg/mL) and stored at −20 °C until further use. **Apoptotic neurons**. Apoptotic neurons were prepared using a modified protocol (*23*, *80*). SH-SY5Y neurons (ATCC, #CRL-2266) were seeded in 6-well tissue culture-treated corning plates and grown to confluence. To induce apoptosis, SH-SY5Y cells were placed inside a tissue culture biosafety cabinet without the plate cover and exposed to 60 lux of UV light for 1 min. UV lux was determined by Digital Lux Meter (Dr. Meter, # LX1010B). Neurons were harvested by pipetting with 1 mL Ca²⁺/Mg²⁺-free DPBS (Gibco, #14200075) and washed twice in PBS. Cell pellets were resuspended in 1 mL PBS supplemented with 2 μL (10.2 mM) pHrodo Red SE (Fisher, #P36600) and incubated for 15 minutes at room temperature in the dark. Labeled neurons were washed twice in PBS +20% FBS to remove any unbound pHrodo, resuspended in PBS, and counted using a Countess II FL automated cell counter (ThermoFisher). Aliquots of labeled apoptotic neurons were stored at −80 °C until used.

### Validation of GPNMB and MITF

#### HMC3 Cell Line Maintenance

The HMC3 human immortalized microglia line (ATCC, #CRL-3304) was maintained in Minimum Essential Medium (MEM) (Gibco, #11095098) supplemented with 10% heat inactivated Fetal Bovine Serum (FBS) (Avantor, #97068-091) and 100 U/mL Penicillin-Streptomycin (Gibco, #15140122). Cells were maintained in 10 cm dishes (ThermoFisher, #150466) and passaged by trypsinization (Gibco, #25200072). **Lentiviral Transduction of dCas9-VPR**. The dCas9-VPR effector system was expressed in HMC3 cells via lentiviral transduction. The lenti-EF1a-dCas9-VPR-Puro (Addgene, #99373) plasmid was packaged into lentivirus using the VectorBuilder lentiviral packaging service, and stable cell lines were generated according to the manufacturer’s instructions (https://www.addgene.org/protocols/generating-stable-cell-lines/). Parent lines expressing the dCas9-VPR system were clonalized, and dCas9 expression confirmed by qPCR. **Guide RNA Design and Preparation**. Guide RNA design and preparation protocols were adapted from the previously established protocols by Li and colleagues (*81*). Guide RNA (gRNA) sequences for MITF and GPNMB were identified by searching the CRISPR-ERA database for “gene activation” using the most up-to-date human reference genome (Human GRCh37/h19). Genes were searched indicating “U6 promoter” given our specific gRNA cloning vector. gRNA sequences with the highest efficiency and specificity (“E+S”) scores were selected and confirmed to have no off-target binding using the Cas-OFFinder database (http://www.rgenome.net/cas-offinder/). gRNA sequences were then cloned into the lentiGuide-Hygro-mTagBFP2 (Addgene, #99374) backbone using Golden Gate Assembly for digestion and ligation. Competent e. coli (NEB, #C3019I) were transformed using the gRNAs cloned into the lentiGuide-Hygro-mTagBFP2 (Addgene, #99374) plasmid vector according to the manufacturer’s protocol for heat shock transformation. Transformed bacterial colonies were grown on Ampicillin Agar (InvivoGen, #FAS-AM-S) at 37 °C overnight prior to inoculation of liquid cultures (InvivoGen, #FAS-AM-B). Plasmid DNA was isolated using the Plasmid Mini Kit (Qiagen, #12125) and subjected to sanger sequencing (Genewiz Azenta Life Sciences) to confirm the presence of gRNA sequences. **Lentivirus Preparations**. Human embryonic kidney cells (HEK293T) were used to package gRNA plasmids into lentivirus. Lentiviral production was completed using a polyethyleneimine (PEI) (Polysciences, #23966-2) transfection strategy as described by Li and colleagues (*81*). Transfections in HEK293T cells were performed in 15 cm plates (Nunc, #150468) when cells reached approximately 80% confluency. A “PEI transfection-mixture” of 110 μL PEI to 250 μL Opti-MEM (Gibco, #31985-062) was combined in a 1:1 ratio with a “DNA-mixture” containing 250 μL Opti-MEM, 8.1 μg pMDLg/pRRE (Addgene, #12251), 3.1 μg pRSV-Rev (Addgene, #12253), 4.1 μg pCMV-VSV-G (Addgene, #8454) and 12.2 μg gRNA plasmid DNA. PEI-mixture and DNA-mixtures were mixed and incubated at room-temperature for 15 minutes. After incubation, 700 μL of the mixture was added to the HEK293T cells and incubated at 37 °C for 6 hours before being replaced with 15 mL complete media. After 48 hours of incubation, viral media was collected, stored at 4 °C and fresh HEK media was replaced for an additional 24 hours incubation. Viral media was collected a second time before concentrating with Lenti-X Concentrator (Takara, #631232) according to the manufacturer protocol. Viral quantification was completed by isolating viral RNA using the NucleoSpin RNA Virus Kit (Takara, #740956), and viral copies determined using the Lenti-X qRT-PCR Titration Kit (Takara, #631235). **CRISPR activation of MITF and GPNMB**. HMC3 cells were plated in 96-well plates at a seeding density of 10,000 cells per well. After adhering overnight, cells were treated with gRNA-containing lentivirus at a final concentration of 3225 viral copies/μL (based on qPCR). Cells were transduced with virus for 24 hours before removing vial media and replacing with hygromycin (1 mg/mL) antibiotic selection media for 48 hours. **Phagocytosis**. Seventy-two hours after transduction, pHrodo-labeled substrate; myelin (5 μg/mL), amyloid-beta (5 μg/mL) or SHSY5Y Apoptotic Neurons (5000 cells/well) was added, and plates were imaged by Incucyte. Fluorescent intensity was recorded, capturing four images per well across two replicate wells. All conditions were plated in triplicate across two replicate plates and imaged after 24 hours. For RNA isolation and quantification of target genes by RT-qPCR, cells in replicate wells were harvested and pooled to increase RNA yield. **RT-qPCR**. HMC3 cells were harvested by trypsinization and RNA isolated using the Arcturus PicoPure RNA Isolation Kit (Applied Biosystems, #12204-01). RNA was quantified by NanoDrop (ThermoScientific, #840274100). RT-qPCR was performed using the Power SYBR Green RNA-to-CT 1-Step Kit (Applied Biosystems, #4389986) on the QuantStudio 5 Real-Time PCR System (Applied Biosystems, #A28135). qPCR primers were obtained from Integrated DNA Technologies (IDT), sequences for the PrimeTime qPCR Primers can be found in **Supplementary Table S16**.

### TREM2 validation using iPSC-derived microglia

#### Preparation of iPSC-derived microglia

Isogenic TREM2 wild-type (WT) (FujiFilm, #C1110), heterozygous knockout (HZ) (FujiFilm, #C1134), and homozygous knockout (HO) (FujiFilm, #C1136) lines were obtained from FujiFilm and maintained in iCell Culture media according to the manufacturer’s protocols. TREM2 WT, HZ, and HO iCell microglia were plated at 125,000 cells/mL in Poly-D-Lysine (Gibco, #A3890401) coated plates according to the manufacturer’s protocol. **Phagocytosis**. Cells were maintained in culture media for 72 hours before addition of phagocytosis substrates: amyloid-beta (5 μg/mL), myelin (5 μg/mL), synaptic protein (5 ug/mL), or apoptotic neurons (35,000/mL), with or without the MITF pathway inhibitor, ML329 (2 μM, MedChemExpress, #HY-101464)(*23*, *82*). Phagocytosis was assessed using the Sartorius Incucyte S3 Live-Cell Analysis System by imaging four fields per well. RFP was used to visualize and measure phagocytosed pHrodo-labeled substrate, while bright-field was used to estimate cell confluency. Phagocytosis was analyzed 24 hours after introduction of the substrate using the Incucyte analysis software. Statistical analyses were conducted in GraphPad Prism using two-tailed t-tests to compare phagocytosis across cell lines. **RT-qPCR**. For RT-qPCR analyses, cells were harvested by pipetting and prepared for RNA isolation using the Arcturus PicoPure RNA Isolation Kit (Applied Biosystems, #12204-01). RNA was quantified by NanoDrop (ThermoScientific, #840274100). RT-qPCR was performed on isolated RNA using the Power SYBR Green RNA-to-CT 1-Step Kit (Applied Biosystems, #4389986) on the QuantStudio 5 Real-Time PCR System (Applied Biosystems, #A28135). qPCR primers were obtained from Integrated DNA Technologies (IDT), sequences for the PrimeTime qPCR Primers can be found in **Supplementary Table S16**. **FACS**. For analysis of phagocytosis by flow cytometry, cells were harvested by pipetting before incubating with GPNMB Monoclonal antibody (Proteintech, #66926-1) at 1:1000 for 1 hour, followed by Goat anti-Mouse IgG (H+L) Cross-Adsorbed Secondary Antibody, Alexa Fluor™ 750 (ThermoFisher, #A-21037) at 1:1000 for 30 minutes. Just prior to FACS, DAPI (ThermoFisher, #62248) was added at 1:10,000. Cells were then analyzed by flow cytometry for fluorescence of pHrodo-positive phagocytosed substrate and for the presence of GPNMB protein. Cells were sorted into low and high substrate populations for RNA isolation and qPCR, as previously described.

### Compositional variation analysis

We applied `crumblr` method (https://diseaseneurogenomics.github.io/crumblr) for testing the variation of cell type composition. Analysis of count ratio data (i.e., fractions) requires special consideration since data is non-normal, heteroskedastic, and spans a low-rank space. While counts can be considered directly using Poisson, negative binomial, or Dirichlet-multinomial models for simple regression applications, these can be problematic since they 1) can be very computationally expensive, 2) can produce poorly calibrated hypothesis tests, and 3) are challenging to extend to other applications. The widely used centered log-ratio (CLR) transform from compositional data analysis makes count ratio data more normal and enables use the of linear models and other standard methods. Yet CLR-transformed data is still highly heteroskedastic: the precision of measurements varies widely. This important factor is not considered by existing methods. `crumblr` uses a fast asymptotic normal approximation of CLR-transformed counts from a Dirichlet-multinomial distribution to model the sampling variance of the transformed counts. `crumblr` enables incorporating the sampling variance as precision weights to linear (mixed) models in order to increase power and control the false positive rate. `crumblr` also uses a variance stabilizing transform (vst) based on the precision weights to improve the performance of PCA and clustering.

### Differential gene expression analysis

We applied `dreamlet` for differential expression analysis. Building from the previously developed statistical tool Dream (*83*), it applies linear mixed models to the differential expression problem in single-cell omics data. It starts by aggregating cells by the donor using a pseudobulk approach (*84*, *85*) and fits a regression model and cell. For each feature and cell cluster, the following mixed model was applied: `Gene expression ∼ Intercept + age + (1|sex) + (1|ancestry) + PMI + (1|batch) + (1|source) + phenotype,` where categorical and numerical variables were modeled as random and fixed effects, respectively. We ran gene set analysis using the full spectrum of gene-level t-statistics (*86*).

### Mediation analysis

Causal Mediation Analysis was performed using the mediation R package (*87*). From PsychAD cohort, we subsetted to 645 individuals with European ancestry who have AD phenotype variables and PRS calculations from the latest AD GWAS(*8*). For each regression, we used the following covariates:

`Age + Sex + PMI + PC1 + PC2 + PC3 + PC4 + PC5 + PC6 + PC7 + PC8 + PC9 + PC10` where PC1-10 indicate genotype PCs. For the subtype composition, we used CLR-transformed cell count fractions obtained from `crumblr` analysis. For bootstrapping, we used 10,000 simulations with 50th percentiles of the treatment variable used as the control condition and 90th percentile of the treatment variable used as the treatment condition.

### Constructing gene regulatory networks

We inferred gene regulatory networks with the pySCENIC (v 0.12.1)(*31*, *32*) pipeline using a concatenated dataset of FreshMG and PsychAD microglia cohorts. We followed the standard SCENIC expression preprocessing; log normalizing expression counts and selecting highly variable genes (3,192 total) while accounting for batch correction between datasets with scanpy (v1.9.3). The pySCENIC’s GRNboost2 (arboreto v0.1.6) method was utilized for gene regulatory network inference. The pySCENIC’s cisTarget function with Human motif database v10 (https://resources.aertslab.org/cistarget/motif2tf/motifs-v10nr_clust-nr.hgnc-m0.001-o0.0.tbl) was used to enrich for gene signatures and pruned based on *cis*-regulatory cues using default settings. The “aucell” positional argument was utilized to find the enrichment of regulons across single cells.

To compare regulon enrichment between subtypes, the resulting AUCell matrix was z-score normalized. To assess the concordance between regulon enrichment across subtypes between FreshMG and PsychAD cohorts, we computed the Pearson correlations between normalized z-scores using the pandas `corrwith` function. For each dataset, we computed the normalized regulon enrichment Z-score, and performed a meta-analysis between two datasets using Stouffer’s method.

To test whether there is a significant association between the target genes of a TF and disease signatures, we performed Fisher’s exact tests between SCENIC GRN target genes and AD risk gene signatures, based on `Dreamlet` DEG analysis. Disease-associated genes with FDR < 0.05 were selected based on four different measures of AD severity, namely, case-control diagnosis, Braak, CERAD, and dementia status. The top 3 regulons were prioritized based on the overall enrichment of the AD DEG signature. The similarity between target genes of regulons was evaluated using the Jaccard similarity index. We found the target genes of the top 3 TFs shared high similarity and clustered distinctly from other regulons (**Supplementary Fig. S6B**). Node centrality was calculated using PageRank analysis, which measures a ranking of the nodes in the graph based on the structure of the incoming links (**Supplementary Fig. S6C**). The mean estimate and -log10FDR of target risk genes in SCENIC regulons are visualized using the importance score edge weights from GRNboost2 with networkX (v3.1). For each regulon, a list of SCENIC GRN TF target genes after cisTarget pruning were obtained and, using gseapy (v1.0.5)(*88*), tested for gene-set enrichment based on the Gene Ontology Biological Processes 2023 reference (*88*). GO terms were clustered based on their Ward distance between -log10FDR values.

### Constructing a pseudotemporal trajectory of AD

We followed the same steps when constructing Braak-stage-informed pseudotimes for the FreshMG and PsychAD datasets, respectively. First, cells that were not assigned a Braak stage were excluded and the data subsetted to contain either only cells annotated as adaptive, homeostatic, ADAM, or PVM. We then computed a k-nearest-neighbor graph using Scanpy (*73*) (`scanpy.pp.neighbors`, k = 30, n_pcs = 15) on the PCA embedding, regressed with Harmony. Finally, transport maps were computed between pairs of consecutive disease stages using moscot’s TemporalProblem (*29*).

### Identification of driver genes with CellRank 2

To identify drivers of disease progression within each cell type, we first estimated a cell-cell transition matrix *T* using the *`RealTimeKernel`* implemented in the CellRank framework (*30*). Disease stage heterogeneity within each Braak stage was accounted for by including cell-cell similarity (`conn_weight=0.1`). Since moscot relies on entropical regularization to solve the underlying optimal transport problem, it returns dense transport maps, leading to negligible transition probabilities and large memory requirements when computing the transition matrix. To render our downstream analysis computationally feasible, we thresholded the constructed transition matrix using the *`RealTimeKernel`*’s automatic thresholding scheme (`threshold=”auto”`).

Based on the *`RealTimeKernel`-*derived transition matrix, we estimated terminal states with the GPCCA estimator (*30*, *89*). In the case of adaptive and homeostatic cells, we computed terminal states at the resolution of subclasses (MG_Adapt, MG_Homeo), for the PVM subset based on the subtype (PVM_CD163, ADAM_GPNMB) annotation. We confirmed the estimated terminal states as biologically plausible by quantifying the cell type and disease stage purity, respectively. Given *n* cells (n = 30 in this study) identifying a terminal state, the disease stage purity is defined as

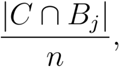

with *C* denoting the set of cells identifying the terminal state, *B_j_* the set of cells with Braak stage *j*, and |.| the cardinality of the set. Cell type purity is defined similarly.

We computed fate probabilities towards each terminal state next and identified driver genes by correlating gene expression with fate probabilities as outlined in the corresponding CellRank tutorial. To compare the gene ranking between the FreshMG and PsychAD cohorts, we subsetted to genes present in both datasets and computed the Pearson correlation coefficient between the gene-specific correlations *r_FMG_^(g)^* and *r_PAD_^(g)^* by CellRank:

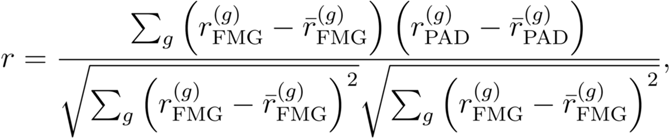

with sample mean *r^-^*.

### Pseudotime construction

DPT (*90*) is traditionally calculated on a symmetric connectivity matrix, the constructed transition matrix *T* is, however, non-symmetric. To compute DPT based on our Braak-stage-informed transition matrix, we, thus, symmetrized *T* via ½ *(T + T^T^)* and row-normalized entries. Following, we computed diffusion pseudotime using SCANPY’s `scanpy.tl.dpt` function with default values. The corresponding initial cells were identified via extreme points in diffusion components (*91*). To verify that the constructed pseudotime recapitulates our findings that homeostatic CECR2 cells decline with disease progression, while the number of homeostatic PICALM cells increases, we stratified pseudotime by subtype (Fig. 5C). Next, we performed a Welch’s t-test to validate that homeostatic CECR2 cells are assigned significantly smaller pseudotime values compared to the set of homeostatic PICALM. We performed similar Welch’s t-tests to assess the change between consecutive Braak stages. For each dataset, we first computed the T-statistics from Welch’s t-test and then combined them across datasets using Stouffer’s method: Considering T-statistics *T^freshMG^* and *T^psychAD^*, the reported T-statistic T given by

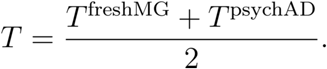

As a final analysis, we correlated fate probabilities with gene expression for each lineage to identify putative driver genes for each lineage. Here, we included genes present in both datasets. To confirm concordance between the two independent cohorts, we computed the Spearman correlation between the two rankings.

### Inferring cell-to-cell interactions

CCI analysis relies on inference of ligand-receptor interactions given a gene expression matrix with annotated cell types. By comparing to a known set of ligand-receptor (LR) pairs (reference “resource”), CCI can be inferred and scored through a variety of methods. LR pairs across cell types that are expressed above a set threshold and are differentially co-expressed, are inferred to represent interactions between the cell types. The `LIANA` (*33*) framework combines several established CCI inference methods and reference resources. We used the Python implementation of `LIANA` (v0.1.8). For each donor in FreshMG and PsychAD, we separately ran the standard rank_aggregate pipeline (resource_name = ‘consensus’, expr_prop = 0.1, min_cells = 5, n_perms = 1000). This utilizes the RobustRankAggregate (*92*) algorithm for aggregating scores from several methods (CellPhoneDB, Connectome, log2FC, NATMI, SingleCellSignalR, and CellChat)(*33*, *93–97*) into a single magnitude score for each CCI. We used the standard “consensus” CCI reference resource provided by `LIANA`. For a given pair of cell types and pair of genes to be considered for CCI scoring, each gene must be found in the reference resource, and be expressed in at least 10% of each involved cell type and in at least 5 cells. We evaluated CCI using the rank aggregate magnitude score for cell types annotated at the subtype level. We negative log-transformed these scores so that low magnitude CCI are scored close to 0 and normalized the score distribution. Additional post-processing was then performed to filter out CCI that were not expressed in both FreshMG and PsychAD.

To assess the concordance between FreshMG and PsychAD CCI scores, we first computed the average CCI score for each CCI across donors within each cohort. We then grouped the average scores by each possible pair of subtypes and measured the Spearman rank correlation. We additionally computed an activity z-score for the frequency of CCI involving each subtype pair (Fig. 4A).

To determine whether CCI are differentially expressed in AD, we applied Dream^13^ to construct a linear mixed model and account for technical and biological covariates (batch, age, sex). Linear mixed model regression was performed separately for FreshMG and PsychAD across four measures of AD progression: diagnosis (dx_AD), CERAD score, Braak stage, and dementia status. We then also used Dream to meta-analyze CCI that are significantly differentially expressed across both cohorts and to estimate the log fold change effect of each CCI for each AD progression measure (Fig. 4B**, Supplementary Fig. S7C**).

To evaluate whether significantly differentially expressed CCI are enriched in AD genome-wide association studies (GWAS), we utilize the Multi-marker Analysis of GenoMic Annotation (`MAGMA`)(*34*). The `MAGMA` gene set was constructed from significantly differentially expressed CCI LR pairs. These include CCI with at least a false discovery rate (FDR) of 0.05 significance, as determined by the Dream regression meta-analyses across each AD progression measure. We normalized the log2 fold change (log2FC) values for these CCI for each progression measure and split the CCI into AD (log2FC > 0) and CTRL (log2FC < 0) groups. Top LR pairs for AD and CTRL were then ranked by selecting the LR pairs involved in the CCI with the largest absolute normalized log2FC values. The ranked AD and CTRL LR pair gene sets were analyzed by the `MAGMA` for enrichment across several GWAS (Fig. 4C). We generated a network diagram to highlight the top-ranked three AD and CTRL CCI in our gene set (Fig. 4D).

To investigate the biological processes AD-relevant CCI are involved in, we performed a gene set enrichment analysis using the cell2cell (*98*) package and the human Gene Ontology Biological Processes 2023 reference (*99*, *100*) The AD-relevant CCI gene set was constructed in the same way as the `MAGMA` analysis gene set. We aggregated these CCI by the sender and receiver subtype to improve statistical power, and then computed normalized enrichment scores for each process based on our AD-relevant LR pair gene set. We prioritized processes that pass the 0.05 FDR significance threshold (Fig. 4E).

### Analysis of genetic heritability of AD polygenic risk

We established a standardized pipeline for Multi-marker Analysis of GenoMic Annotation (`MAGMA`) followed by single-cell Disease-Relevance Scoring (`scDRS`). `MAGMA` incorporates the association p-values of genetic variants from the latest AD genome-wide association study (GWAS)(*8*). We applied `MAGMA` using a standard window of 35 kbp upstream and 10 kbp downstream around the gene body. We executed `scDRS` using the top 1000 gene weights, sorted by Z score. The `MAGMA` and `scDRS` pipeline was run separately on FreshMG and PsychAD single-cell cohorts using the following parameters. `MAGMA` was run using `-snp-loc g1000_eur.bim` (SNP location file corresponding to the Phase 3 1000 Genome Project) and `--gene-loc NCBI38.gene.loc` (gene location file from NCBI build 38). Both files were obtained from https://ctg.cncr.nl/software/magma.

To justify the use of the top 1000 genes for `scDRS`, genes were sorted by `MAGMA` z-score and top 200, 500, and 1000 genes were used for testing the concordance between cohorts in subsequent `scDRS` analysis. `scDRS` command `scdrs.preprocess` was run with the default parameters of `n_mean_bin=20`, `n_var_bin=20w`, while `scdrs.score_cell` was run using `n_ctrl=100`. Best concordance between FreshMG and PsychAD cohorts of average `scDRS` scores (per myeloids subtype) was achieved using top 1000 genes (Pearson’s R of top n `MAGMA` genes; n = 200 was 0.85, n = 500 was 0.94, n = 1000 was 0.97, hence we proceeded with the analysis using top 1000 `MAGMA` genes).

We tested the following GWAS summary stats in `scDRS`/`MAGMA` pipeline: AD (*8*), MS (*101*), PD (*102*), MDD (*103*), ASD (*104*), BD (*105*), SCZ (*106*) and ALS (*107*). The `scDRS` scores were highly reproducible between the FreshMG and PsychAD cohorts, with AD having the greatest correlation (r = 0.97), followed by MS (r = 0.92), PD (r = 0.83), MDD (r = 0.79), ASD (r = 0.71) and ALS (r = 0.66). The lowest correlation was noted for BD (r = 0.60) and SCZ (r = 0.42), both of which are neuropsychiatric diseases. This emphasized a high reproducibility of myeloid cell heritability estimates in AD, MS, and PD, all of which are neurodegenerative diseases with progressive damage to neuronal cell types, and whose primary pathogenic mechanisms involve non-neuronal cells, including microglia and PVM. Neuropsychiatric diseases such as SCZ and BD, whose primary risks are enriched in synaptic dysfunctions of neurons, had lower correlations (**Supplementary Fig. S3C**). Per cluster association z-scores (`scDRS` assoc_mcz) were obtained using `scdrs.method.downstream_group_analysis` function. Stouffer’s method for meta-analysis was used to combine FreshMG and PsychAD `scDRS` association z-scores using the number of cells per cell cluster as cluster weights. For each cohort, `scDRS` permutation p-values per cluster were combined using Stouffer’s method on p-values with weights (cell counts of clusters). Meta p-values were further corrected for multiple testing using FDR correction (both per cell type and using all cell types combined). A more stringent correction was achieved using the per-cell type method hence we applied this correction.

### Case-control residual analysis of heritability estimates

To test for concordance in heritability estimates, we separated cases and controls and repeated `MAGMA`/`scDRS` pipe for both FreshMG and PsychAD single-cell cohorts. We calculated meta z-scores for cases and controls separately using Stouffer’s method with weights (cluster’s cell counts). We correlated meta-z-scores of cases and controls using linear regression and derived residuals as a deviation from the regression line, indicative of per-cell cluster heritability. For both cohorts, per-cell pseudotime scores were correlated with per-cell `scDRS` scores for every microglia subtype and the correlation coefficient was calculated using Spearman’s test. FreshMG and PsychAD coefficients were combined in a meta value by applying Stouffer’s method and the number of cells per cell cluster as cluster weights.

### Processing of genotypes

The FreshMG cohort genotype data consisted of samples from the Mount Sinai and Rush brain banks, as has been previously described(*9*). The PsychAD cohort genotype data consisted of samples from the Mount Sinai brain bank. For both cohorts, DNA extraction and genotyping were performed as described previously(*9*). In brief, genomic DNA was extracted from buffy coat or frozen brain tissue using the QIAamp DNA Mini Kit (Qiagen), according to the manufacturer’s instructions. Samples were genotyped using the Infinium Psych Chip Array (Illumina) at the Mount Sinai Sequencing Core. Pre-imputation processing consisted of running the quality control script HRC-1000G-check-bim.pl from the McCarthy Lab Group (https://www.well.ox.ac.uk/~wrayner/tools/), using the Trans-Omics for Precision Medicine (TOPMed) reference(*108*). Genotypes were then phased and imputed on the TOPMed Imputation Server (https://imputation.biodatacatalyst.nhlbi.nih.gov). Samples with a mismatch between one’s self-reported and genetically inferred sex, suspected sex chromosome aneuploidies, high relatedness as defined by the KING kinship coefficient (*109*) (KING > 0.177), and outlier heterozygosity (+/- 3SD from mean) were removed. Additionally, samples with a sample-level missingness > 0.05 were removed and calculated within a subset of high-quality variants (variant-level missingness ≤ 0.02).

Samples of European (EUR) ancestry, as defined by assignment to the EUR superpopulation described by the 1000 Genomes Project (*110*, *111*), were isolated using a 3SD ellipsoid method. Genotypes were first merged with GRCh38 v2a 1000 Genomes Project data (https://wellcomeopenresearch.org/articles/4-50)(111) using BCFtools version 1.9 (*112*). PLINK 2.0 (*113*) was then used to calculate the merged genotypes’ principal components (PCs), following filtering (minor allele frequency (MAF) ≥ 0.01, Hardy-Weinberg equilibrium (HWE) p-value≥1×10^−10^, variant-level missingness ≤ 0.01, regions with high linkage disequilibrium (LD) removed) and LD pruning (window size = 1000 kb, step size = 10, r2 = 0.2) steps. An ellipsoid with a radius of 3SD, corresponding to the 1000 Genomes Project EUR superpopulation, was constructed using the first three genotype PCs. Only samples that fell within this ellipsoid (FreshMG: n = 178, PsychAD: n = 759) were retained for subsequent variant-level filtering. Autosomal bi-allelic variants with an imputation R^2^ > 0.8, HWE p-value≥1×10^−6^, and variant-level missingness ≤ 0.02 were retained. Genotypes were then annotated with ancestry-specific MAF values from the National Center for Biotechnology Information’s Allele Frequency Aggregator (ALFA) (https://ftp.ncbi.nih.gov/snp/population_frequency/latest_release/). Only variants with an ancestry-specific ALFA MAF ≥ 0.01 (FreshMG: n = 10,828,658, PsychAD: n = 18,490,352) were retained.

### PRS calculation

Polygenic risk scores (PRS) were calculated on the FreshMG and PsychAD cohort samples using AD GWAS summary statistics (*8*). The PRS-CS-auto method (*114*) was used to apply continuous shrinkage priors to the effect sizes from these summary statistics. A EUR LD reference panel provided by the developers of PRS-CS was utilized (https://github.com/getian107/PRScs), which draws from 1000 Genomes Project data (*111*).

The following PRS-CS default settings were used: parameter a in the γ-γ prior = 1, parameter b in the γ-γ prior = 0.5, MCMC iterations = 1000, number of burn-in iterations = 500, and thinning of the Markov chain factor = 5. The global shrinkage parameter phi was set using a fully Bayesian determination method. Individual-level PRS were calculated using PLINK 2.0 (*113*).

### Transcriptional variation with PRS

PRS analysis was performed using the `dreamlet` package (v0.99) that applies a linear mixed model with precision weights to fit a regression model. Instead of applying a fixed contrast between two coefficients, we used AD polygenic risk scores (PRS) as a continuous variable to test for the presence of non-linear effects on variance. We subtracted admixed donors from this analysis using the PCA analysis of the first 5 genotype PCs (2 individuals were removed from PsychAD and 1 from the FreshMG cohort as clear outliers in the PCA plots). In addition, we removed a cluster of 200 individuals that were clustering separately on the PC1/PC2 plot in psychAD as admixed individuals with a percentage of a non-EUR ancestry ranging from 2.5%-10% (mainly AFR ancestry). Similarly, we removed 6 more donors from FreshMG that were outliers and showed decreased EUR and increased AFR ancestry. The removal of admixed individuals allowed us to achieve a high level of concordance in `Dreamlet` PRS analysis between FreshMG and PsychAD cohorts. For the PsychAD cohort, we considered the following covariates to model the variance: source, log(n_counts), PMI, age, AD_Bellenguez PRS, sex, dx, genotype PCs 1-5. For the FreshMG cohort, we used the same set of covariates, in addition to the sequencing batch as a covariate. We further performed a fixed-effect meta-analysis with the R metafor package using effect sizes and standard error from the `dreamlet` output (metafor `rma` function with parameters: `yi = logFC, sei = logFC / t, method = “FE”`). Meta p-values were further corrected for multiple testing using FDR correction (using the per-cell type correction).

To test whether the observed cell proportions change with PRS as a continuous variable, we applied the `crumblr` R package to each cohort and performed analysis of count ratio data with precision-weighted linear mixed models. Similar to the `Dreamlet` analysis, we modeled PRS as a continuous variable and tested for non-linear effects. FreshMG and PsychAD effect sizes were combined in a meta value using Stouffer’s method and the number of cells per cell cluster as cluster weights.

## Supplementary Information

### Assessment of *ex-vivo* activation in Microglia

Microglia are highly reactive cells and prolonged exposure to non-physiological conditions could induce unintended responses (*115*, *116*). It has previously been shown that enzymatic dissociation of brain tissue performed at elevated temperatures can stimulate microglial cells, termed *ex-vivo* activated microglia (exAM)(*17*, *79*, *117*). Failing to account for dissociation-induced changes can compromise the interpretation of microglial biology (*118*). Although artificially induced, we wondered if the proportion of exAM might reflect microglial reactivity to pathological lesions and, as such, could provide valuable insights into disease mechanisms. In this study, we are uniquely positioned to interrogate the impact of exAM as myeloids from the FreshMG cohort were isolated via enzymatic dissociation at 37°C while samples from the PsychAD cohort, consisting of flash frozen tissue, were processed using an ice-cold, enzyme free, dissociation buffer. Assessing the extent and prevalence of the exAM signature in our datasets, we observed a distinct subtype in the FreshMG dataset, called MG_exAM_ERN1, which accounts for about 14.8% of all myeloid cells, and shows up-regulation of genes implicated in cell-cell adhesion, including ERN1, PLK2 (serine/threonine-protein kinase), CSKMT, and SNHG5. Notably, PLK2 is an enzyme regulating synaptic activity and has been implicated in stimulating Aβ production (*119*, *120*). We note that the PsychAD dataset had very few cells identified as the ERN1 subtype, which belongs to the exAM subclass. This result suggests a number of possibilities. Given that the transcripts in the PsychAD dataset originate from the nuclear fraction of frozen nuclei and are free from enzymatic treatment or dissociation bias, it’s possible that the exAM cluster is predominantly derived from fresh tissue and is artificially induced during processing (*17*). In addition, the transcripts that define the ERN1 subtype may be predominantly cytoplasmic and are, thus, missing from the PsychAD samples. Moreover, we acknowledge that we observe a substantial compositional variation among three different cohorts. However, to properly assess the extent of compositional variation, we need to model this by accounting for various technical effects, including the dissection bias. In conclusion, this comparative analysis confirmed the robustness of the human microglial taxonomy, independent of tissue source, agonal state, and postmortem interval (PMI).

### Disease trajectory of human myeloid cells

Understanding the dynamic changes that take place during the onset and progression of AD at a molecular-level requires modeling of gene expression change along the measures of disease progression and identifying corresponding putative drivers. Analyses solely based on donor-level clinical covariate, such as CERAD scores or Braak staging, are limited due to the discrete nature of these variables. However, we reasoned that each labeled disease stage contains observations spanning the range of disease development and sought to identify drivers and order cells along a disease pseudotime trajectory.

Trajectory inference allows us to expand our ability to describe molecular changes at single-cell resolution. Trajectory inference methods have been remarkably successful in describing normal developmental processes faithfully and identifying regulatory mechanisms based on single-cell sequencing data (*90*, *121*, *122*). Classical pseudotime algorithms order cells along a developmental axis, with early and late cells being assigned low and high pseudotime values, respectively. To take advantage of advances in experimental design and different sources of information, alternative methods have been developed. To link observations across experimental time points, optimal transport-based solutions have been proposed to assign each measurement from one experimental time point its likely future state in the following via a probabilistic assignment in the form of transport maps (*29*, *123*). These couplings can then be used to quantify cellular change and determine the fate and putative drivers using CellRank 2’s *RealTimeKernel* (*30*). Using it together with and GPCCA estimator (*30*, *89*), we automatically inferred the terminal states of the disease dynamics. As AD onset and progression do not result in the emergence or disappearance of novel cell types, we expected to recover the major subclasses of the data, *i.e.*, adaptive, homeostatic, ADAM, or PVM. In concordance with this ground truth, we recovered the terminal states accordingly and observed a high terminal state purity defined by the fraction of cells with the correct cell type and Braak stage six (terminal state purity was 1.0 for all subclasses used). Following, we computed driver genes of these respective fates by correlating gene expression with fate probabilities (**Methods**). We replicated the same analysis using the PsychAD and observed high concordance between the correlations between gene expression and fate probabilities associated with each gene (**Supplementary Fig. S5I**). While CellRank 2 identifies putative lineage drivers, it does not align cells along the disease trajectory. To construct this orthogonal information, we used the transition matrix computed by the *RealTimeKernel* to compute a Braak-stage-informed pseudotime for the two subsets of the data in a similar fashion as previously proposed for experimental time points by the CellRank 2 study (*30*) (**Methods**). We tested if the gene expression changes over pseudotime inferred on the FreshMG cohort agrees with the corresponding change in the PsychAD dataset. As expected, the inferred change in gene expression showed high concordance between the two independent cohorts for both homeostatic and adaptive lineages (**Supplementary Figs. S5J-K**).

## Code availability

All the source codes used in this study are available via GitHub: https://github.com/DiseaseNeuroGenomics/scMyeloidAD

## Data Availability

The single-cell dataset, clinical metadata, and analysis outputs are available via the AD Knowledge Portal (https://adknowledgeportal.org). The AD Knowledge Portal is a platform for accessing data, analyses, and tools generated by the Accelerating Medicines Partnership (AMP-AD) Target Discovery Program and other National Institute on Aging (NIA)-supported programs to enable open-science practices and accelerate translational learning. The data, analyses, and tools are shared early in the research cycle without a publication embargo on secondary use. Data is available for general research use according to the following requirements for data access and data attribution (https://adknowledgeportal.org/DataAccess/Instructions). For access to data described in this manuscript see: https://www.synapse.org/#!Synapse:syn52795287.

## Acknowledgments

We would like to express our deep gratitude to the patients and their families who generously donated the invaluable biological material essential for the success of this study. We are profoundly indebted to their participation and commitment to advancing scientific knowledge and improving human health. We acknowledge the National Institute on Aging for their generous support in funding this research with the following NIH grants: R01AG065582 (PR, VH), R01AG067025 (PR, VH), and R01AG082185 (PR, VH, DL). ROSMAP is supported by P30AG10161, P30AG72975, R01AG15819, R01AG17917. U01AG46152, U01AG61356. ROSMAP resources can be requested at https://www.radc.rush.edu. We also thank Stefano Marenco and Pavan Auluck from the Human Brain Collection Core (HBCC) at the National Institute of Mental Health-Intramural Research Program for their contribution to the resource. The HBCC is supported by the NIMH-IRP under project ZIC MH002903.

## Author contributions

Conceptualization: DL, VH, JFF, PR. Methodology: DL, GEH, JFF, PR.

Investigation: JV, XW, KP, PM, EH, JMF, SPK, ZS, SA, MA, CC, AH, JFF. Formal analysis: DL, CP, CS, MP, SK, PW, RK, JB, PNM, SZ, KT, DM.

Validation: JV, XW, JMF, JFF. Resources: KGB, RS, CPK, DAB, VH.

Writing: DL, JFF, PR, with contributions from all authors. Visualization: DL, JV.

Supervision: DL, GCY, GV, FJT, GEH, VH, JFF, PR.

Project administration: DL, PR. Funding acquisition: DL, VH, PR.

All authors read and approved the final draft of the paper.

## Materials & Correspondence

Correspondence to Donghoon Lee and Panos Roussos.

## Supplementary Figures

**Supplementary Figure S1.**
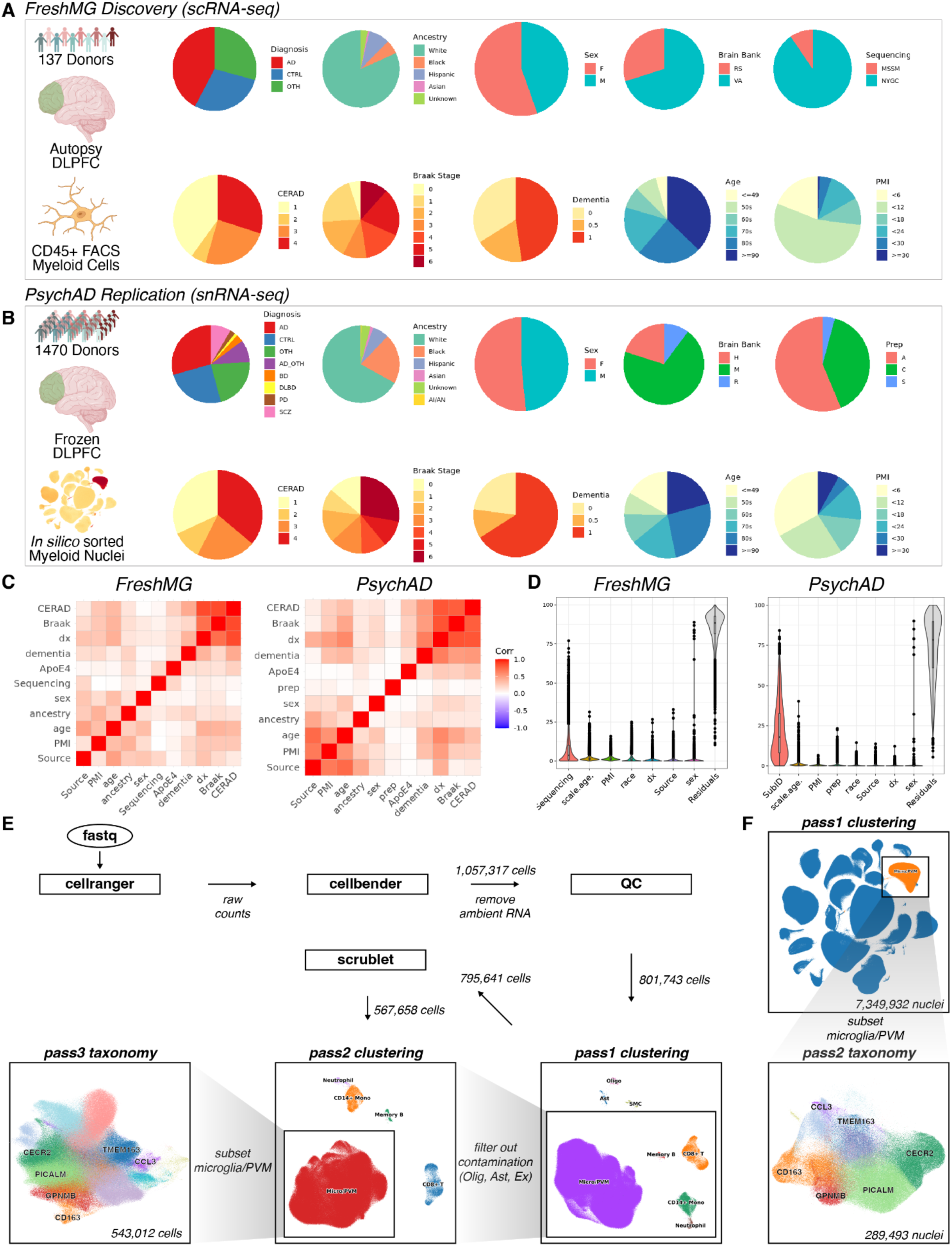
**(A)** Demographic and clinical metadata of the FreshMG cohort. **(B)** Demographic and clinical metadata of the PsychAD cohort. **(C)** Pairwise correlation of donor-level clinical variables. **(D)** Partition of gene expression variance using technical and donor-level covariates used in the study. **(E)** Schematic overview of the FreshMG single-cell data processing. **(F)** Schematic overview of the PsychAD single-cell data processing.

**Supplementary Figure S2.**
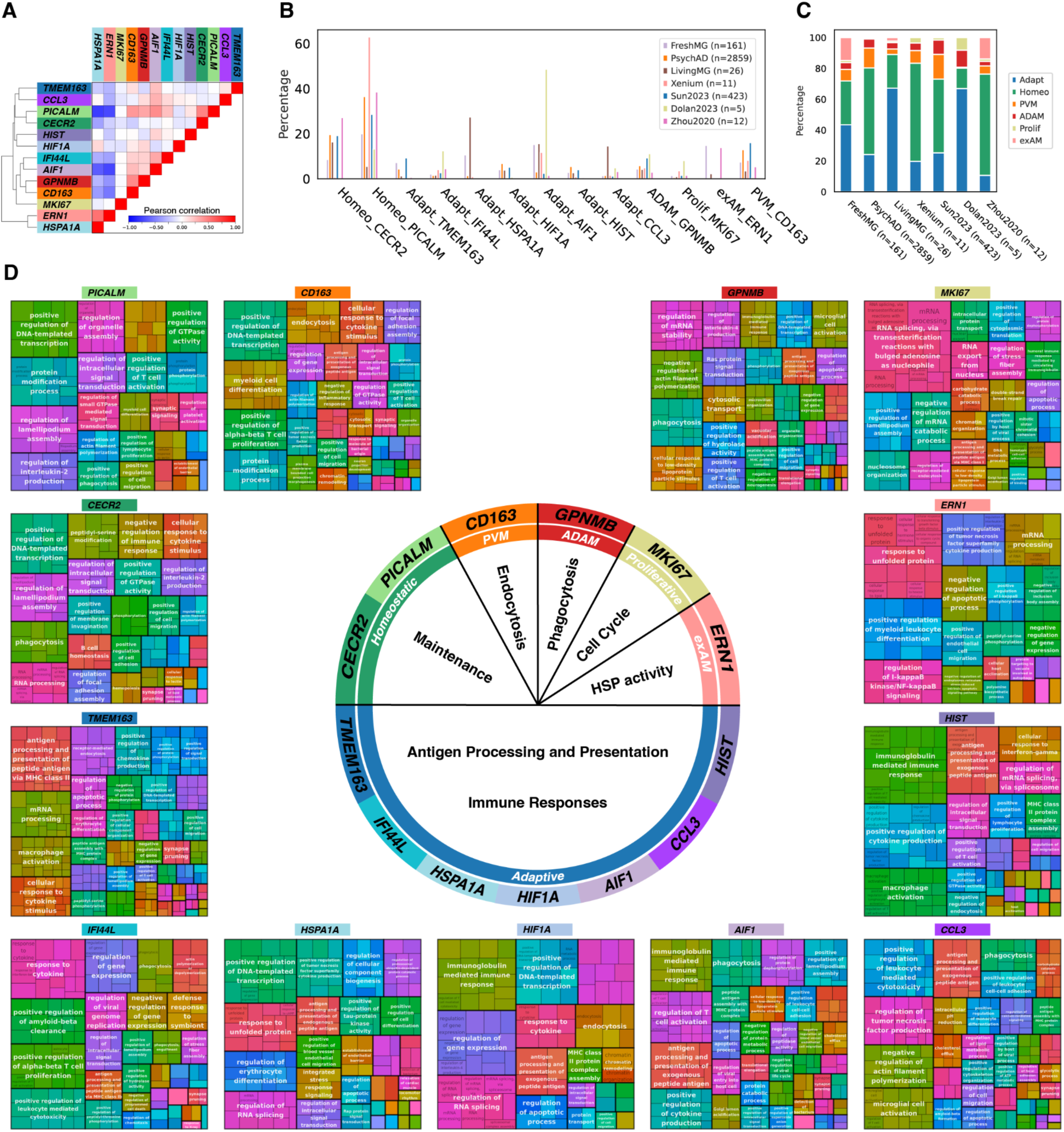
**(A)** Pairwise Pearson correlation between FreshMG and PsychAD myeloid subtypes. **(B)** Compositional differences of myeloid subtypes across independent (*LivingMG* sourced from biopsy specimens and represents living brains) and published datasets; Sun2023 (*14*), Dolan2023 (*23*), and Zhou2020 (*27*). **(C)** Compositional differences of myeloid subclasses. **(D)** Pathway enrichment analysis of human myeloid subtypes using GO biological process database. Subtype-specific genes were prioritized using Mann–Whitney U tests of one-vs-the-rest subsets.

**Supplementary Figure S3.**
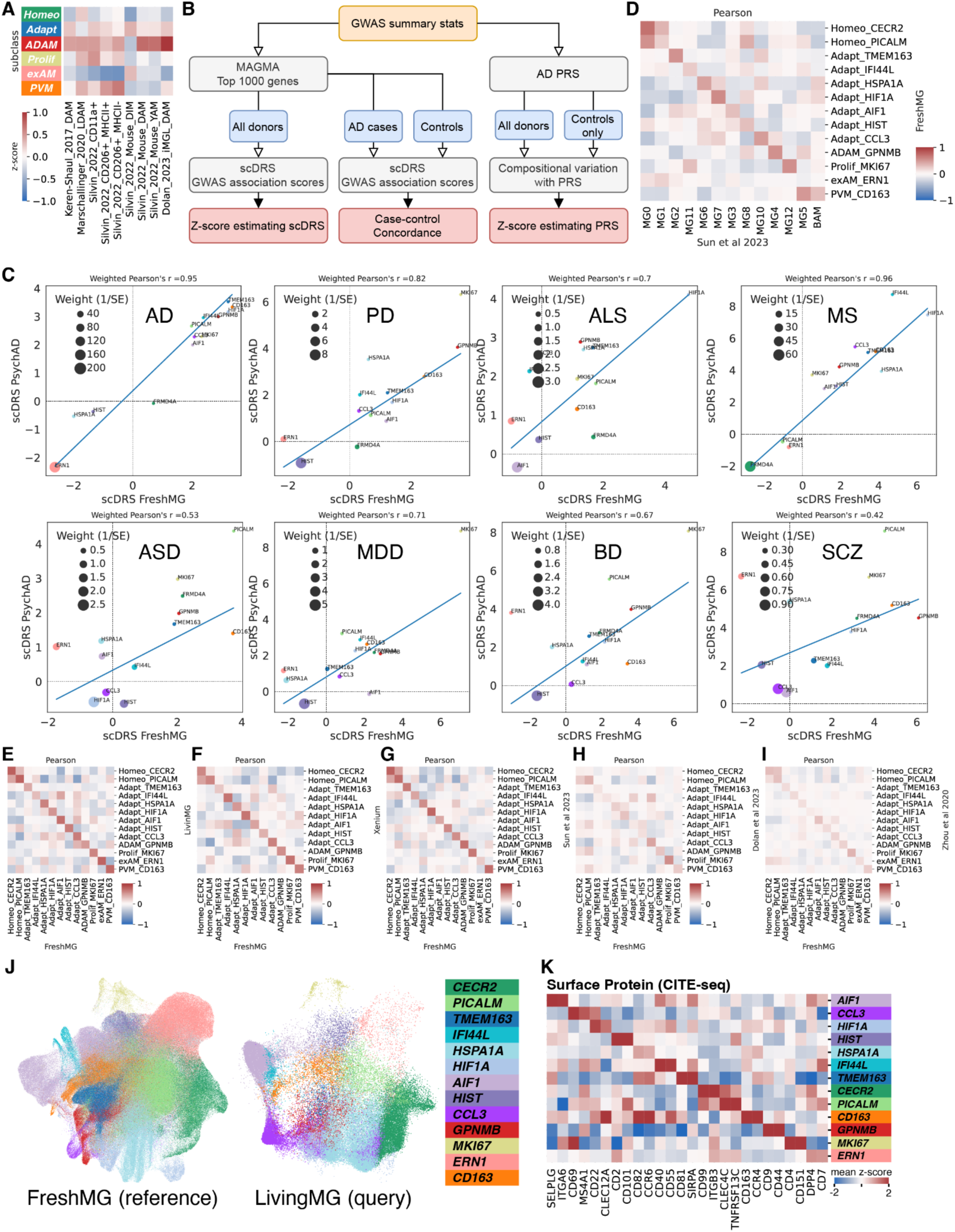
**(A)** Enrichment of gene signatures from published studies. DAM: disease-associated microglia, LDAM: lipid-droplet accumulating microglia, DIM: disease inflammatory macrophages, YAM: youth-associated microglia, iMGL: human stem-cell-differentiated microglia. **(B)** Schematic overview of the `scDRS` and PRS methods for evaluating heritability of AD risk. **(C)** Correlation of the scDRS scores between the FreshMG and PsychAD cohorts across human brain disorders. **(D)** Comparison of myeloid subtypes to microglial states defined in Sun *et al*. 2023 dataset. **(E)** Pairwise Pearson correlation between FreshMG and LivingMG myeloid subtypes. **(F)** Pairwise Pearson correlation between FreshMG and Xenium myeloid subtypes. **(G)** Pairwise Pearson correlation between FreshMG and Sun *et al*. 2023 myeloid subtypes. **(H)** Pairwise Pearson correlation between FreshMG and Dolan *et al*. 2023 myeloid subtypes. **(I)** Pairwise Pearson correlation between FreshMG and Zhou *et al*. 2020 myeloid subtypes. **(J)** Annotation of the LivingMG (biopsies) dataset using the FreshMG as the reference annotation. **(K)** Subtype-specific surface protein markers from the CITE-seq antibody derived tags (ADT).

**Supplementary Figure S4.**
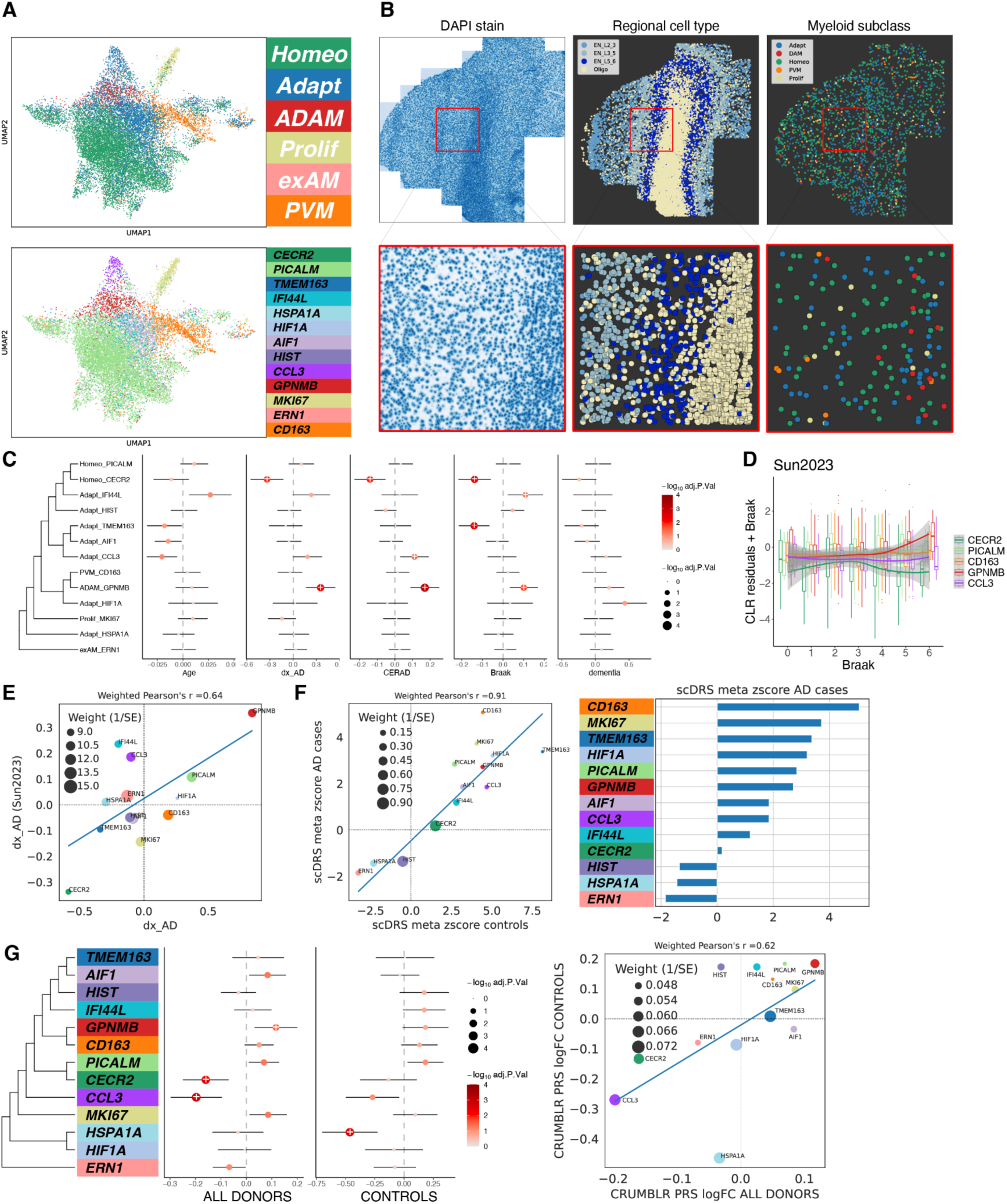
**(A)** UMAP of myeloid subclasses and subtypes from the Xenium *in situ* spatial transcriptomics dataset. **(B)** Representative slide of Xenium *in situ* spatial transcriptomics data with zoomed in region of interest. Left: DAPI stain, Middle: laminar distribution of neuronal cell types, Right: distribution of myeloid cells annotated by subclasses. **(C)** Compositional variation of aging and AD phenotypes using Sun *et al*. 2023 dataset. **(D)** Covariate adjusted compositional variation with Braak staging using Sun *et al*. 2023 dataset. **(E)** Comparison of compositional variation of dx_AD between this study and Sun *et al*. 2023. **(F)** Correlation of `scDRS` z-scores between AD cases and controls. **(G)** Compositional variation of PRS, side-by-side comparison between one using all donors and another using control donors only. Weighted Pearson’s correlation using inverse of average of standard error as weights.

**Supplementary Figure S5.**
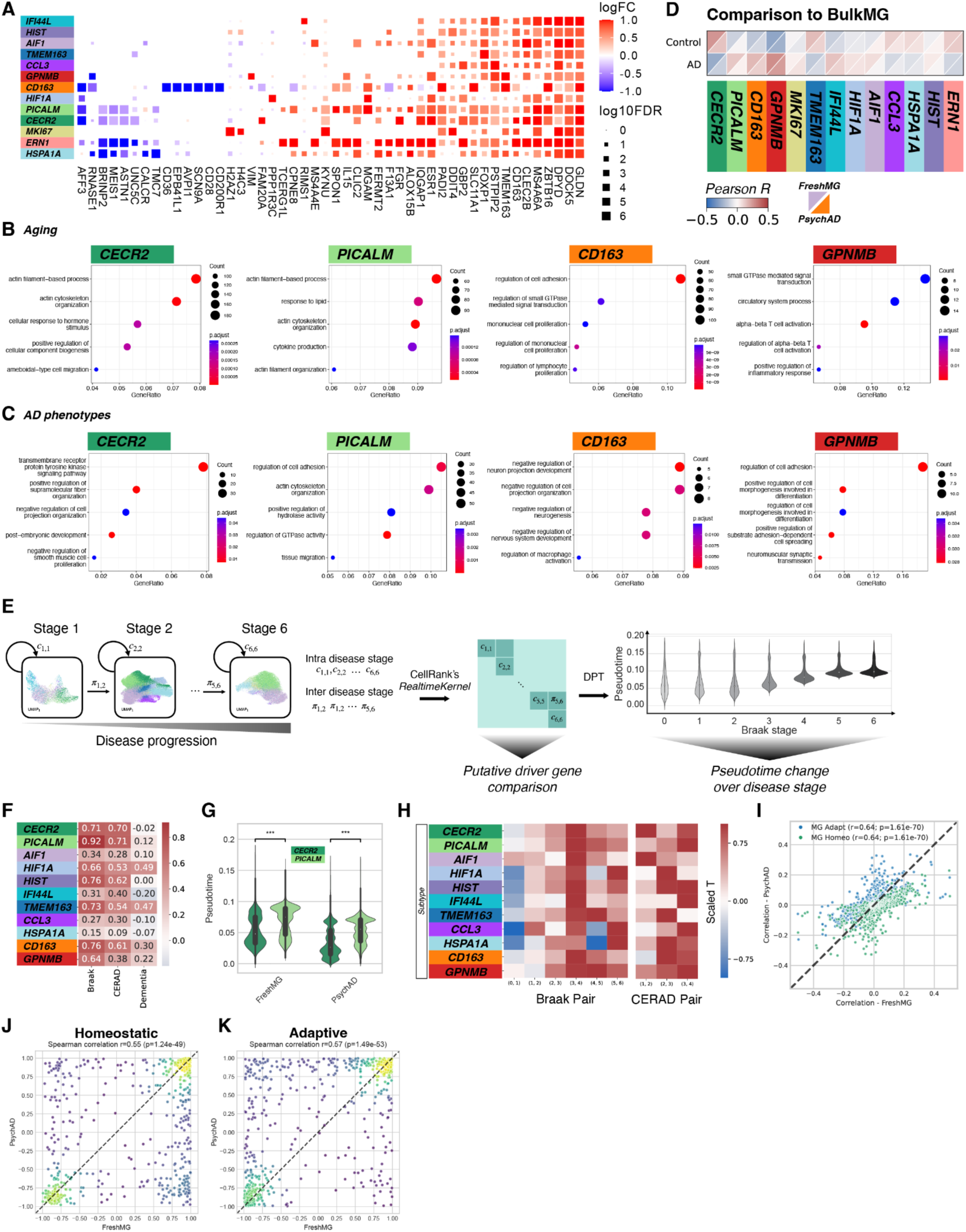
**(A)** Differentially expressed genes by myeloid subtypes associated with disease-free aging. **(B)** Pathway enrichment analysis using the GO biological process database for disease-free aging. **(C)** Pathway enrichment analysis using the GO biological process database for AD phenotypes. **(D)** Comparison of myeloid subtypes to homogenate bulk RNA-seq expression stratified by AD diagnosis. **(E)** Schematic overview of the pseudotime inference based on Braak staging. State transition matrix inferred using the optimal transport algorithm (Moscot). **(F)** Spearman correlations between inferred disease pseudotime and different measures of AD phenotypes. **(G)** Distribution of disease pseudotime between two homeostatic subtypes. **(H)** Magnitude of changes in disease pseudotime between two adjacent disease stages measured by Braak and CERAD. **(I)** Concordance of the putative drivers of two fates (adaptive and homeostatic) between FreshMG and PsychAD cohorts. Putative drivers are defined by correlating gene expression with fate probabilities. **(J)** Correlation between gene expression change and inferred disease pseudotime showing the concordance of the correlation between FreshMG and PsychAD cohorts for homeostatic and **(K)** adaptive lineages.

**Supplementary Figure S6.**
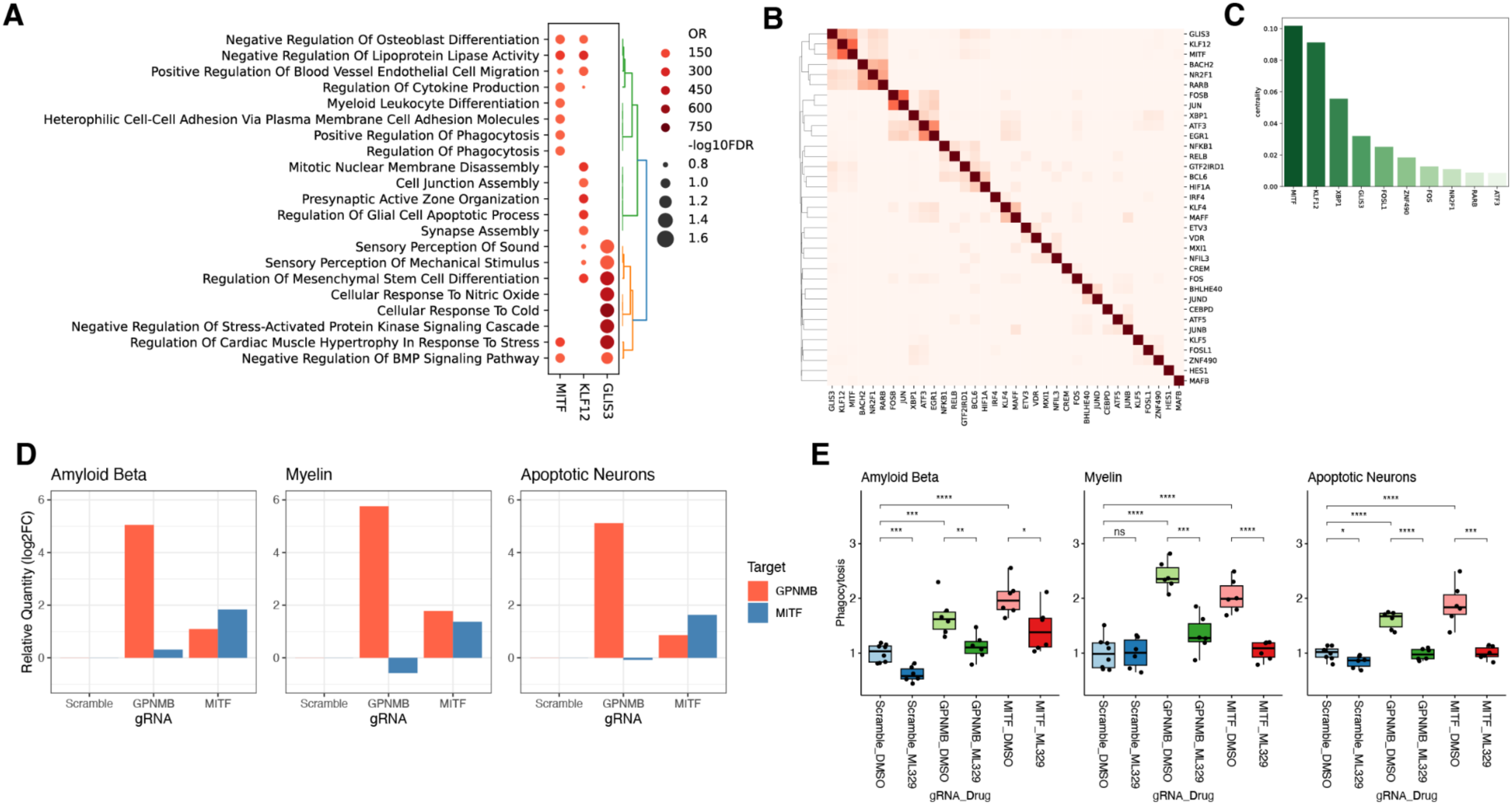
**(A)** Pathway enrichment analysis of *KLF12*, *MITF*, and *GLIS3* regulons using GO biological process database. **(B)** Jaccard Index of SCENIC regulon target genes. **(C)** PageRank centrality scores of the GRN nodes. **(D)** RT-qPCR of *GPNMB* and *MITF* after CRISPR activation in HMC3 cell line. **(E)** Relative level of phagocytosis after CRISPR activation in HMC3 cell line with or without the *MITF* pathway inhibitor ML329.

**Supplementary Figure S7.**
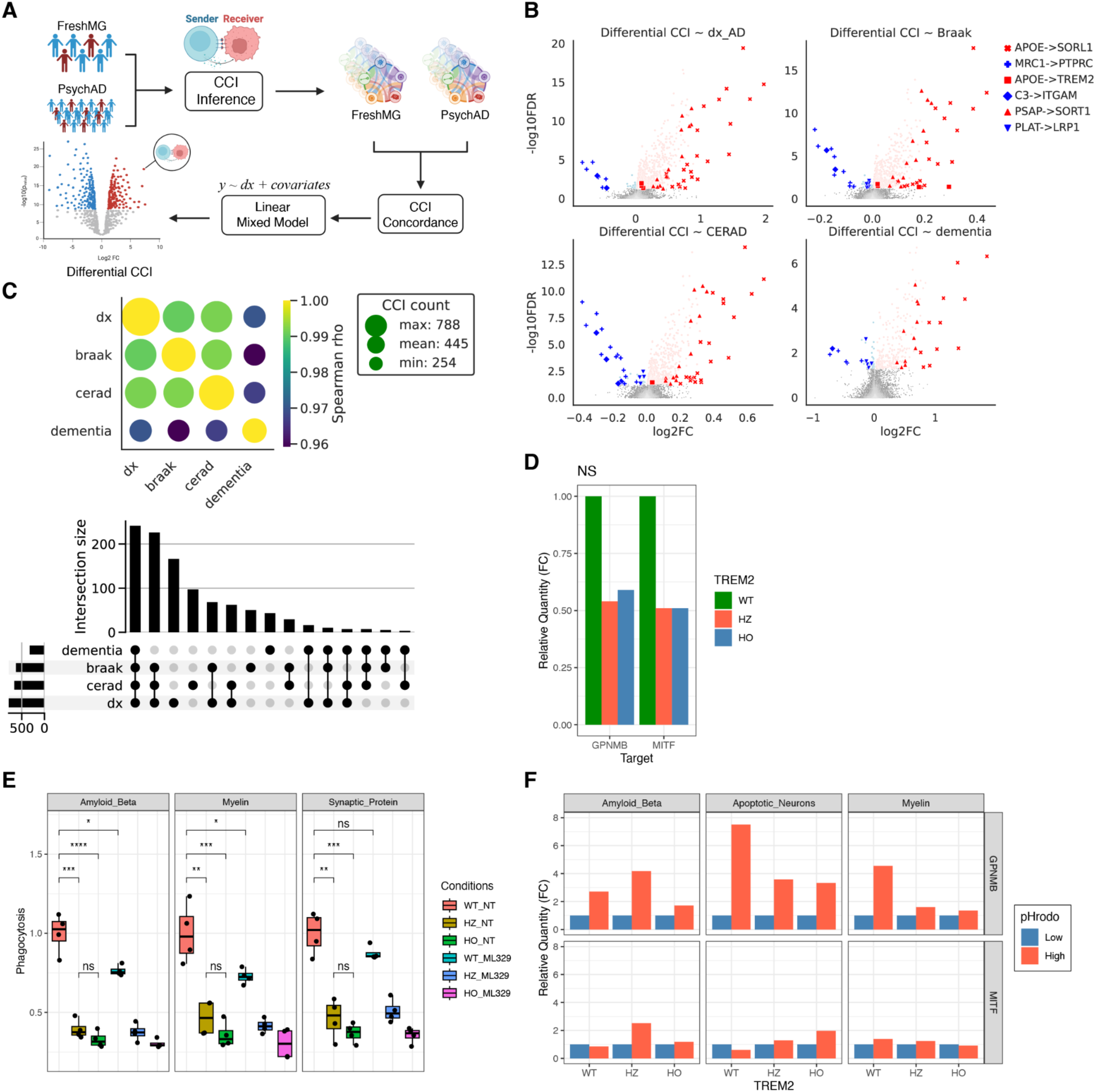
**(A)** Schematic overview of the methods for inferring disease-associated CCI. **(B)** Differential CCI using 4 different measures of AD phenotypes; AD cases vs controls (dx_AD), CERAD, Braak, and dementia. **(C)** Concordance of differential CCIs among different measures of AD phenotypes. **(D)** Relative mRNA expression of *GPNMB* and *MITF* measured by RT-qPCR for *TREM2* knockouts. **(E)** Relative level of phagocytosis among WT, TREM2 heterozygous, and homozygous knockouts in iPSC-derived microglia using Aβ, myelin, and synaptic protein as substrates. **(F)** Relative mRNA expression of *GPNMB* and *MITF* measured by RT-qPCR for high and low phagocytosing microglia using Aβ, myelin, and synaptic protein as substrates.

## Supplementary Tables

**Supplementary Table S1**. Myeloid taxonomy of the FreshMG dataset with de-identified individual ID.

**Supplementary Table S2**. Myeloid taxonomy of the PsychAD dataset with de-identified individual ID.

**Supplementary Table S3**. Clinical metadata of the FreshMG dataset with de-identified individual ID and age ranges.

**Supplementary Table S4**. Clinical metadata of the PsychAD dataset with de-identified individual ID and age ranges.

**Supplementary Table S5**. List of Akoya antibodies.

**Supplementary Table S6**. Compositional variation meta-analysis for Aging.

**Supplementary Table S7**. Compositional variation meta-analysis for AD phenotypes.

**Supplementary Table S8**. Differential gene expression meta-analysis for Aging.

**Supplementary Table S9**. Differential gene expression meta-analysis for AD phenotypes.

**Supplementary Table S10**. Pseudotime estimates for the FreshMG dataset.

**Supplementary Table S11**. Pseudotime estimates for the PsychAD dataset.

**Supplementary Table S12**. GRN adjacencies.

**Supplementary Table S13**. Regulon enrichment analysis for AD signatures.

**Supplementary Table S14**. Differential CCI meta-analysis.

**Supplementary Table S15**. scDRS scores from meta-analysis.

**Supplementary Table S16.** List of probes for the Xenium custom panel

**Supplementary Table S17**. qPCR Primers for targeted proteins

## Reference

1. J. M. Long, D. M. Holtzman, Alzheimer Disease: An Update on Pathobiology and Treatment Strategies. Cell 179, 312–339 (2019).

2. S. Hickman, S. Izzy, P. Sen, L. Morsett, J. El Khoury, Microglia in neurodegeneration. Nat. Neurosci. 21, 1359–1369 (2018).

3. F. Leng, P. Edison, Neuroinflammation and microglial activation in Alzheimer disease: where do we go from here? Nat. Rev. Neurol. 17, 157–172 (2021).

4. S. De Schepper, J. Z. Ge, G. Crowley, L. S. S. Ferreira, D. Garceau, C. E. Toomey, D. Sokolova, J. Rueda-Carrasco, S.-H. Shin, J.-S. Kim, T. Childs, T. Lashley, J. J. Burden, M. Sasner, C. Sala Frigerio, S. Jung, S. Hong, Perivascular cells induce microglial phagocytic states and synaptic engulfment via SPP1 in mouse models of Alzheimer’s disease. Nat. Neurosci. 26, 406–415 (2023).

5. H. Keren-Shaul, A. Spinrad, A. Weiner, O. Matcovitch-Natan, R. Dvir-Szternfeld, T. K. Ulland, E. David, K. Baruch, D. Lara-Astaiso, B. Toth, S. Itzkovitz, M. Colonna, M. Schwartz, I. Amit, A Unique Microglia Type Associated with Restricting Development of Alzheimer’s Disease. Cell 169, 1276–1290.e17 (2017).

6. T. Jonsson, H. Stefansson, S. Steinberg, I. Jonsdottir, P. V. Jonsson, J. Snaedal, S. Bjornsson, J. Huttenlocher, A. I. Levey, J. J. Lah, D. Rujescu, H. Hampel, I. Giegling, O. A. Andreassen, K. Engedal, I. Ulstein, S. Djurovic, C. Ibrahim-Verbaas, A. Hofman, M. A. Ikram, C. M. van Duijn, U. Thorsteinsdottir, A. Kong, K. Stefansson, Variant of TREM2 associated with the risk of Alzheimer’s disease. N. Engl. J. Med. 368, 107–116 (2013).

7. R. Guerreiro, A. Wojtas, J. Bras, M. Carrasquillo, E. Rogaeva, E. Majounie, C. Cruchaga, C. Sassi, J. S. K. Kauwe, S. Younkin, L. Hazrati, J. Collinge, J. Pocock, T. Lashley, J. Williams, J.-C. Lambert, P. Amouyel, A. Goate, R. Rademakers, K. Morgan, J. Powell, P. St George-Hyslop, A. Singleton, J. Hardy, Alzheimer Genetic Analysis Group, TREM2 variants in Alzheimer’s disease. N. Engl. J. Med. 368, 117–127 (2013).

8. C. Bellenguez, F. Küçükali, I. E. Jansen, L. Kleineidam, S. Moreno-Grau, N. Amin, A. C. Naj, R. Campos-Martin, B. Grenier-Boley, V. Andrade, P. A. Holmans, A. Boland, V. Damotte, S. J. van der Lee, M. R. Costa, T. Kuulasmaa, Q. Yang, I. de Rojas, J. C. Bis, A. Yaqub, I. Prokic, J. Chapuis, S. Ahmad, V. Giedraitis, D. Aarsland, P. Garcia-Gonzalez, C. Abdelnour, E. Alarcón-Martín, D. Alcolea, M. Alegret, I. Alvarez, V. Álvarez, N. J. Armstrong, A. Tsolaki, C. Antúnez, I. Appollonio, M. Arcaro, S. Archetti, A. A. Pastor, B. Arosio, L. Athanasiu, H. Bailly, N. Banaj, M. Baquero, S. Barral, A. Beiser, A. B. Pastor, J. E. Below, P. Benchek, L. Benussi, C. Berr, C. Besse, V. Bessi, G. Binetti, A. Bizarro, R. Blesa, M. Boada, E. Boerwinkle, B. Borroni, S. Boschi, P. Bossù, G. Bråthen, J. Bressler, C. Bresner, H. Brodaty, K. J. Brookes, L. I. Brusco, D. Buiza-Rueda, K. Bûrger, V. Burholt, W. S. Bush, M. Calero, L. B. Cantwell, G. Chene, J. Chung, M. L. Cuccaro, Á. Carracedo, R. Cecchetti, L. Cervera-Carles, C. Charbonnier, H.-H. Chen, C. Chillotti, S. Ciccone, J. A. H. R. Claassen, C. Clark, E. Conti, A. Corma-Gómez, E. Costantini, C. Custodero, D. Daian, M. C. Dalmasso, A. Daniele, E. Dardiotis, J.-F. Dartigues, P. P. de Deyn, K. de Paiva Lopes, L. D. de Witte, S. Debette, J. Deckert, T. Del Ser, N. Denning, A. DeStefano, M. Dichgans, J. Diehl-Schmid, M. Diez-Fairen, P. D. Rossi, S. Djurovic, E. Duron, E. Düzel, C. Dufouil, G. Eiriksdottir, S. Engelborghs, V. Escott-Price, A. Espinosa, M. Ewers, K. M. Faber, T. Fabrizio, S. F. Nielsen, D. W. Fardo, L. Farotti, C. Fenoglio, M. Fernández-Fuertes, R. Ferrari, C. B. Ferreira, E. Ferri, B. Fin, P. Fischer, T. Fladby, K. Fließbach, B. Fongang, M. Fornage, J. Fortea, T. M. Foroud, S. Fostinelli, N. C. Fox, E. Franco-Macías, M. J. Bullido, A. Frank-García, L. Froelich, B. Fulton-Howard, D. Galimberti, J. M. García-Alberca, P. García-González, S. Garcia-Madrona, G. Garcia-Ribas, R. Ghidoni, I. Giegling, G. Giorgio, A. M. Goate, O. Goldhardt, D. Gomez-Fonseca, A. González-Pérez, C. Graff, G. Grande, E. Green, T. Grimmer, E. Grünblatt, M. Grunin, V. Gudnason, T. Guetta-Baranes, A. Haapasalo, G. Hadjigeorgiou, J. L. Haines, K. L. Hamilton-Nelson, H. Hampel, O. Hanon, J. Hardy, A. M. Hartmann, L. Hausner, J. Harwood, S. Heilmann-Heimbach, S. Helisalmi, M. T. Heneka, I. Hernández, M. J. Herrmann, P. Hoffmann, C. Holmes, H. Holstege, R. H. Vilas, M. Hulsman, J. Humphrey, G. J. Biessels, X. Jian, C. Johansson, G. R. Jun, Y. Kastumata, J. Kauwe, P. G. Kehoe, L. Kilander, A. K. Ståhlbom, M. Kivipelto, A. Koivisto, J. Kornhuber, M. H. Kosmidis, W. A. Kukull, P. P. Kuksa, B. W. Kunkle, A. B. Kuzma, C. Lage, E. J. Laukka, L. Launer, A. Lauria, C.-Y. Lee, J. Lehtisalo, O. Lerch, A. Lleó, W. Longstreth Jr, O. Lopez, A. L. de Munain, S. Love, M. Löwemark, L. Luckcuck, K. L. Lunetta, Y. Ma, J. Macías, C. A. MacLeod, W. Maier, F. Mangialasche, M. Spallazzi, M. Marquié, R. Marshall, E. R. Martin, A. M. Montes, C. M. Rodríguez, C. Masullo, R. Mayeux, S. Mead, P. Mecocci, M. Medina, A. Meggy, S. Mehrabian, S. Mendoza, M. Menéndez-González, P. Mir, S. Moebus, M. Mol, L. Molina-Porcel, L. Montrreal, L. Morelli, F. Moreno, K. Morgan, T. Mosley, M. M. Nöthen, C. Muchnik, S. Mukherjee, B. Nacmias, T. Ngandu, G. Nicolas, B. G. Nordestgaard, R. Olaso, A. Orellana, M. Orsini, G. Ortega, A. Padovani, C. Paolo, G. Papenberg, L. Parnetti, F. Pasquier, P. Pastor, G. Peloso, A. Pérez-Cordón, J. Pérez-Tur, P. Pericard, O. Peters, Y. A. L. Pijnenburg, J. A. Pineda, G. Piñol-Ripoll, C. Pisanu, T. Polak, J. Popp, D. Posthuma, J. Priller, R. Puerta, O. Quenez, I. Quintela, J. Q. Thomassen, A. Rábano, I. Rainero, F. Rajabli, I. Ramakers, L. M. Real, M. J. T. Reinders, C. Reitz, D. Reyes-Dumeyer, P. Ridge, S. Riedel-Heller, P. Riederer, N. Roberto, E. Rodriguez-Rodriguez, A. Rongve, I. R. Allende, M. Rosende-Roca, J. L. Royo, E. Rubino, D. Rujescu, M. E. Sáez, P. Sakka, I. Saltvedt, Á. Sanabria, M. B. Sánchez-Arjona, F. Sanchez-Garcia, P. S. Juan, R. Sánchez-Valle, S. B. Sando, C. Sarnowski, C. L. Satizabal, M. Scamosci, N. Scarmeas, E. Scarpini, P. Scheltens, N. Scherbaum, M. Scherer, M. Schmid, A. Schneider, J. M. Schott, G. Selbæk, D. Seripa, M. Serrano, J. Sha, A. A. Shadrin, O. Skrobot, S. Slifer, G. J. L. Snijders, H. Soininen, V. Solfrizzi, A. Solomon, Y. Song, S. Sorbi, O. Sotolongo-Grau, G. Spalletta, A. Spottke, A. Squassina, E. Stordal, J. P. Tartan, L. Tárraga, N. Tesí, A. Thalamuthu, T. Thomas, G. Tosto, L. Traykov, L. Tremolizzo, A. Tybjærg-Hansen, A. Uitterlinden, A. Ullgren, I. Ulstein, S. Valero, O. Valladares, C. Van Broeckhoven, J. Vance, B. N. Vardarajan, A. van der Lugt, J. Van Dongen, J. van Rooij, J. van Swieten, R. Vandenberghe, F. Verhey, J.-S. Vidal, J. Vogelgsang, M. Vyhnalek, M. Wagner, D. Wallon, L.-S. Wang, R. Wang, L. Weinhold, J. Wiltfang, G. Windle, B. Woods, M. Yannakoulia, H. Zare, Y. Zhao, X. Zhang, C. Zhu, M. Zulaica, EADB, GR@ACE, DEGESCO, EADI, GERAD, Demgene, FinnGen, ADGC, CHARGE, L. A. Farrer, B. M. Psaty, M. Ghanbari, T. Raj, P. Sachdev, K. Mather, F. Jessen, M. A. Ikram, A. de Mendonça, J. Hort, M. Tsolaki, M. A. Pericak-Vance, P. Amouyel, J. Williams, R. Frikke-Schmidt, J. Clarimon, J.-F. Deleuze, G. Rossi, S. Seshadri, O. A. Andreassen, M. Ingelsson, M. Hiltunen, K. Sleegers, G. D. Schellenberg, C. M. van Duijn, R. Sims, W. M. van der Flier, A. Ruiz, A. Ramirez, J.-C. Lambert, New insights into the genetic etiology of Alzheimer’s disease and related dementias. Nat. Genet. 54, 412–436 (2022).

9. R. Kosoy, J. F. Fullard, B. Zeng, J. Bendl, P. Dong, S. Rahman, S. P. Kleopoulos, Z. Shao, K. Girdhar, J. Humphrey, K. de Paiva Lopes, A. W. Charney, B. H. Kopell, T. Raj, D. Bennett, C. P. Kellner, V. Haroutunian, G. E. Hoffman, P. Roussos, Genetics of the human microglia regulome refines Alzheimer’s disease risk loci. Nat. Genet. 54, 1145–1154 (2022).

10. Y. Chen, M. Colonna, Two-faced behavior of microglia in Alzheimer’s disease. Nat. Neurosci. 25, 3–4 (2022).

11. M. Olah, V. Menon, N. Habib, M. F. Taga, Y. Ma, C. J. Yung, M. Cimpean, A. Khairallah, G. Coronas-Samano, R. Sankowski, D. Grün, A. A. Kroshilina, D. Dionne, R. A. Sarkis, G. R. Cosgrove, J. Helgager, J. A. Golden, P. B. Pennell, M. Prinz, J. P. G. Vonsattel, A. F. Teich, J. A. Schneider, D. A. Bennett, A. Regev, W. Elyaman, E. M. Bradshaw, P. L. De Jager, Single cell RNA sequencing of human microglia uncovers a subset associated with Alzheimer’s disease. Nat. Commun. 11, 6129 (2020).

12. H. Mathys, J. Davila-Velderrain, Z. Peng, F. Gao, S. Mohammadi, J. Z. Young, M. Menon, L. He, F. Abdurrob, X. Jiang, A. J. Martorell, R. M. Ransohoff, B. P. Hafler, D. A. Bennett, M. Kellis, L.-H. Tsai, Single-cell transcriptomic analysis of Alzheimer’s disease. Nature 570, 332–337 (2019).

13. T. Masuda, R. Sankowski, O. Staszewski, C. Böttcher, L. Amann Sagar, C. Scheiwe, S. Nessler, P. Kunz, G. van Loo, V. A. Coenen, P. C. Reinacher, A. Michel, U. Sure, R. Gold, D. Grün, J. Priller, C. Stadelmann, M. Prinz, Spatial and temporal heterogeneity of mouse and human microglia at single-cell resolution. Nature 566, 388–392 (2019).

14. N. Sun, M. B. Victor, Y. P. Park, X. Xiong, A. N. Scannail, N. Leary, S. Prosper, S. Viswanathan, X. Luna, C. A. Boix, B. T. James, Y. Tanigawa, K. Galani, H. Mathys, X. Jiang, A. P. Ng, D. A. Bennett, L.-H. Tsai, M. Kellis, Human microglial state dynamics in Alzheimer’s disease progression. Cell 186, 4386–4403.e29 (2023).

15. R. C. Paolicelli, A. Sierra, B. Stevens, M.-E. Tremblay, A. Aguzzi, B. Ajami, I. Amit, E. Audinat, I. Bechmann, M. Bennett, F. Bennett, A. Bessis, K. Biber, S. Bilbo, M. Blurton-Jones, E. Boddeke, D. Brites, B. Brône, G. C. Brown, O. Butovsky, M. J. Carson, B. Castellano, M. Colonna, S. A. Cowley, C. Cunningham, D. Davalos, P. L. De Jager, B. de Strooper, A. Denes, B. J. L. Eggen, U. Eyo, E. Galea, S. Garel, F. Ginhoux, C. K. Glass, O. Gokce, D. Gomez-Nicola, B. González, S. Gordon, M. B. Graeber, A. D. Greenhalgh, P. Gressens, M. Greter, D. H. Gutmann, C. Haass, M. T. Heneka, F. L. Heppner, S. Hong, D. A. Hume, S. Jung, H. Kettenmann, J. Kipnis, R. Koyama, G. Lemke, M. Lynch, A. Majewska, M. Malcangio, T. Malm, R. Mancuso, T. Masuda, M. Matteoli, B. W. McColl, V. E. Miron, A. V. Molofsky, M. Monje, E. Mracsko, A. Nadjar, J. J. Neher, U. Neniskyte, H. Neumann, M. Noda, B. Peng, F. Peri, V. H. Perry, P. G. Popovich, C. Pridans, J. Priller, M. Prinz, D. Ragozzino, R. M. Ransohoff, M. W. Salter, A. Schaefer, D. P. Schafer, M. Schwartz, M. Simons, C. J. Smith, W. J. Streit, T. L. Tay, L.-H. Tsai, A. Verkhratsky, R. von Bernhardi, H. Wake, V. Wittamer, S. A. Wolf, L.-J. Wu, T. Wyss-Coray, Microglia states and nomenclature: A field at its crossroads. Neuron 110, 3458–3483 (2022).

16. N. Thrupp, C. Sala Frigerio, L. Wolfs, N. G. Skene, N. Fattorelli, S. Poovathingal, Y. Fourne, P. M. Matthews, T. Theys, R. Mancuso, B. de Strooper, M. Fiers, Single-Nucleus RNA-Seq Is Not Suitable for Detection of Microglial Activation Genes in Humans. Cell Rep. 32, 108189 (2020).

17. S. E. Marsh, A. J. Walker, T. Kamath, L. Dissing-Olesen, T. R. Hammond, T. Y. de Soysa, A. M. H. Young, S. Murphy, A. Abdulraouf, N. Nadaf, C. Dufort, A. C. Walker, L. E. Lucca, V. Kozareva, C. Vanderburg, S. Hong, H. Bulstrode, P. J. Hutchinson, D. J. Gaffney, D. A. Hafler, R. J. M. Franklin, E. Z. Macosko, B. Stevens, Dissection of artifactual and confounding glial signatures by single-cell sequencing of mouse and human brain. Nat. Neurosci. 25, 306–316 (2022).

18. M. Stoeckius, C. Hafemeister, W. Stephenson, B. Houck-Loomis, P. K. Chattopadhyay, H. Swerdlow, R. Satija, P. Smibert, Simultaneous epitope and transcriptome measurement in single cells. Nat. Methods 14, 865–868 (2017).

19. S. S. Mirra, A. Heyman, D. McKeel, S. M. Sumi, B. J. Crain, L. M. Brownlee, F. S. Vogel, J. P. Hughes, G. van Belle, L. Berg, The Consortium to Establish a Registry for Alzheimer’s Disease (CERAD). Part II. Standardization of the neuropathologic assessment of Alzheimer’s disease. Neurology 41, 479–486 (1991).

20. H. Braak, E. Braak, Evolution of the neuropathology of Alzheimer’s disease. Acta Neurol. Scand. Suppl. 165, 3–12 (1996).

21. A. Ajoolabady, D. Lindholm, J. Ren, D. Pratico, ER stress and UPR in Alzheimer’s disease: mechanisms, pathogenesis, treatments. Cell Death Dis. 13, 706 (2022).

22. A. M. Smith, K. Davey, S. Tsartsalis, C. Khozoie, N. Fancy, S. S. Tang, E. Liaptsi, M. Weinert, A. McGarry, R. C. J. Muirhead, S. Gentleman, D. R. Owen, P. M. Matthews, Diverse human astrocyte and microglial transcriptional responses to Alzheimer’s pathology. Acta Neuropathol. 143, 75–91 (2022).

23. M.-J. Dolan, M. Therrien, S. Jereb, T. Kamath, V. Gazestani, T. Atkeson, S. E. Marsh, A. Goeva, N. M. Lojek, S. Murphy, C. M. White, J. Joung, B. Liu, F. Limone, K. Eggan, N. Hacohen, B. E. Bernstein, C. K. Glass, V. Leinonen, M. Blurton-Jones, F. Zhang, C. B. Epstein, E. Z. Macosko, B. Stevens, Exposure of iPSC-derived human microglia to brain substrates enables the generation and manipulation of diverse transcriptional states in vitro. Nat. Immunol. 24, 1382–1390 (2023).

24. J. Marschallinger, T. Iram, M. Zardeneta, S. E. Lee, B. Lehallier, M. S. Haney, J. V. Pluvinage, V. Mathur, O. Hahn, D. W. Morgens, J. Kim, J. Tevini, T. K. Felder, H. Wolinski, C. R. Bertozzi, M. C. Bassik, L. Aigner, T. Wyss-Coray, Lipid-droplet-accumulating microglia represent a dysfunctional and proinflammatory state in the aging brain. Nat. Neurosci. 23, 194–208 (2020).

25. A. Silvin, S. Uderhardt, C. Piot, S. Da Mesquita, K. Yang, L. Geirsdottir, K. Mulder, D. Eyal, Z. Liu, C. Bridlance, M. S. Thion, X. M. Zhang, W. T. Kong, M. Deloger, V. Fontes, A. Weiner, R. Ee, R. Dress, J. W. Hang, A. Balachander, S. Chakarov, B. Malleret, G. Dunsmore, O. Cexus, J. Chen, S. Garel, C. A. Dutertre, I. Amit, J. Kipnis, F. Ginhoux, Dual ontogeny of disease-associated microglia and disease inflammatory macrophages in aging and neurodegeneration. Immunity 55, 1448–1465.e6 (2022).

26. A. C. Lieber, I. T. McNeill, J. Scaggiante, D. A. Nistal, M. Fowkes, M. Umphlett, J. Pan, P. Roussos, C. V. Mobbs, J. Mocco, C. P. Kellner, Biopsy During Minimally Invasive Intracerebral Hemorrhage Clot Evacuation. World Neurosurg., doi: 10.1016/j.wneu.2018.12.058 (2018).

27. Y. Zhou, W. M. Song, P. S. Andhey, A. Swain, T. Levy, K. R. Miller, P. L. Poliani, M. Cominelli, S. Grover, S. Gilfillan, M. Cella, T. K. Ulland, K. Zaitsev, A. Miyashita, T. Ikeuchi, M. Sainouchi, A. Kakita, D. A. Bennett, J. A. Schneider, M. R. Nichols, S. A. Beausoleil, J. D. Ulrich, D. M. Holtzman, M. N. Artyomov, M. Colonna, Human and mouse single-nucleus transcriptomics reveal TREM2-dependent and TREM2-independent cellular responses in Alzheimer’s disease. Nat. Med. 26, 131–142 (2020).

28. J. Ma, J.-T. Yu, L. Tan, MS4A Cluster in Alzheimer’s Disease. Mol. Neurobiol. 51, 1240– 1248 (2015).

29. D. Klein, G. Palla, M. Lange, M. Klein, Z. Piran, M. Gander, L. Meng-Papaxanthos, M. Sterr, A. Bastidas-Ponce, M. Tarquis-Medina, H. Lickert, M. Bakhti, M. Nitzan, M. Cuturi, F. J. Theis, Mapping cells through time and space with moscot, bioRxiv (2023)p. 2023.05.11.540374.

30. P. Weiler, M. Lange, M. Klein, D. Pe’er, F. J. Theis, Unified fate mapping in multiview single-cell data, bioRxiv (2023)p. 2023.07.19.549685.

31. S. Aibar, C. B. González-Blas, T. Moerman, V. A. Huynh-Thu, H. Imrichova, G. Hulselmans, F. Rambow, J.-C. Marine, P. Geurts, J. Aerts, J. van den Oord, Z. K. Atak, J. Wouters, S. Aerts, SCENIC: single-cell regulatory network inference and clustering. Nat. Methods 14, 1083–1086 (2017).

32. B. Van de Sande, C. Flerin, K. Davie, M. De Waegeneer, G. Hulselmans, S. Aibar, R. Seurinck, W. Saelens, R. Cannoodt, Q. Rouchon, T. Verbeiren, D. De Maeyer, J. Reumers, Y. Saeys, S. Aerts, A scalable SCENIC workflow for single-cell gene regulatory network analysis. Nat. Protoc. 15, 2247–2276 (2020).

33. D. Dimitrov, D. Türei, M. Garrido-Rodriguez, P. L. Burmedi, J. S. Nagai, C. Boys, R. O. Ramirez Flores, H. Kim, B. Szalai, I. G. Costa, A. Valdeolivas, A. Dugourd, J. Saez-Rodriguez, Comparison of methods and resources for cell-cell communication inference from single-cell RNA-Seq data. Nat. Commun. 13, 3224 (2022).

34. C. A. de Leeuw, J. M. Mooij, T. Heskes, D. Posthuma, MAGMA: generalized gene-set analysis of GWAS data. PLoS Comput. Biol. 11, e1004219 (2015).

35. Y. Wang, M. Cella, K. Mallinson, J. D. Ulrich, K. L. Young, M. L. Robinette, S. Gilfillan, G. M. Krishnan, S. Sudhakar, B. H. Zinselmeyer, D. M. Holtzman, J. R. Cirrito, M. Colonna, TREM2 lipid sensing sustains the microglial response in an Alzheimer’s disease model. Cell 160, 1061–1071 (2015).

36. T. R. Jay, C. M. Miller, P. J. Cheng, L. C. Graham, S. Bemiller, M. L. Broihier, G. Xu, D. Margevicius, J. C. Karlo, G. L. Sousa, A. C. Cotleur, O. Butovsky, L. Bekris, S. M. Staugaitis, J. B. Leverenz, S. W. Pimplikar, G. E. Landreth, G. R. Howell, R. M. Ransohoff, B. T. Lamb, TREM2 deficiency eliminates TREM2+ inflammatory macrophages and ameliorates pathology in Alzheimer’s disease mouse models. J. Exp. Med. 212, 287–295 (2015).

37. P. Yuan, C. Condello, C. D. Keene, Y. Wang, T. D. Bird, S. M. Paul, W. Luo, M. Colonna, D. Baddeley, J. Grutzendler, TREM2 haplodeficiency in mice and humans impairs the microglia barrier function leading to decreased amyloid compaction and severe axonal dystrophy. Neuron 90, 724–739 (2016).

38. Y. Wang, T. K. Ulland, J. D. Ulrich, W. Song, J. A. Tzaferis, J. T. Hole, P. Yuan, T. E. Mahan, Y. Shi, S. Gilfillan, M. Cella, J. Grutzendler, R. B. DeMattos, J. R. Cirrito, D. M. Holtzman, M. Colonna, TREM2-mediated early microglial response limits diffusion and toxicity of amyloid plaques. J. Exp. Med. 213, 667–675 (2016).

39. T. K. Ulland, M. Colonna, TREM2 - a key player in microglial biology and Alzheimer disease. Nat. Rev. Neurol. 14, 667–675 (2018).

40. W. M. Song, S. Joshita, Y. Zhou, T. K. Ulland, S. Gilfillan, M. Colonna, Humanized TREM2 mice reveal microglia-intrinsic and -extrinsic effects of R47H polymorphism. J. Exp. Med. 215, 745–760 (2018).

41. H. Tanaka, M. Shimazawa, M. Kimura, M. Takata, K. Tsuruma, M. Yamada, H. Takahashi, I. Hozumi, J.-I. Niwa, Y. Iguchi, T. Nikawa, G. Sobue, T. Inuzuka, H. Hara, The potential of GPNMB as novel neuroprotective factor in amyotrophic lateral sclerosis. Sci. Rep. 2, 573 (2012).

42. M. L. Neal, A. M. Boyle, K. M. Budge, F. F. Safadi, J. R. Richardson, The glycoprotein GPNMB attenuates astrocyte inflammatory responses through the CD44 receptor. J. Neuroinflammation 15, 73 (2018).

43. D. A. Kia, D. Zhang, S. Guelfi, C. Manzoni, L. Hubbard, R. H. Reynolds, J. Botía, M. Ryten, R. Ferrari, P. A. Lewis, N. Williams, D. Trabzuni, J. Hardy, N. W. Wood, United Kingdom Brain Expression Consortium (UKBEC) and the International Parkinson’s Disease Genomics Consortium (IPDGC), Identification of Candidate Parkinson Disease Genes by Integrating Genome-Wide Association Study, Expression, and Epigenetic Data Sets. JAMA Neurol. 78, 464–472 (2021).

44. C. S. Storm, D. A. Kia, M. M. Almramhi, S. Bandres-Ciga, C. Finan, International Parkinson’s Disease Genomics Consortium (IPDGC), A. D. Hingorani, N. W. Wood, Finding genetically-supported drug targets for Parkinson’s disease using Mendelian randomization of the druggable genome. Nat. Commun. 12, 7342 (2021).

45. M. Hüttenrauch, I. Ogorek, H. Klafki, M. Otto, C. Stadelmann, S. Weggen, J. Wiltfang, O. Wirths, Glycoprotein NMB: a novel Alzheimer’s disease associated marker expressed in a subset of activated microglia. Acta Neuropathol. Commun. 6, 108 (2018).

46. M. Liguori, E. Digifico, A. Vacchini, R. Avigni, F. S. Colombo, E. M. Borroni, F. M. Farina, S. Milanesi, A. Castagna, L. Mannarino, I. Craparotta, S. Marchini, E. Erba, N. Panini, M. Tamborini, V. Rimoldi, P. Allavena, C. Belgiovine, The soluble glycoprotein NMB (GPNMB) produced by macrophages induces cancer stemness and metastasis via CD44 and IL-33. Cell. Mol. Immunol. 18, 711–722 (2021).

47. Y. Lin, Y. Qi, M. Jiang, W. Huang, B. Li, Lactic acid-induced M2-like macrophages facilitate tumor cell migration and invasion via the GPNMB/CD44 axis in oral squamous cell carcinoma. Int. Immunopharmacol. 124, 110972 (2023).

48. J. Doroszkiewicz, A. Kulczyńska-Przybik, M. Dulewicz, R. Borawska, M. Zajkowska, A. Słowik, B. Mroczko, Potential utility of cerebrospinal fluid glycoprotein nonmetastatic melanoma protein B as a neuroinflammatory diagnostic biomarker in mild cognitive impairment and Alzheimer’s disease. J. Clin. Med. 12 (2023).

49. J. Doroszkiewicz, A. Kulczynska-Przybik, M. Dulewicz, R. Borawska, A. Slowik, B. Mroczko, The cerebrospinal fluid Glycoprotein Nonmetastatic Melanoma Protein B (GPNMB) concentration in Alzheimer’s Disease (AD). Alzheimers. Dement. 19 (2023).

50. K. Srinivasan, B. A. Friedman, A. Etxeberria, M. A. Huntley, M. P. van der Brug, O. Foreman, J. S. Paw, Z. Modrusan, T. G. Beach, G. E. Serrano, D. V. Hansen, Alzheimer’s Patient Microglia Exhibit Enhanced Aging and Unique Transcriptional Activation. Cell Rep. 31, 107843 (2020).

51. T. Schwabe, K. Srinivasan, H. Rhinn, Shifting paradigms: The central role of microglia in Alzheimer’s disease. Neurobiol. Dis. 143, 104962 (2020).

52. F. Agostini, R. Agostinis, D. L. Medina, M. Bisaglia, E. Greggio, N. Plotegher, The Regulation of MiTF/TFE Transcription Factors Across Model Organisms: from Brain Physiology to Implication for Neurodegeneration. Mol. Neurobiol. 59, 5000–5023 (2022).

53. J. Miao, H. Ma, Y. Yang, Y. Liao, C. Lin, J. Zheng, M. Yu, J. Lan, Microglia in Alzheimer’s disease: pathogenesis, mechanisms, and therapeutic potentials. Front. Aging Neurosci. 15, 1201982 (2023).

54. B. A. Friedman, K. Srinivasan, G. Ayalon, W. J. Meilandt, H. Lin, M. A. Huntley, Y. Cao, S.-H. Lee, P. C. G. Haddick, H. Ngu, Z. Modrusan, J. L. Larson, J. S. Kaminker, M. P. van der Brug, D. V. Hansen, Diverse Brain Myeloid Expression Profiles Reveal Distinct Microglial Activation States and Aspects of Alzheimer’s Disease Not Evident in Mouse Models. Cell Rep. 22, 832–847 (2018).

55. D. A. Bennett, A. S. Buchman, P. A. Boyle, L. L. Barnes, R. S. Wilson, J. A. Schneider, Religious Orders Study and Rush Memory and Aging Project. J. Alzheimers. Dis. 64, S161– S189 (2018).

56. D. A. Bennett, J. A. Schneider, Z. Arvanitakis, J. F. Kelly, N. T. Aggarwal, R. C. Shah, R. S. Wilson, Neuropathology of older persons without cognitive impairment from two community-based studies. Neurology 66, 1837–1844 (2006).

57. D. A. Bennett, R. S. Wilson, J. A. Schneider, D. A. Evans, L. A. Beckett, N. T. Aggarwal, L. L. Barnes, J. H. Fox, J. Bach, Natural history of mild cognitive impairment in older persons. Neurology 59, 198–205 (2002).

58. D. A. Bennett, J. A. Schneider, N. T. Aggarwal, Z. Arvanitakis, R. C. Shah, J. F. Kelly, J. H. Fox, E. J. Cochran, D. Arends, A. D. Treinkman, R. S. Wilson, Decision rules guiding the clinical diagnosis of Alzheimer’s disease in two community-based cohort studies compared to standard practice in a clinic-based cohort study. Neuroepidemiology 27, 169–176 (2006).

59. H. Braak, E. Braak, Neuropathological stageing of Alzheimer-related changes. Acta Neuropathol. 82, 239–259 (1991).

60. H. Braak, I. Alafuzoff, T. Arzberger, H. Kretzschmar, K. Del Tredici, Staging of Alzheimer disease-associated neurofibrillary pathology using paraffin sections and immunocytochemistry. Acta Neuropathol. 112, 389–404 (2006).

61. H. Braak, D. R. Thal, E. Ghebremedhin, K. Del Tredici, Stages of the pathologic process in Alzheimer disease: age categories from 1 to 100 years. J. Neuropathol. Exp. Neurol. 70, 960–969 (2011).

62. M. Stoeckius, S. Zheng, B. Houck-Loomis, S. Hao, B. Z. Yeung, W. M Mauck3rd, P. Smibert, R. Satija, Cell Hashing with barcoded antibodies enables multiplexing and doublet detection for single cell genomics. Genome Biol. 19, 224 (2018).

63. S. J. Fleming, M. D. Chaffin, A. Arduini, A.-D. Akkad, E. Banks, J. C. Marioni, A. A. Philippakis, P. T. Ellinor, M. Babadi, Unsupervised removal of systematic background noise from droplet-based single-cell experiments using CellBender. Nat. Methods 20, 1323–1335 (2023).

64. B. Li, J. Gould, Y. Yang, S. Sarkizova, M. Tabaka, O. Ashenberg, Y. Rosen, M. Slyper, M. S. Kowalczyk, A.-C. Villani, T. Tickle, N. Hacohen, O. Rozenblatt-Rosen, A. Regev, Cumulus provides cloud-based data analysis for large-scale single-cell and single-nucleus RNA-seq. Nat. Methods 17, 793–798 (2020).

65. S. L. Wolock, R. Lopez, A. M. Klein, Scrublet: Computational Identification of Cell Doublets in Single-Cell Transcriptomic Data. Cell Syst 8, 281–291.e9 (2019).

66. I. Korsunsky, N. Millard, J. Fan, K. Slowikowski, F. Zhang, K. Wei, Y. Baglaenko, M. Brenner, P.-R. Loh, S. Raychaudhuri, Fast, sensitive and accurate integration of single-cell data with Harmony. Nat. Methods 16, 1289–1296 (2019).

67. V. A. Traag, L. Waltman, N. J. van Eck, From Louvain to Leiden: guaranteeing well-connected communities. Sci. Rep. 9, 5233 (2019).

68. L. McInnes, J. Healy, J. Melville, UMAP: Uniform Manifold Approximation and Projection for Dimension Reduction, arXiv [stat.ML] (2018). http://arxiv.org/abs/1802.03426.

69. C. Xu, R. Lopez, E. Mehlman, J. Regier, M. I. Jordan, N. Yosef, Probabilistic harmonization and annotation of single-cell transcriptomics data with deep generative models. Mol. Syst. Biol. 17, e9620 (2021).

70. A. Dobin, C. A. Davis, F. Schlesinger, J. Drenkow, C. Zaleski, S. Jha, P. Batut, M. Chaisson, T. R. Gingeras, STAR: ultrafast universal RNA-seq aligner. Bioinformatics 29, 15–21 (2013).

71. B. Kaminow, D. Yunusov, A. Dobin, STARsolo: accurate, fast and versatile mapping/quantification of single-cell and single-nucleus RNA-seq data, bioRxiv (2021)p. 2021.05.05.442755.

72. Y. Huang, D. J. McCarthy, O. Stegle, Vireo: Bayesian demultiplexing of pooled single-cell RNA-seq data without genotype reference. Genome Biol. 20, 273 (2019).

73. F. A. Wolf, P. Angerer, F. J. Theis, SCANPY: large-scale single-cell gene expression data analysis. Genome Biol. 19, 15 (2018).

74. S. Ma, M. Skarica, Q. Li, C. Xu, R. D. Risgaard, A. T. N. Tebbenkamp, X. Mato-Blanco, R. Kovner, Ž. Krsnik, X. de Martin, V. Luria, X. Martí-Pérez, D. Liang, A. Karger, D. K. Schmidt, Z. Gomez-Sanchez, C. Qi, K. T. Gobeske, S. Pochareddy, A. Debnath, C. J. Hottman, J. Spurrier, L. Teo, A. G. Boghdadi, J. Homman-Ludiye, J. J. Ely, E. W. Daadi, D. Mi, M. Daadi, O. Marín, P. R. Hof, M.-R. Rasin, J. Bourne, C. C. Sherwood, G. Santpere, M. J. Girgenti, S. M. Strittmatter, A. M. M. Sousa, N. Sestan, Molecular and cellular evolution of the primate dorsolateral prefrontal cortex. Science 377, eabo7257 (2022).

75. N. Bray, H. Pimentel, P. Melsted, L. Pachter, Near-optimal RNA-Seq quantification with kallisto. Nat. Biotechnol.

76. P. Bankhead, M. B. Loughrey, J. A. Fernández, Y. Dombrowski, D. G. McArt, P. D. Dunne, S. McQuaid, R. T. Gray, L. J. Murray, H. G. Coleman, J. A. James, M. Salto-Tellez, P. W. Hamilton, QuPath: Open source software for digital pathology image analysis. Sci. Rep. 7, 16878 (2017).

77. A. Gayoso, R. Lopez, G. Xing, P. Boyeau, V. Valiollah Pour Amiri, J. Hong, K. Wu, M. Jayasuriya, E. Mehlman, M. Langevin, Y. Liu, J. Samaran, G. Misrachi, A. Nazaret, O. Clivio, C. Xu, T. Ashuach, M. Gabitto, M. Lotfollahi, V. Svensson, E. da Veiga Beltrame, V. Kleshchevnikov, C. Talavera-López, L. Pachter, F. J. Theis, A. Streets, M. I. Jordan, J. Regier, N. Yosef, A Python library for probabilistic analysis of single-cell omics data. Nat. Biotechnol. 40, 163–166 (2022).

78. I. Virshup, D. Bredikhin, L. Heumos, G. Palla, G. Sturm, A. Gayoso, I. Kats, M. Koutrouli, Scverse Community, B. Berger, D. Pe’er, A. Regev, S. A. Teichmann, F. Finotello, F. A. Wolf, N. Yosef, O. Stegle, F. J. Theis, The scverse project provides a computational ecosystem for single-cell omics data analysis. Nat. Biotechnol. 41, 604–606 (2023).

79. D. Mattei, A. Ivanov, M. van Oostrum, S. Pantelyushin, J. Richetto, F. Mueller, M. Beffinger, L. Schellhammer, J. Vom Berg, B. Wollscheid, D. Beule, R. C. Paolicelli, U. Meyer, Enzymatic Dissociation Induces Transcriptional and Proteotype Bias in Brain Cell Populations. Int. J. Mol. Sci. 21 (2020).

80. S. Krasemann, C. Madore, R. Cialic, C. Baufeld, N. Calcagno, R. El Fatimy, L. Beckers, E. O’Loughlin, Y. Xu, Z. Fanek, D. J. Greco, S. T. Smith, G. Tweet, Z. Humulock, T. Zrzavy, P. Conde-Sanroman, M. Gacias, Z. Weng, H. Chen, E. Tjon, F. Mazaheri, K. Hartmann, A. Madi, J. D. Ulrich, M. Glatzel, A. Worthmann, J. Heeren, B. Budnik, C. Lemere, T. Ikezu, F. L. Heppner, V. Litvak, D. M. Holtzman, H. Lassmann, H. L. Weiner, J. Ochando, C. Haass, O. Butovsky, The TREM2-APOE pathway drives the transcriptional phenotype of dysfunctional microglia in neurodegenerative diseases. Immunity 47, 566–581.e9 (2017).

81. Cell Press: STAR Protocols. https://star-protocols.cell.com/protocols/694.

82. L. Li, X. Cao, L. Huang, X. Huang, J. Gu, X. Yu, Y. Zhu, Y. Zhou, Y. Song, M. Zhu, Lycopene inhibits endothelial-to-mesenchymal transition of choroidal vascular endothelial cells in laser-induced mouse choroidal neovascularization. J. Cell. Mol. Med. 27, 1327– 1340 (2023).

83. G. E. Hoffman, P. Roussos, Dream: powerful differential expression analysis for repeated measures designs. Bioinformatics 37, 192–201 (2021).

84. J. W. Squair, M. Gautier, C. Kathe, M. A. Anderson, N. D. James, T. H. Hutson, R. Hudelle, T. Qaiser, K. J. E. Matson, Q. Barraud, A. J. Levine, G. La Manno, M. A. Skinnider, G. Courtine, Confronting false discoveries in single-cell differential expression. Nat. Commun. 12, 5692 (2021).

85. H. L. Crowell, C. Soneson, P.-L. Germain, D. Calini, L. Collin, C. Raposo, D. Malhotra, M. D. Robinson, muscat detects subpopulation-specific state transitions from multi-sample multi-condition single-cell transcriptomics data. Nat. Commun. 11, 6077 (2020).

86. D. Wu, G. K. Smyth, Camera: a competitive gene set test accounting for inter-gene correlation. Nucleic Acids Res. 40, e133 (2012).

87. D. Tingley, T. Yamamoto, K. Hirose, L. Keele, K. Imai, Mediation:RPackage for causal mediation analysis. J. Stat. Softw. 59, 1–38 (2014).

88. Z. Fang, X. Liu, G. Peltz, GSEApy: a comprehensive package for performing gene set enrichment analysis in Python. Bioinformatics 39 (2023).

89. B. Reuter, K. Fackeldey, M. Weber, Generalized Markov modeling of nonreversible molecular kinetics. J. Chem. Phys. 150, 174103 (2019).

90. L. Haghverdi, M. Büttner, F. A. Wolf, F. Buettner, F. J. Theis, Diffusion pseudotime robustly reconstructs lineage branching. Nat. Methods 13, 845–848 (2016).

91. L. Heumos, A. C. Schaar, C. Lance, A. Litinetskaya, F. Drost, L. Zappia, M. D. Lücken, D. C. Strobl, J. Henao, F. Curion, Single-cell Best Practices Consortium, H. B. Schiller, F. J. Theis, Best practices for single-cell analysis across modalities. Nat. Rev. Genet., 1–23 (2023).

92. R. Kolde, S. Laur, P. Adler, J. Vilo, Robust rank aggregation for gene list integration and meta-analysis. Bioinformatics 28, 573–580 (2012).

93. M. Efremova, M. Vento-Tormo, S. A. Teichmann, R. Vento-Tormo, CellPhoneDB: inferring cell-cell communication from combined expression of multi-subunit ligand-receptor complexes. Nat. Protoc. 15, 1484–1506 (2020).

94. M. S. B. Raredon, J. Yang, J. Garritano, M. Wang, D. Kushnir, J. C. Schupp, T. S. Adams, A. M. Greaney, K. L. Leiby, N. Kaminski, Y. Kluger, A. Levchenko, L. E. Niklason, Computation and visualization of cell-cell signaling topologies in single-cell systems data using Connectome. Sci. Rep. 12, 4187 (2022).

95. R. Hou, E. Denisenko, H. T. Ong, J. A. Ramilowski, A. R. R. Forrest, Predicting cell-to-cell communication networks using NATMI. Nat. Commun. 11, 5011 (2020).

96. S. Cabello-Aguilar, M. Alame, F. Kon-Sun-Tack, C. Fau, M. Lacroix, J. Colinge, SingleCellSignalR: inference of intercellular networks from single-cell transcriptomics. Nucleic Acids Res. 48, e55 (2020).

97. S. Jin, C. F. Guerrero-Juarez, L. Zhang, I. Chang, R. Ramos, C.-H. Kuan, P. Myung, M. V. Plikus, Q. Nie, Inference and analysis of cell-cell communication using CellChat. Nat. Commun. 12, 1088 (2021).

98. E. Armingol, H. M. Baghdassarian, C. Martino, A. Perez-Lopez, C. Aamodt, R. Knight, N. E. Lewis, Context-aware deconvolution of cell–cell communication with Tensor-cell2cell. Nat. Commun. 13, 1–15 (2022).

99. M. Ashburner, C. A. Ball, J. A. Blake, D. Botstein, H. Butler, J. M. Cherry, A. P. Davis, K. Dolinski, S. S. Dwight, J. T. Eppig, M. A. Harris, D. P. Hill, L. Issel-Tarver, A. Kasarskis, S. Lewis, J. C. Matese, J. E. Richardson, M. Ringwald, G. M. Rubin, G. Sherlock, Gene ontology: tool for the unification of biology. The Gene Ontology Consortium. Nat. Genet. 25, 25–29 (2000).

100. Gene Ontology Consortium, S. A. Aleksander, J. Balhoff, S. Carbon, J. M. Cherry, H. J. Drabkin, D. Ebert, M. Feuermann, P. Gaudet, N. L. Harris, D. P. Hill, R. Lee, H. Mi, S. Moxon, C. J. Mungall, A. Muruganugan, T. Mushayahama, P. W. Sternberg, P. D. Thomas, K. Van Auken, J. Ramsey, D. A. Siegele, R. L. Chisholm, P. Fey, M. C. Aspromonte, M. V. Nugnes, F. Quaglia, S. Tosatto, M. Giglio, S. Nadendla, G. Antonazzo, H. Attrill, G. Dos Santos, S. Marygold, V. Strelets, C. J. Tabone, J. Thurmond, P. Zhou, S. H. Ahmed, P. Asanitthong, D. Luna Buitrago, M. N. Erdol, M. C. Gage, M. Ali Kadhum, K. Y. C. Li, M. Long, A. Michalak, A. Pesala, A. Pritazahra, S. C. C. Saverimuttu, R. Su, K. E. Thurlow, R. C. Lovering, C. Logie, S. Oliferenko, J. Blake, K. Christie, L. Corbani, M. E. Dolan, H. J. Drabkin, D. P. Hill, L. Ni, D. Sitnikov, C. Smith, A. Cuzick, J. Seager, L. Cooper, J. Elser, P. Jaiswal, P. Gupta, P. Jaiswal, S. Naithani, M. Lera-Ramirez, K. Rutherford, V. Wood, J. L. De Pons, M. R. Dwinell, G. T. Hayman, M. L. Kaldunski, A. E. Kwitek, S. J. F. Laulederkind, M. A. Tutaj, M. Vedi, S.-J. Wang, P. D’Eustachio, L. Aimo, K. Axelsen, A. Bridge, N. Hyka-Nouspikel, A. Morgat, S. A. Aleksander, J. M. Cherry, S. R. Engel, K. Karra, S. R. Miyasato, R. S. Nash, M. S. Skrzypek, S. Weng, E. D. Wong, E. Bakker, T. Z. Berardini, L. Reiser, A. Auchincloss, K. Axelsen, G. Argoud-Puy, M.-C. Blatter, E. Boutet, L. Breuza, A. Bridge, C. Casals-Casas, E. Coudert, A. Estreicher, M. Livia Famiglietti, M. Feuermann, A. Gos, N. Gruaz-Gumowski, C. Hulo, N. Hyka-Nouspikel, F. Jungo, P. Le Mercier, D. Lieberherr, P. Masson, A. Morgat, I. Pedruzzi, L. Pourcel, S. Poux, C. Rivoire, S. Sundaram, A. Bateman, E. Bowler-Barnett, H. Bye-A-Jee, P. Denny, A. Ignatchenko, R. Ishtiaq, A. Lock, Y. Lussi, M. Magrane, M. J. Martin, S. Orchard, P. Raposo, E. Speretta, N. Tyagi, K. Warner, R. Zaru, A. D. Diehl, R. Lee, J. Chan, S. Diamantakis, D. Raciti, M. Zarowiecki, M. Fisher, C. James-Zorn, V. Ponferrada, A. Zorn, S. Ramachandran, L. Ruzicka, M. Westerfield, The Gene Ontology knowledgebase in 2023. Genetics 224 (2023).

101. International Multiple Sclerosis Genetics Consortium, Multiple sclerosis genomic map implicates peripheral immune cells and microglia in susceptibility. Science 365 (2019).

102. M. A. Nalls, C. Blauwendraat, C. L. Vallerga, K. Heilbron, S. Bandres-Ciga, D. Chang, M. Tan, D. A. Kia, A. J. Noyce, A. Xue, J. Bras, E. Young, R. von Coelln, J. Simón-Sánchez, C. Schulte, M. Sharma, L. Krohn, L. Pihlstrøm, A. Siitonen, H. Iwaki, H. Leonard, F. Faghri, J. R. Gibbs, D. G. Hernandez, S. W. Scholz, J. A. Botia, M. Martinez, J.-C. Corvol, S. Lesage, J. Jankovic, L. M. Shulman, M. Sutherland, P. Tienari, K. Majamaa, M. Toft, O. A. Andreassen, T. Bangale, A. Brice, J. Yang, Z. Gan-Or, T. Gasser, P. Heutink, J. M. Shulman, N. W. Wood, D. A. Hinds, J. A. Hardy, H. R. Morris, J. Gratten, P. M. Visscher, R. R. Graham, A. B. Singleton, 23andMe Research Team, System Genomics of Parkinson’s Disease Consortium, International Parkinson’s Disease Genomics Consortium, Identification of novel risk loci, causal insights, and heritable risk for Parkinson’s disease: a meta-analysis of genome-wide association studies. Lancet Neurol. 18, 1091–1102 (2019).

103. T. D. Als, M. I. Kurki, J. Grove, G. Voloudakis, K. Therrien, E. Tasanko, T. T. Nielsen, J. Naamanka, K. Veerapen, D. F. Levey, J. Bendl, J. Bybjerg-Grauholm, B. Zeng, D. Demontis, A. Rosengren, G. Athanasiadis, M. Bækved-Hansen, P. Qvist, G. Bragi Walters, T. Thorgeirsson, H. Stefánsson, K. L. Musliner, V. M. Rajagopal, L. Farajzadeh, J. Thirstrup, B. J. Vilhjálmsson, J. J. McGrath, M. Mattheisen, S. Meier, E. Agerbo, K. Stefánsson, M. Nordentoft, T. Werge, D. M. Hougaard, P. B. Mortensen, M. B. Stein, J. Gelernter, I. Hovatta, P. Roussos, M. J. Daly, O. Mors, A. Palotie, A. D. Børglum, Depression pathophysiology, risk prediction of recurrence and comorbid psychiatric disorders using genome-wide analyses. Nat. Med. 29, 1832–1844 (2023).

104. J. Grove, S. Ripke, T. D. Als, M. Mattheisen, R. K. Walters, H. Won, J. Pallesen, E. Agerbo, O. A. Andreassen, R. Anney, Others, Identification of common genetic risk variants for autism spectrum disorder. Nat. Genet. 51, 431–444 (2019).

105. N. Mullins, A. J. Forstner, K. S. O’Connell, B. Coombes, J. R. I. Coleman, Z. Qiao, T. D. Als, T. B. Bigdeli, S. Børte, J. Bryois, A. W. Charney, O. K. Drange, M. J. Gandal, S. P. Hagenaars, M. Ikeda, N. Kamitaki, M. Kim, K. Krebs, G. Panagiotaropoulou, B. M. Schilder, L. G. Sloofman, S. Steinberg, V. Trubetskoy, B. S. Winsvold, H.-H. Won, L. Abramova, K. Adorjan, E. Agerbo, M. Al Eissa, D. Albani, N. Alliey-Rodriguez, A. Anjorin, V. Antilla, A. Antoniou, S. Awasthi, J. H. Baek, M. Bækvad-Hansen, N. Bass, M. Bauer, E. C. Beins, S. E. Bergen, A. Birner, C. Bøcker Pedersen, E. Bøen, M. P. Boks, R. Bosch, M. Brum, B. M. Brumpton, N. Brunkhorst-Kanaan, M. Budde, J. Bybjerg-Grauholm, W. Byerley, M. Cairns, M. Casas, P. Cervantes, T.-K. Clarke, C. Cruceanu, A. Cuellar-Barboza, J. Cunningham, D. Curtis, P. M. Czerski, A. M. Dale, N. Dalkner, F. S. David, F. Degenhardt, S. Djurovic, A. L. Dobbyn, A. Douzenis, T. Elvsåshagen, V. Escott-Price, I. N. Ferrier, A. Fiorentino, T. M. Foroud, L. Forty, J. Frank, O. Frei, N. B. Freimer, L. Frisén, K. Gade, J. Garnham, J. Gelernter, M. Giørtz Pedersen, I. R. Gizer, S. D. Gordon, K. Gordon-Smith, T. A. Greenwood, J. Grove, J. Guzman-Parra, K. Ha, M. Haraldsson, M. Hautzinger, U. Heilbronner, D. Hellgren, S. Herms, P. Hoffmann, P. A. Holmans, L. Huckins, S. Jamain, J. S. Johnson, J. L. Kalman, Y. Kamatani, J. L. Kennedy, S. Kittel-Schneider, J. A. Knowles, M. Kogevinas, M. Koromina, T. M. Kranz, H. R. Kranzler, M. Kubo, R. Kupka, S. A. Kushner, C. Lavebratt, J. Lawrence, M. Leber, H.-J. Lee, P. H. Lee, S. E. Levy, C. Lewis, C. Liao, S. Lucae, M. Lundberg, D. J. MacIntyre, S. H. Magnusson, W. Maier, A. Maihofer, D. Malaspina, E. Maratou, L. Martinsson, M. Mattheisen, S. A. McCarroll, N. W. McGregor, P. McGuffin, J. D. McKay, H. Medeiros, S. E. Medland, V. Millischer, G. W. Montgomery, J. L. Moran, D. W. Morris, T. W. Mühleisen, N. O’Brien, C. O’Donovan, L. M. Olde Loohuis, L. Oruc, S. Papiol, A. F. Pardiñas, A. Perry, A. Pfennig, E. Porichi, J. B. Potash, D. Quested, T. Raj, M. H. Rapaport, J. R. DePaulo, E. J. Regeer, J. P. Rice, F. Rivas, M. Rivera, J. Roth, P. Roussos, D. M. Ruderfer, C. Sánchez-Mora, E. C. Schulte, F. Senner, S. Sharp, P. D. Shilling, E. Sigurdsson, L. Sirignano, C. Slaney, O. B. Smeland, D. J. Smith, J. L. Sobell, C. Søholm Hansen, M. Soler Artigas, A. T. Spijker, D. J. Stein, J. S. Strauss, B. Świątkowska, C. Terao, T. E. Thorgeirsson, C. Toma, P. Tooney, E.-E. Tsermpini, M. P. Vawter, H. Vedder, J. T. R. Walters, S. H. Witt, S. Xi, W. Xu, J. M. K. Yang, A. H. Young, H. Young, P. P. Zandi, H. Zhou, L. Zillich, HUNT All-In Psychiatry, R. Adolfsson, I. Agartz, M. Alda, L. Alfredsson, G. Babadjanova, L. Backlund, B. T. Baune, F. Bellivier, S. Bengesser, W. H. Berrettini, D. H. R. Blackwood, M. Boehnke, A. D. Børglum, G. Breen, V. J. Carr, S. Catts, A. Corvin, N. Craddock, U. Dannlowski, D. Dikeos, T. Esko, B. Etain, P. Ferentinos, M. Frye, J. M. Fullerton, M. Gawlik, E. S. Gershon, F. S. Goes, M. J. Green, M. Grigoroiu-Serbanescu, J. Hauser, F. Henskens, J. Hillert, K. S. Hong, D. M. Hougaard, C. M. Hultman, K. Hveem, N. Iwata, A. V. Jablensky, I. Jones, L. A. Jones, R. S. Kahn, J. R. Kelsoe, G. Kirov, M. Landén, M. Leboyer, C. M. Lewis, Q. S. Li, J. Lissowska, C. Lochner, C. Loughland, N. G. Martin, C. A. Mathews, F. Mayoral, S. L. McElroy, A. M. McIntosh, F. J. McMahon, I. Melle, P. Michie, L. Milani, P. B. Mitchell, G. Morken, O. Mors, P. B. Mortensen, B. Mowry, B. Müller-Myhsok, R. M. Myers, B. M. Neale, C. M. Nievergelt, M. Nordentoft, M. M. Nöthen, M. C. O’Donovan, K. J. Oedegaard, T. Olsson, M. J. Owen, S. A. Paciga, C. Pantelis, C. Pato, M. T. Pato, G. P. Patrinos, R. H. Perlis, D. Posthuma, J. A. Ramos-Quiroga, A. Reif, E. Z. Reininghaus, M. Ribasés, M. Rietschel, S. Ripke, G. A. Rouleau, T. Saito, U. Schall, M. Schalling, P. R. Schofield, T. G. Schulze, L. J. Scott, R. J. Scott, A. Serretti, C. Shannon Weickert, J. W. Smoller, H. Stefansson, K. Stefansson, E. Stordal, F. Streit, P. F. Sullivan, G. Turecki, A. E. Vaaler, E. Vieta, J. B. Vincent, I. D. Waldman, T. W. Weickert, T. Werge, N. R. Wray, J.-A. Zwart, J. M. Biernacka, J. I. Nurnberger, S. Cichon, H. J. Edenberg, E. A. Stahl, A. McQuillin, A. Di Florio, R. A. Ophoff, O. A. Andreassen, Genome-wide association study of more than 40,000 bipolar disorder cases provides new insights into the underlying biology. Nat. Genet. 53, 817–829 (2021).

106. V. Trubetskoy, A. F. Pardiñas, T. Qi, G. Panagiotaropoulou, S. Awasthi, T. B. Bigdeli, J. Bryois, C.-Y. Chen, C. A. Dennison, L. S. Hall, M. Lam, K. Watanabe, O. Frei, T. Ge, J. C. Harwood, F. Koopmans, S. Magnusson, A. L. Richards, J. Sidorenko, Y. Wu, J. Zeng, J. Grove, M. Kim, Z. Li, G. Voloudakis, W. Zhang, M. Adams, I. Agartz, E. G. Atkinson, E. Agerbo, M. Al Eissa, M. Albus, M. Alexander, B. Z. Alizadeh, K. Alptekin, T. D. Als, F. Amin, V. Arolt, M. Arrojo, L. Athanasiu, M. H. Azevedo, S. A. Bacanu, N. J. Bass, M. Begemann, R. A. Belliveau, J. Bene, B. Benyamin, S. E. Bergen, G. Blasi, J. Bobes, S. Bonassi, A. Braun, R. A. Bressan, E. J. Bromet, R. Bruggeman, P. F. Buckley, R. L. Buckner, J. Bybjerg-Grauholm, W. Cahn, M. J. Cairns, M. E. Calkins, V. J. Carr, D. Castle, S. V. Catts, K. D. Chambert, R. C. K. Chan, B. Chaumette, W. Cheng, E. F. C. Cheung, S. A. Chong, D. Cohen, A. Consoli, Q. Cordeiro, J. Costas, C. Curtis, M. Davidson, K. L. Davis, L. de Haan, F. Degenhardt, L. E. DeLisi, D. Demontis, F. Dickerson, D. Dikeos, T. Dinan, S. Djurovic, J. Duan, G. Ducci, F. Dudbridge, J. G. Eriksson, L. Fañanás, S. V. Faraone, A. Fiorentino, A. Forstner, J. Frank, N. B. Freimer, M. Fromer, A. Frustaci, A. Gadelha, G. Genovese, E. S. Gershon, M. Giannitelli, I. Giegling, P. Giusti-Rodríguez, S. Godard, J. I. Goldstein, J. González Peñas, A. González-Pinto, S. Gopal, J. Gratten, M. F. Green, T. A. Greenwood, O. Guillin, S. Gülöksüz, R. E. Gur, R. C. Gur, B. Gutiérrez, E. Hahn, H. Hakonarson, V. Haroutunian, A. M. Hartmann, C. Harvey, C. Hayward, F. A. Henskens, S. Herms, P. Hoffmann, D. P. Howrigan, M. Ikeda, C. Iyegbe, I. Joa, A. Julià, A. K. Kähler, T. Kam-Thong, Y. Kamatani, S. Karachanak-Yankova, O. Kebir, M. C. Keller, B. J. Kelly, A. Khrunin, S.-W. Kim, J. Klovins, N. Kondratiev, B. Konte, J. Kraft, M. Kubo, V. Kučinskas, Z. A. Kučinskiene, A. Kusumawardhani, H. Kuzelova-Ptackova, S. Landi, L. C. Lazzeroni, P. H. Lee, S. E. Legge, D. S. Lehrer, R. Lencer, B. Lerer, M. Li, J. Lieberman, G. A. Light, S. Limborska, C.-M. Liu, J. Lönnqvist, C. M. Loughland, J. Lubinski, J. J. Luykx, A. Lynham, M. Macek Jr, A. Mackinnon, P. K. E. Magnusson, B. S. Maher, W. Maier, D. Malaspina, J. Mallet, S. R. Marder, S. Marsal, A. R. Martin, L. Martorell, M. Mattheisen, R. W. McCarley, C. McDonald, J. J. McGrath, H. Medeiros, S. Meier, B. Melegh, I. Melle, R. I. Mesholam-Gately, A. Metspalu, P. T. Michie, L. Milani, V. Milanova, M. Mitjans, E. Molden, E. Molina, M. D. Molto, V. Mondelli, C. Moreno, C. P. Morley, G. Muntané, K. C. Murphy, I. Myin-Germeys, I. Nenadić, G. Nestadt, L. Nikitina-Zake, C. Noto, K. H. Nuechterlein, N. L. O’Brien, F. A. O’Neill, S.-Y. Oh, A. Olincy, V. K. Ota, C. Pantelis, G. N. Papadimitriou, M. Parellada, T. Paunio, R. Pellegrino, S. Periyasamy, D. O. Perkins, B. Pfuhlmann, O. Pietiläinen, J. Pimm, D. Porteous, J. Powell, D. Quattrone, D. Quested, A. D. Radant, A. Rampino, M. H. Rapaport, A. Rautanen, A. Reichenberg, C. Roe, J. L. Roffman, J. Roth, M. Rothermundt, B. P. F. Rutten, S. Saker-Delye, V. Salomaa, J. Sanjuan, M. L. Santoro, A. Savitz, U. Schall, R. J. Scott, L. J. Seidman, S. I. Sharp, J. Shi, L. J. Siever, E. Sigurdsson, K. Sim, N. Skarabis, P. Slominsky, H.-C. So, J. L. Sobell, E. Söderman, H. J. Stain, N. E. Steen, A. A. Steixner-Kumar, E. Stögmann, W. S. Stone, R. E. Straub, F. Streit, E. Strengman, T. S. Stroup, M. Subramaniam, C. A. Sugar, J. Suvisaari, D. M. Svrakic, N. R. Swerdlow, J. P. Szatkiewicz, T. M. T. Ta, A. Takahashi, C. Terao, F. Thibaut, D. Toncheva, P. A. Tooney, S. Torretta, S. Tosato, G. B. Tura, B. I. Turetsky, A. Üçok, A. Vaaler, T. van Amelsvoort, R. van Winkel, J. Veijola, J. Waddington, H. Walter, A. Waterreus, B. T. Webb, M. Weiser, N. M. Williams, S. H. Witt, B. K. Wormley, J. Q. Wu, Z. Xu, R. Yolken, C. C. Zai, W. Zhou, F. Zhu, F. Zimprich, E. C. Atbaşoğlu, M. Ayub, C. Benner, A. Bertolino, D. W. Black, N. J. Bray, G. Breen, N. G. Buccola, W. F. Byerley, W. J. Chen, C. R. Cloninger, B. Crespo-Facorro, G. Donohoe, R. Freedman, C. Galletly, M. J. Gandal, M. Gennarelli, D. M. Hougaard, H.-G. Hwu, A. V. Jablensky, S. A. McCarroll, J. L. Moran, O. Mors, P. B. Mortensen, B. Müller-Myhsok, A. L. Neil, M. Nordentoft, M. T. Pato, T. L. Petryshen, M. Pirinen, A. E. Pulver, T. G. Schulze, J. M. Silverman, J. W. Smoller, E. A. Stahl, D. W. Tsuang, E. Vilella, S.-H. Wang, S. Xu, Indonesia Schizophrenia Consortium, PsychENCODE, Psychosis Endophenotypes International Consortium, SynGO Consortium, R. Adolfsson, C. Arango, B. T. Baune, S. I. Belangero, A. D. Børglum, D. Braff, E. Bramon, J. D. Buxbaum, D. Campion, J. A. Cervilla, S. Cichon, D. A. Collier, A. Corvin, D. Curtis, M. D. Forti, E. Domenici, H. Ehrenreich, V. Escott-Price, T. Esko, A. H. Fanous, A. Gareeva, M. Gawlik, P. V. Gejman, M. Gill, S. J. Glatt, V. Golimbet, K. S. Hong, C. M. Hultman, S. E. Hyman, N. Iwata, E. G. Jönsson, R. S. Kahn, J. L. Kennedy, E. Khusnutdinova, G. Kirov, J. A. Knowles, M.-O. Krebs, C. Laurent-Levinson, J. Lee, T. Lencz, D. F. Levinson, Q. S. Li, J. Liu, A. K. Malhotra, D. Malhotra, A. McIntosh, A. McQuillin, P. R. Menezes, V. A. Morgan, D. W. Morris, B. J. Mowry, R. M. Murray, V. Nimgaonkar, M. M. Nöthen, R. A. Ophoff, S. A. Paciga, A. Palotie, C. N. Pato, S. Qin, M. Rietschel, B. P. Riley, M. Rivera, D. Rujescu, M. C. Saka, A. R. Sanders, S. G. Schwab, A. Serretti, P. C. Sham, Y. Shi, D. St Clair, H. Stefánsson, K. Stefansson, M. T. Tsuang, J. van Os, M. P. Vawter, D. R. Weinberger, T. Werge, D. B. Wildenauer, X. Yu, W. Yue, P. A. Holmans, A. J. Pocklington, P. Roussos, E. Vassos, M. Verhage, P. M. Visscher, J. Yang, D. Posthuma, O. A. Andreassen, K. S. Kendler, M. J. Owen, N. R. Wray, M. J. Daly, H. Huang, B. M. Neale, P. F. Sullivan, S. Ripke, J. T. R. Walters, M. C. O’Donovan, Schizophrenia Working Group of the Psychiatric Genomics Consortium, Mapping genomic loci implicates genes and synaptic biology in schizophrenia. Nature 604, 502–508 (2022).

107. W. van Rheenen, R. A. A. van der Spek, M. K. Bakker, J. J. F. A. van Vugt, P. J. Hop, R. A. J. Zwamborn, N. de Klein, H.-J. Westra, O. B. Bakker, P. Deelen, G. Shireby, E. Hannon, M. Moisse, D. Baird, R. Restuadi, E. Dolzhenko, A. M. Dekker, K. Gawor, H.-J. Westeneng, G. H. P. Tazelaar, K. R. van Eijk, M. Kooyman, R. P. Byrne, M. Doherty, M. Heverin, A. Al Khleifat, A. Iacoangeli, A. Shatunov, N. Ticozzi, J. Cooper-Knock, B. N. Smith, M. Gromicho, S. Chandran, S. Pal, K. E. Morrison, P. J. Shaw, J. Hardy, R. W. Orrell, M. Sendtner, T. Meyer, N. Başak, A. J. van der Kooi, A. Ratti, I. Fogh, C. Gellera, G. Lauria, S. Corti, C. Cereda, D. Sproviero, S. D’Alfonso, G. Sorarù, G. Siciliano, M. Filosto, A. Padovani, A. Chiò, A. Calvo, C. Moglia, M. Brunetti, A. Canosa, M. Grassano, E. Beghi, E. Pupillo, G. Logroscino, B. Nefussy, A. Osmanovic, A. Nordin, Y. Lerner, M. Zabari, M. Gotkine, R. H. Baloh, S. Bell, P. Vourc’h, P. Corcia, P. Couratier, S. Millecamps, V. Meininger, F. Salachas, J. S. Mora Pardina, A. Assialioui, R. Rojas-García, P. A. Dion, J. P. Ross, A. C. Ludolph, J. H. Weishaupt, D. Brenner, A. Freischmidt, G. Bensimon, A. Brice, A. Durr, C. A. M. Payan, S. Saker-Delye, N. W. Wood, S. Topp, R. Rademakers, L. Tittmann, W. Lieb, A. Franke, S. Ripke, A. Braun, J. Kraft, D. C. Whiteman, C. M. Olsen, A. G. Uitterlinden, A. Hofman, M. Rietschel, S. Cichon, M. M. Nöthen, P. Amouyel, SLALOM Consortium, PARALS Consortium, SLAGEN Consortium, SLAP Consortium, B. J. Traynor, A. B. Singleton, M. Mitne Neto, R. J. Cauchi, R. A. Ophoff, M. Wiedau-Pazos, C. Lomen-Hoerth, V. M. van Deerlin, J. Grosskreutz, A. Roediger, N. Gaur, A. Jörk, T. Barthel, E. Theele, B. Ilse, B. Stubendorff, O. W. Witte, R. Steinbach, C. A. Hübner, C. Graff, L. Brylev, V. Fominykh, V. Demeshonok, A. Ataulina, B. Rogelj, B. Koritnik, J. Zidar, M. Ravnik-Glavač, D. Glavač, Z. Stević, V. Drory, M. Povedano, I. P. Blair, M. C. Kiernan, B. Benyamin, R. D. Henderson, S. Furlong, S. Mathers, P. A. McCombe, M. Needham, S. T. Ngo, G. A. Nicholson, R. Pamphlett, D. B. Rowe, F. J. Steyn, K. L. Williams, K. A. Mather, P. S. Sachdev, A. K. Henders, L. Wallace, M. de Carvalho, S. Pinto, S. Petri, M. Weber, G. A. Rouleau, V. Silani, C. J. Curtis, G. Breen, J. D. Glass, R. H. Brown Jr, J. E. Landers, C. E. Shaw, P. M. Andersen, E. J. N. Groen, M. A. van Es, R. J. Pasterkamp, D. Fan, F. C. Garton, A. F. McRae, G. Davey Smith, T. R. Gaunt, M. A. Eberle, J. Mill, R. L. McLaughlin, O. Hardiman, K. P. Kenna, N. R. Wray, E. Tsai, H. Runz, L. Franke, A. Al-Chalabi, P. Van Damme, L. H. van den Berg, J. H. Veldink, Common and rare variant association analyses in amyotrophic lateral sclerosis identify 15 risk loci with distinct genetic architectures and neuron-specific biology. Nat. Genet. 53, 1636–1648 (2021).

108. D. Taliun, D. N. Harris, M. D. Kessler, J. Carlson, Z. A. Szpiech, R. Torres, S. A. G. Taliun, A. Corvelo, S. M. Gogarten, H. M. Kang, A. N. Pitsillides, J. LeFaive, S.-B. Lee, X. Tian, B. L. Browning, S. Das, A.-K. Emde, W. E. Clarke, D. P. Loesch, A. C. Shetty, T. W. Blackwell, A. V. Smith, Q. Wong, X. Liu, M. P. Conomos, D. M. Bobo, F. Aguet, C. Albert, A. Alonso, K. G. Ardlie, D. E. Arking, S. Aslibekyan, P. L. Auer, J. Barnard, R. G. Barr, L. Barwick, L. C. Becker, R. L. Beer, E. J. Benjamin, L. F. Bielak, J. Blangero, M. Boehnke, D. W. Bowden, J. A. Brody, E. G. Burchard, B. E. Cade, J. F. Casella, B. Chalazan, D. I. Chasman, Y.-D. I. Chen, M. H. Cho, S. H. Choi, M. K. Chung, C. B. Clish, A. Correa, J. E. Curran, B. Custer, D. Darbar, M. Daya, M. de Andrade, D. L. DeMeo, S. K. Dutcher, P. T. Ellinor, L. S. Emery, C. Eng, D. Fatkin, T. Fingerlin, L. Forer, M. Fornage, N. Franceschini, C. Fuchsberger, S. M. Fullerton, S. Germer, M. T. Gladwin, D. J. Gottlieb, X. Guo, M. E. Hall, J. He, N. L. Heard-Costa, S. R. Heckbert, M. R. Irvin, J. M. Johnsen, A. D. Johnson, R. Kaplan, S. L. R. Kardia, T. Kelly, S. Kelly, E. E. Kenny, D. P. Kiel, R. Klemmer, B. A. Konkle, C. Kooperberg, A. Köttgen, L. A. Lange, J. Lasky-Su, D. Levy, X. Lin, K.-H. Lin, C. Liu, R. J. F. Loos, L. Garman, R. Gerszten, S. A. Lubitz, K. L. Lunetta, A. C. Y. Mak, A. Manichaikul, A. K. Manning, R. A. Mathias, D. D. McManus, S. T. McGarvey, J. B. Meigs, D. A. Meyers, J. L. Mikulla, M. A. Minear, B. D. Mitchell, S. Mohanty, M. E. Montasser, C. Montgomery, A. C. Morrison, J. M. Murabito, A. Natale, P. Natarajan, S. C. Nelson, K. E. North, J. R. O’Connell, N. D. Palmer, N. Pankratz, G. M. Peloso, P. A. Peyser, J. Pleiness, W. S. Post, B. M. Psaty, D. C. Rao, S. Redline, A. P. Reiner, D. Roden, J. I. Rotter, I. Ruczinski, C. Sarnowski, S. Schoenherr, D. A. Schwartz, J.-S. Seo, S. Seshadri, V. A. Sheehan, W. H. Sheu, M. B. Shoemaker, N. L. Smith, J. A. Smith, N. Sotoodehnia, A. M. Stilp, W. Tang, K. D. Taylor, M. Telen, T. A. Thornton, R. P. Tracy, D. J. Van Den Berg, R. S. Vasan, K. A. Viaud-Martinez, S. Vrieze, D. E. Weeks, B. S. Weir, S. T. Weiss, L.-C. Weng, C. J. Willer, Y. Zhang, X. Zhao, D. K. Arnett, A. E. Ashley-Koch, K. C. Barnes, E. Boerwinkle, S. Gabriel, R. Gibbs, K. M. Rice, S. S. Rich, E. K. Silverman, P. Qasba, W. Gan, NHLBI Trans-Omics for Precision Medicine (TOPMed) Consortium, G. J. Papanicolaou, D. A. Nickerson, S. R. Browning, M. C. Zody, S. Zöllner, J. G. Wilson, L. A. Cupples, C. C. Laurie, C. E. Jaquish, R. D. Hernandez, T. D. O’Connor, G. R. Abecasis, Sequencing of 53,831 diverse genomes from the NHLBI TOPMed Program. Nature 590, 290–299 (2021).

109. A. Manichaikul, J. C. Mychaleckyj, S. S. Rich, K. Daly, M. Sale, W.-M. Chen, Robust relationship inference in genome-wide association studies. Bioinformatics 26, 2867–2873 (2010).

110. 110. 1000 Genomes Project Consortium, G. R. Abecasis, A. Auton, L. D. Brooks, M. A. DePristo, R. M. Durbin, R. E. Handsaker, H. M. Kang, G. T. Marth, G. A. McVean, An integrated map of genetic variation from 1,092 human genomes. Nature 491, 56–65 (2012).

111. 111. 1000 Genomes Project Consortium, A. Auton, L. D. Brooks, R. M. Durbin, E. P. Garrison, H. M. Kang, J. O. Korbel, J. L. Marchini, S. McCarthy, G. A. McVean, G. R. Abecasis, A global reference for human genetic variation. Nature 526, 68–74 (2015).

112. P. Danecek, J. K. Bonfield, J. Liddle, J. Marshall, V. Ohan, M. O. Pollard, A. Whitwham, T. Keane, S. A. McCarthy, R. M. Davies, H. Li, Twelve years of SAMtools and BCFtools. Gigascience 10 (2021).

113. C. C. Chang, C. C. Chow, L. C. Tellier, S. Vattikuti, S. M. Purcell, J. J. Lee, Second-generation PLINK: rising to the challenge of larger and richer datasets. Gigascience 4, 7 (2015).

114. T. Ge, C.-Y. Chen, Y. Ni, Y.-C. A. Feng, J. W. Smoller, Polygenic prediction via Bayesian regression and continuous shrinkage priors. Nat. Commun. 10, 1776 (2019).

115. C. J. Bohlen, F. C. Bennett, A. F. Tucker, H. Y. Collins, S. B. Mulinyawe, B. A. Barres, Diverse Requirements for Microglial Survival, Specification, and Function Revealed by Defined-Medium Cultures. Neuron 94, 759–773.e8 (2017).

116. D. Gosselin, D. Skola, N. G. Coufal, I. R. Holtman, J. C. M. Schlachetzki, E. Sajti, B. N. Jaeger, C. O’Connor, C. Fitzpatrick, M. P. Pasillas, M. Pena, A. Adair, D. D. Gonda, M. L. Levy, R. M. Ransohoff, F. H. Gage, C. K. Glass, An environment-dependent transcriptional network specifies human microglia identity. Science 356 (2017).

117. S. R. Ocañas, K. D. Pham, H. E. Blankenship, A. H. Machalinski, A. J. Chucair-Elliott, W. M. Freeman, Minimizing the Ex Vivo Confounds of Cell-Isolation Techniques on Transcriptomic and Translatomic Profiles of Purified Microglia. eNeuro 9 (2022).

118. S. C. van den Brink, F. Sage, Á. Vértesy, B. Spanjaard, J. Peterson-Maduro, C. S. Baron, C. Robin, A. van Oudenaarden, Single-cell sequencing reveals dissociation-induced gene expression in tissue subpopulations. Nat. Methods 14, 935–936 (2017).

119. Y. Lee, J. S. Lee, K. J. Lee, R. S. Turner, H.-S. Hoe, D. T. S. Pak, Polo-like kinase 2 phosphorylation of amyloid precursor protein regulates activity-dependent amyloidogenic processing. Neuropharmacology 117, 387–400 (2017).

120. J. S. Lee, Y. Lee, E. A. André, K. J. Lee, T. Nguyen, Y. Feng, N. Jia, B. T. Harris, M. P. Burns, D. T. S. Pak, Inhibition of Polo-like kinase 2 ameliorates pathogenesis in Alzheimer’s disease model mice. PLoS One 14, e0219691 (2019).

121. M. Setty, V. Kiseliovas, J. Levine, A. Gayoso, L. Mazutis, D. Pe’er, Characterization of cell fate probabilities in single-cell data with Palantir. Nat. Biotechnol. 37, 451–460 (2019).

122. K. Street, D. Risso, R. B. Fletcher, D. Das, J. Ngai, N. Yosef, E. Purdom, S. Dudoit, Slingshot: cell lineage and pseudotime inference for single-cell transcriptomics. BMC Genomics 19, 477 (2018).

123. G. Schiebinger, J. Shu, M. Tabaka, B. Cleary, V. Subramanian, A. Solomon, J. Gould, S. Liu, S. Lin, P. Berube, L. Lee, J. Chen, J. Brumbaugh, P. Rigollet, K. Hochedlinger, R. Jaenisch, A. Regev, E. S. Lander, Optimal-Transport Analysis of Single-Cell Gene Expression Identifies Developmental Trajectories in Reprogramming. Cell 176, 928– 943.e22 (2019).

